# The expected polygenic risk score (ePRS) framework: an equitable metric for quantifying polygenetic risk via modeling of ancestral makeup

**DOI:** 10.1101/2024.03.05.24303738

**Authors:** Yu-Jyun Huang, Nuzulul Kurniansyah, Matthew O Goodman, Brian W Spitzer, Jiongming Wang, Adrienne Stilp, Cecelia Laurie, Paul S de Vries, Han Chen, Yuan-I Min, Mario Sims, Gina M Peloso, Xiuqing Guo, Joshua C Bis, Jennifer A Brody, Laura M Raffield, Jennifer A Smith, Wei Zhao, Jerome I Rotter, Stephen S Rich, Susan Redline, Myriam Fornage, Robert Kaplan, Nora Franceschini, Daniel Levy, Alanna C Morrison, Eric Boerwinkle, Nicholas L Smith, Charles Kooperberg, Bruce M Psaty, Sebastian Zöllner, the Trans-Omics in Precision Medicine Consortium, Tamar Sofer

## Abstract

Polygenic risk scores (PRSs) depend on genetic ancestry due to differences in allele frequencies between ancestral populations. This leads to implementation challenges in diverse populations. We propose a framework to calibrate PRS based on ancestral makeup. We define a metric called “expected PRS” (ePRS), the expected value of a PRS based on one’s global or local admixture patterns. We further define the “residual PRS” (rPRS), measuring the deviation of the PRS from the ePRS. Simulation studies confirm that it suffices to adjust for ePRS to obtain nearly unbiased estimates of the PRS-outcome association without further adjusting for PCs. Using the TOPMed dataset, the estimated effect size of the rPRS adjusting for the ePRS is similar to the estimated effect of the PRS adjusting for genetic PCs. Similarly, we applied the ePRS framework to six cardiovascular-related traits in the All of Us dataset, and the results are consistent with those from the TOPMed analysis. The ePRS framework can protect from population stratification in association analysis and provide an equitable strategy to quantify genetic risk across diverse populations.

## Introduction

Polygenic risk scores (PRS) combine information from multiple genetic variants, summarizing disease risk due to genetics into a single score. They are typically constructed based on summary statistics from genome-wide association studies (GWAS). PRSs are now being incorporated into the clinic (1,2). Potential applications of PRS in healthcare settings include disease risk screening, risk prediction, and identification of target populations that may benefit from early interventions; all are the ultimate goals of precision and preventive medicine (3–5). Using PRS in multi-ancestry populations has received increasing attention due to the recognition that genomics research has been “failing on diversity” (6), with most genetic studies being carried out in populations of European ancestry. Consequently, PRS performance is lower in populations that are not of European ancestry or that have some European ancestry admixed with other continental ancestries (7). Aside from efforts to increase the diversity of GWAS participants, recent PRS research has proposed to improve upon PRS in individuals of diverse ancestries by combining information from summary statistics from multiple GWASs, each from a different genetic ancestry, while borrowing information across (8,9). However, there is still a gap in PRS performance between individuals of European ancestry and of other diverse populations (10,11).

To alleviate population stratification bias in association studies, adjusting for principal components (PCs) constructed using genome-wide genetic data is a standard procedure. Recently, the “ancestry-adjusted PRS” method was utilized to analyze diverse populations (4,12,13), where PCs are regressed out of the PRS via a linear model, and then the PRS-outcome association is estimated with the residuals from the first stage regression model. Khera et al. (4) showed that the ancestry-adjusted PRS had similar distribution across diverse populations, which indicated the removal of ancestry-related genetic effects. However, different studies may adjust for different number of PCs, and the meaning of PCs are inequivalent across different datasets. Furthermore, PCs can only capture global genetic structure and not local ancestry pattern. Hence, it still needs to be determined whether the adjustment of PCs in PRS-outcome association analyses in admixed population is sufficient to account for unknown ancestry-related confounding due to the complex admixture pattern (14–16).

Motivated by the challenges of using PCs in the translation of PRS for clinical use in diverse populations, we aim to propose a new method in which the ancestry-related factors that cause the differences in the distribution of PRS between populations can be captured by a single variable, and the interpretation is identical across datasets. The intuitive difference in PRS distribution between populations is that they have different baseline genetic characteristics, i.e., the mean of the PRS distribution, which is caused by the difference in ancestry composition and ancestry-specific allele frequency. Therefore, if the baseline genetic value of each individual can be computed, the corresponding PRS can be calibrated to obtain a distribution that is homogeneous across populations. Because the calibrated PRS is now independent of genetic ancestry, utilizing a single threshold to classify individuals into high-or low-risk groups across diverse populations is feasible, which achieves the goal of equitably quantifying genetic risk.

Based on the above rationale, this work introduces a novel framework that uses individual’s genetic ancestry composition in PRS construction. This framework can be applied in combination with many existing strategies to develop PRSs, as it does not deal with selection of variants and weights, but rather with the use of PRSs in ancestry-heterogenous populations. We propose a metric called expected PRS (ePRS), in which the expected value of a PRS is calculated based on either their global ancestry proportions or their local ancestry pattern. Therefore, the ePRS can be interpreted as the baseline trait-specific genetic parameter based on an individual’s ancestral composition. Coupled with ePRS, we introduce another two metrics: the residual PRS (rPRS), which measures the deviation of the PRS from its expected mean according to admixture patterns (i.e., from the ePRS), and the quantile PRS (qPRS), defining the distribution-based percentile of the PRS accounting for both the ePRS and its variability. Therefore, the rPRS and qPRS can distinguish genetic risk between individuals regardless of their genetic ancestry patterns. We performed simulation studies to examine whether our proposed method can provide a nearly unbiased estimation of the PRS-outcome effect by adjusting for ePRS in the association model. Other standard adjustment procedures are considered as comparators. We applied our method to analyze data from the Trans-Omics for Precision Medicine (TOPMed) study and compare the PRS-outcome effect estimation with the standard PC-adjusted approach. We also implemented the ePRS framework in the All of Us (AoU) research program to analyze six cardiovascular disease (CVD)-related traits, demonstrating the generalizability of our proposed method for multi-ancestry PRS analysis.

## Results

### PRS-outcome association model

The underlying PRS-outcome association model is provided by:

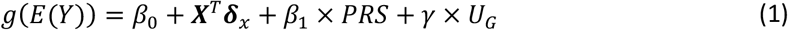

In equation (1), *Y* is the trait of interest. The PRS of individual 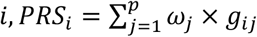, is calculated by the aggregation of SNP allele effect sizes ω_*j*_, *j* = 1, 2, …, *p* and the individual allele counts *g*_*ij*_, *j* = 1, 2, ‥, *p*. The parameter of interest is *β*_1_, representing the effect size of the PRS-outcome association; ***X*** is a vector of covariates and ***δ*** is the vector of corresponding effect sizes; and *U*_*G*_ is an unknown ancestry-related genetic factor, which may confound the PRS-outcome relationship (population stratification bias), and *γ* is its effect. The function *g*(·) is known as the link function, relating the outcome distribution to the covariates.

### Overview of the ePRS framework

We define the ePRS as the expectation of individual’s polygenic risk score *ePRS*_*i*_ = *E*(*PRS*_*i*_). The ePRS provides the baseline genetic characteristic based on one’s ancestry composition. We define the residual PRS (rPRS) as the difference between PRS and ePRS, i.e., *rPRS*_*i*_ = *PRS*_*i*_ – *ePRS*_*i*_. The rPRS can be used to compare genetic risk between individuals with different genetic ancestry composition. The working association model in the ePRS framework is illustrated in equation (2)

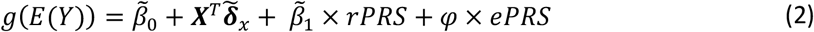

One can see that both the rPRS and the ePRS are used as covariates in the model. The estimated effect of the rPRS is equivalent to the estimated effect of the PRS in a standard association model. Importantly, the ePRS is used to control for stratification bias, instead of the standard adjustment for genetic PCs. This is illustrated in Figure 1a, where one can see that the ePRS blocks the “back door path” from the unknown genetic factors (*U*_*G*_ from equation (1)) that are related to ancestry by having ancestry-related differences in allele frequencies.

**Figure 1.**
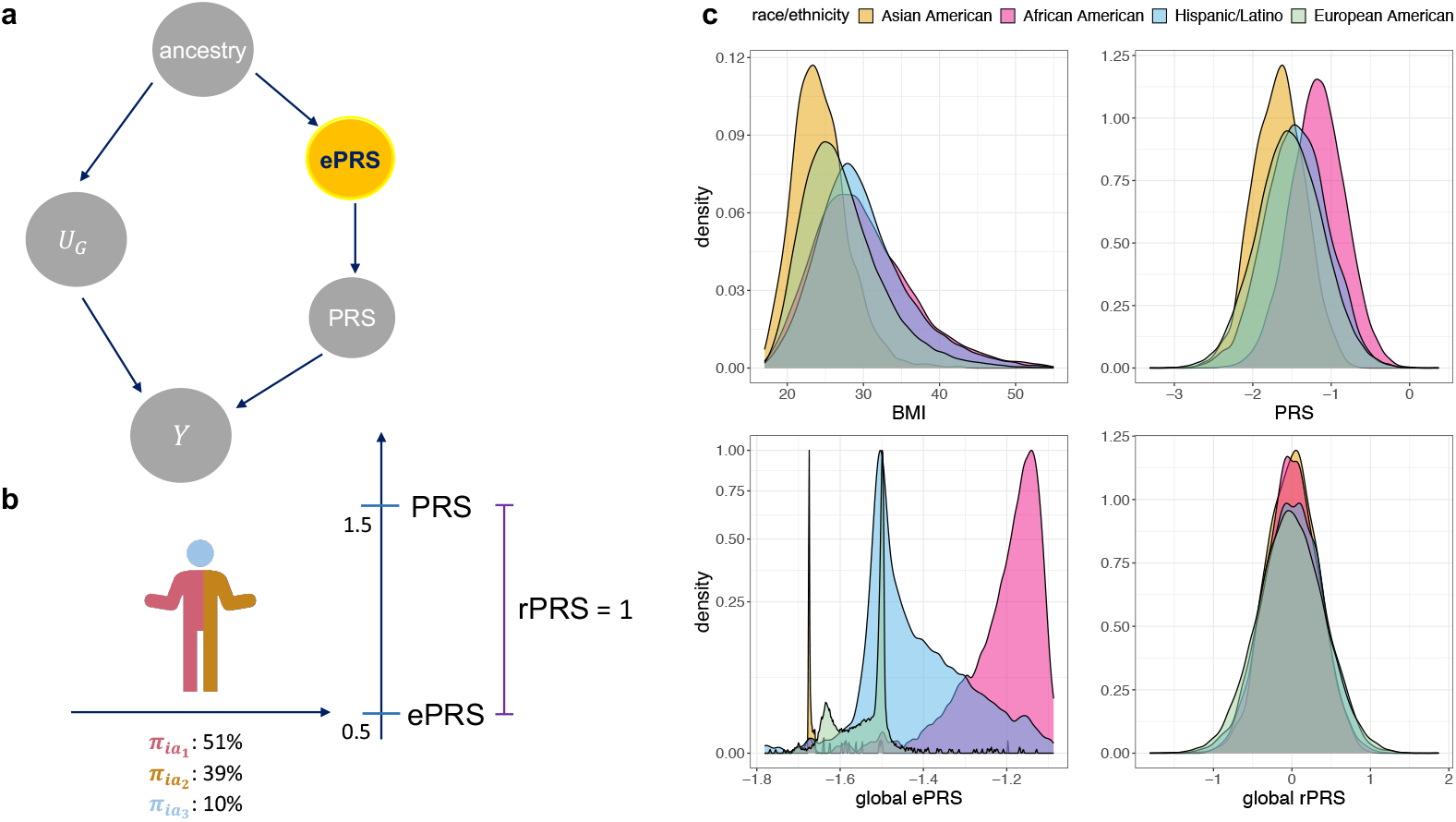
Schematic overview of the ePRS framework. Panel a: Directed acyclic graph demonstrating that the ePRS blocks a pathway between the PRS and unknown confounding genetic measures *U*_*G*_. Panel b: The rPRS is computed by subtracting the value of the ePRS from that of the PRS. The ePRS is determined according to an individual’s ancestral makeup. Panel c: The distribution of BMI and its PRS, ePRS, and rPRS in the TOPMed dataset. The distributions stratified based on harmonized selfreported race/ethnicity. Abbreviations: BMI: body mass index; PRS: polygenic risk scores; ePRS: expected polygenic risk scores; rPRS: residual polygenic risk scores; TOPMed: Trans-Omics for Precision Medicine.

To focus on the phenomena of interest (population stratification bias), in this work we assume that the SNP effect size ω_*j*_, are fixed and known values, and this work does not discuss uncertainty in their estimation nor potential differences in effects across genetic ancestries. To model the allele count of each variant and each individual, we assume that *g*_*ij*_ follows a mixture distribution based on individual *i*’s ancestry makeup:

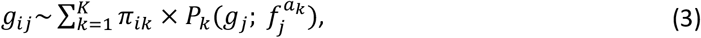

with *K* being the number of genetic ancestries 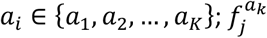 is the ancestry-specific allele frequencies of allele *j* in ancestry 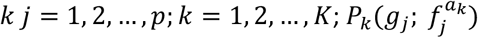 is the Binomial distribution 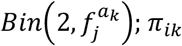 represents the proportion of the entire genome of individual *i* inherited from ancestral population *k*, i.e., the global ancestry proportion. We assume that *π*_*ik*_ are known. Note that if *π*_*ik*_ = 1 for a specific *k* ∈ {1, …, *K*}, equation (3) reduces to a standard allele count model without admixture. We can use global ancestry proportions to calculate the “global” ePRS (gePRS) as:

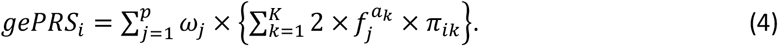

With rearrangement of terms in equation (4), for individual *i* the global ePRS can be expressed as a weighted combination of ancestry-specific ePRSs, with weights being their global ancestry proportions. If the *p* variants are independent, the variance of the PRS conditional on the global ancestry proportions is:

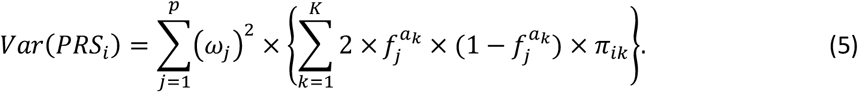

If the ancestry makeup is available in each locus (local ancestry), we can also derive local ePRS (lePRS). Assuming that each chromosomal copy’s local ancestry is known and fixed, we now use the binomial distribution with count of 1 for chromosomal copy m=1,2, i.e. 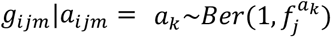. Exploiting the same strategy of deriving global ePRS, the local ePRS can be written as

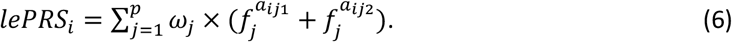

And the variance of PRS based on local ancestry is provided in equation (7):

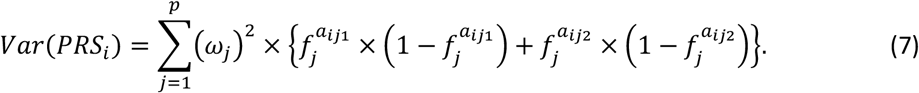

More details about the ePRS framework and derivations can be found in the Methods section.

The mean and variance of a PRS (based on either global or local ancestry patterns) can next be used to construct a third metric: the quantile PRS (qPRS). We assume that given the mean and variance, the distribution of the PRS for individual *i* follows the Normal distribution with mean *E*(*PRS*_*i*_) and variance Var(*PRS*_*i*_), i.e.,

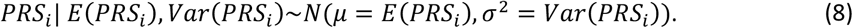

The qPRS then can be computed as

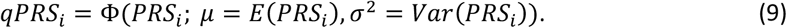

where the notation Φ is the cumulative Normal distribution function. Thus, the qPRS is a person-specific percentile of PRS value conditional on an individual’s ancestral makeup. The schematic illustration of the ePRS framework as well as example distributions of PRS, ePRS and rPRS for body mass index (BMI) in the TOPMed datasets are shown in Figure 1.

## Results from simulation studies

### Overview of the simulation settings

We conducted extensive simulation studies to examine the estimation performance of PRS-outcome association (*β*_1_ in equation (1)) when using the ePRS framework in comparison with other possible association models. We simulated a few ancestry makeup patterns and used them to generate a PRS and an unobserved confounder *U*_*G*_. Figure 2 provides an overview of the procedure for generating global and local ancestries as well as the PRS, the unknown genetic confounder *U*_*G*_, and the outcome. To guide the definition of the simulated PRS, we used summary statistics from a GWAS of systolic blood pressure (SBP) of the UK Biobank and ICBP consortium (17). We used the top 100 SNPs after clumping using PLINK and extracted their estimated effect sizes from the GWAS as the true variant weights. The ancestry-specific frequencies 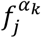 of these SNPs were taken to be the estimated ancestry-specific frequencies from the global ancestry-specific allele frequency estimation in admixed populations (GAFA) procedure applied over the TOPMed dataset (18). Two types of PRSs were considered in our simulations: homogenous weighting PRS, where the variant weights are set to be the same across ancestries, and heterogenous weighting PRS, where some of the weights are ancestry-specific (see Methods). We simulated the unknown confounder *U*_*G*_ in a few forms (Supplementary Table 1), and it was generally sampled to be ancestry-related using a similar strategy to the PRS. Further, a few forms of *U*_*G*_ were a weighted sum of alleles, just like a PRS. Thus, similar to the simulation of the main PRS, we used summary statistics from GWAS to guide the selection of SNPs to inform of ancestry-specific allele frequencies and weights. SNPs were sampled from either the UKBB-ICBP SBP GWAS (conf-PRS1) or from the SBP GWAS of the Million Veteran Program (MVP), in which case, top SNPs that were highly common (MAF > 0.4) in African ancestry and relatively rare (MAF < 0.1) in European ancestry were used (conf-PRS2). We also used a single variant or a combination of two variants as genetic confounders, again enriching for African frequency. The purpose of simulating alleles that are common in one particular ancestry was to create a strong genetic ancestry confounding effect. For comparison, we also set the “unobserved” confounder to be a known (i.e., observed) genetic principal component (conf-pc*), essentially another weighted sum of SNPs. Notably, in all simulations other than when using a conf-pc* as a confounder (for benchmarking purposes), the association model is always different than the data generating model. The PRS value considered in each association model (compare with data generating model) is the homogenous weighting PRS, which forms a model misspecification scenario when the underlying model is generated by the heterogeneous weighting PRS. We summarized the settings and the rationale for creating different simulation scenarios in Supplementary Note 1 and Supplementary Table 1.

Comprehensive details about the simulation studies are provided in the Methods section.

**Figure 2.**
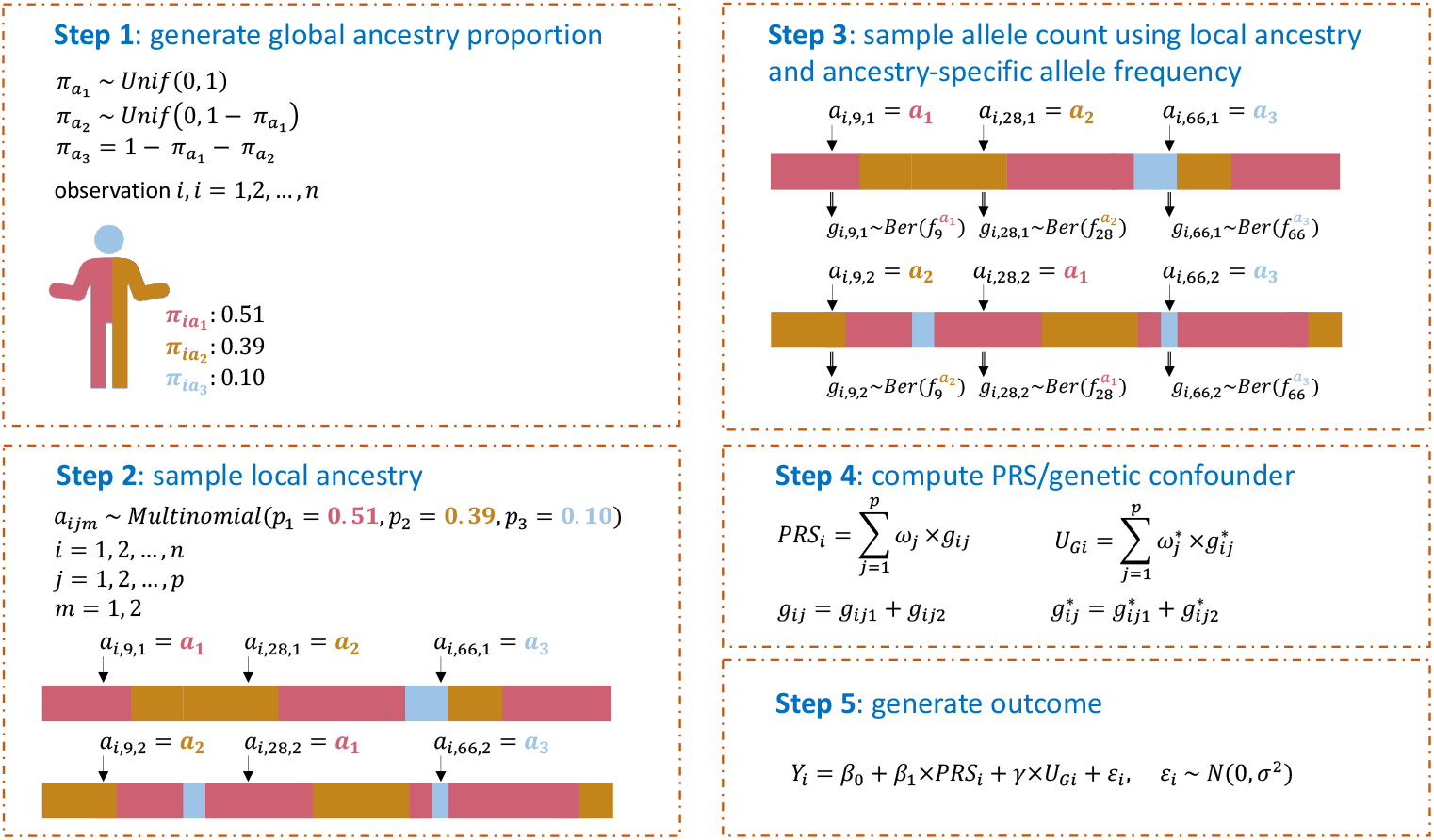
Visual illustration of the data generating mechanism in simulations. To generate data in simulations we used the 5 steps illustrated above. In step 1 we sampled global proportions of ancestry for each individual assuming 3-way admixture. Step 2 sampled the local ancestry at each SNP, on each chromosomal copy, according to the proportions of ancestry. Step 3 used the local ancestry at each SNP position and copy to sample an allele from a Bernoulli distribution (with the probability was set as the ancestry-specific allele frequency). Step 4 summed alleles with weights to obtain a PRS and genetic confounder (*U*_*G*_). Step 5 simulated the outcome from a regression model that includes an intercept, the PRS, the unobserved confounder, and normally distributed errors. Abbreviations: PRS: polygenic risk score; SNP: single nucleotide polymorphism.

### Simulation results: estimation performance of the PRS-outcome association in the ePRS framework

Selected results from simulation studies are shown in Figures 3 to 5, demonstrating that: (a) the estimated effect size of rPRS when adjusting for either global or local ePRS in the model is unbiased for *β*_1_ (Figure 3); (b) if the unknown genetic confounders share similar local ancestry intervals with the variants that make up the PRS of interest (conf-PRS1 setting), adjusted lePRS in the association model results in more efficient estimation than gePRS (Figures 3 and 4); (c) setting strong genetic ancestry confounding (conf-PRS2) causes biased estimates and dramatically increases the MSE of the estimated *β*_1_ in models that do not adjust for ePRSs (none and conf-pc*) but has nearly no increase in bias when applying the ePRS framework (Figures 3 and 4); (d) in the setting where the underlying simulated model is homogenous weighting PRS with conf-pc* confounding, the ePRS framework performed comparably to the benchmark model (conf-pc*) and outperformed other competing approaches (Figures 3 and 4); (e) compared with the approach that adjusts for PCs in the model, in which the PCs were computed based on the genetic data that generated the PRS of interest, the ePRS framework has smaller variance of *β*_1_ estimates (Figure 3) and smaller MSE (Figure 4); (f) estimation using the ePRS framework is more robust to PRS model misspecification (heterogeneous weighting PRS setting, Figures 3 and 4); (g) the efficiency in the estimation of *β*_1_ decreases (i.e., increased MSE) as the effect size *γ* of the genetic confounder, but the increase in MSE is less severe when using the ePRS framework (Figure 5). Full results from these simulations are provided in Supplementary Note 1 and Supplementary Figures 1 to 3.

**Figure 3.**
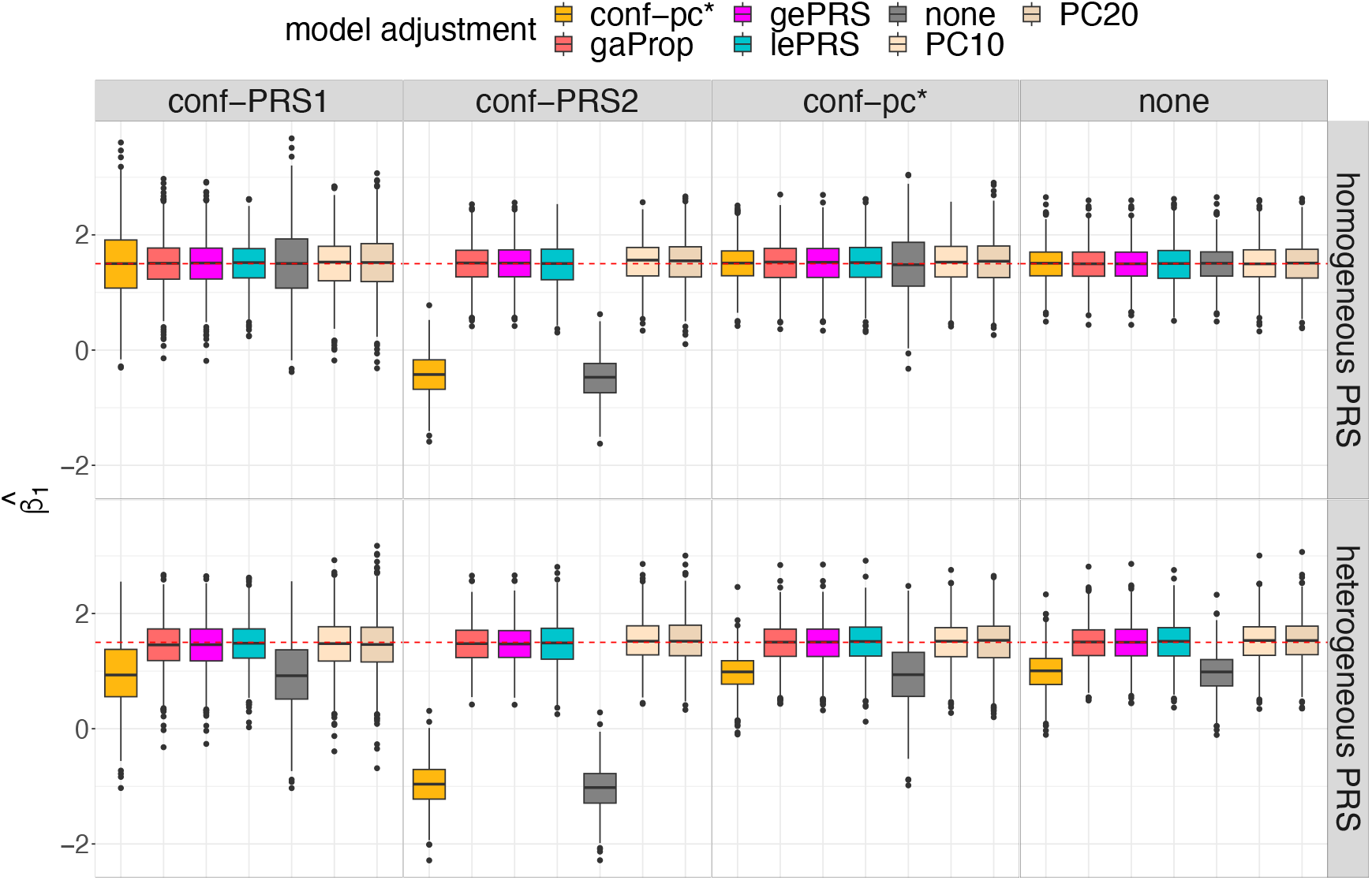
PRS effect size estimates in simulation studies. The figure provides box plots of the distribution of estimated effect sizes of the PRS 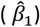 across simulation settings. Results on the top row correspond to data generating model with homogenous weighting PRS, and results in the bottom row correspond to heterogeneous weighting PRS. Columns correspond to four forms of confounding factors defined in the data generating model: conf-PRS1, conf-PRS2, conf-pc*, and no confounding (from the left to the right). Each panel provides boxplots visualizing distributions of 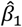 obtained in 7 association analysis models, with the y-axis representing 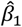 values. The true value, *β*_1_ = 1.5, is highlighted as a red dotted line. Association analyses that estimate (standard) PRS effect while adjusting to covariates include PC10, PC20 (adjusting for top 10 and 20 genetic PCs, respectively), none (no covariate adjustment), conf-pc* (adjustment for a known confounder, for benchmarking), and gaProp (adjustment for global proportions of ancestry). Association analyses applying the ePRS framework include gePRS and lePRS (estimation of rRPS effect adjusting for ePRS, based on global and local models, respectively). Distributions are provided from 1000 simulation repetitions. Abbreviations: conf: confounding; ePRS: expected PRS; gePRS: global ancestry expected PRS; lePRS: local ancestry expected PRS; PC: principal component; PRS: polygenic risk score; gaProp: global ancestry proportion.

**Figure 4.**
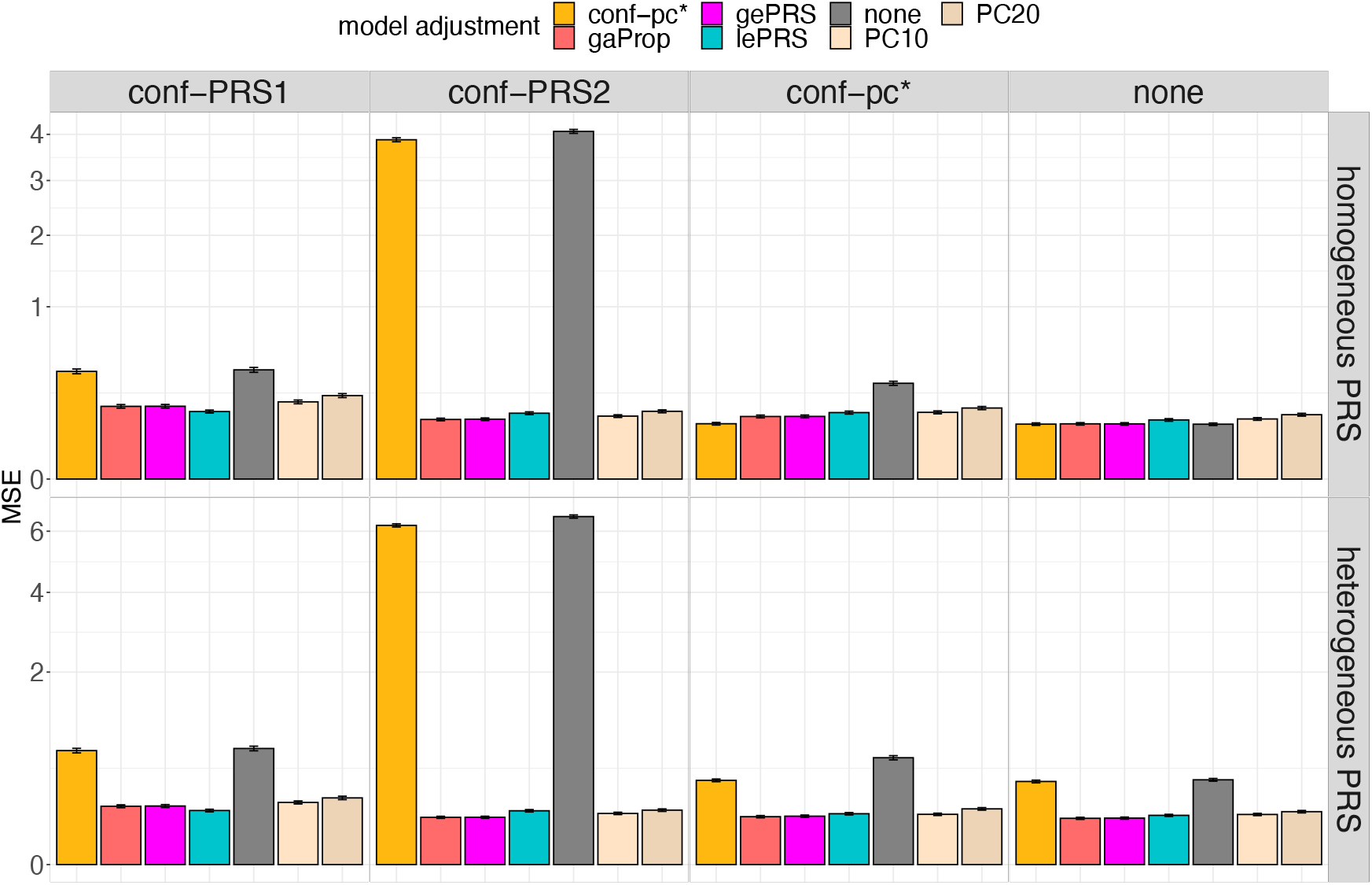
Mean squared error of PRS effect size estimates in simulation studies. The figure provides bar plots visualizing the MSE of estimated effect sizes of the PRS 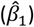 across simulation settings. Results on the top row correspond to data generating model with homogenous weighting PRS, and results in the bottom row correspond to heterogeneous weighting PRS. Columns correspond to four forms of confounding factors defined in the data generating model: conf-PRS1, conf-PRS2, conf-pc*, and no confounding (from the left to the right). Each panel provides bars with heights corresponding to the MSE of 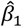 estimated in 7 association analysis models. Association analyses that estimate (standard) PRS effect while adjusting to covariates include PC10, PC20 (adjusting for top 10 and 20 genetic PCs, respectively), none (no covariate adjustment), conf-pc* (adjustment for a known confounder, for benchmarking), and gaProp (adjustment for global proportions of ancestry). Association analyses applying the ePRS framework include gePRS and lePRS (estimation of rRPS effect adjusting for ePRS, based on global and local models, respectively). MSEs were computed over 1000 simulation repetitions. Intervals around the estimated MSE correspond to the MSE +/-one estimated standard error. Abbreviations: MSE: mean square error; conf: confounding; ePRS: expected PRS; gePRS: global ancestry expected PRS; lePRS: local ancestry expected PRS; PC: principal component; PRS: polygenic risk score; gaProp: global ancestry proportion.

**Figure 5.**
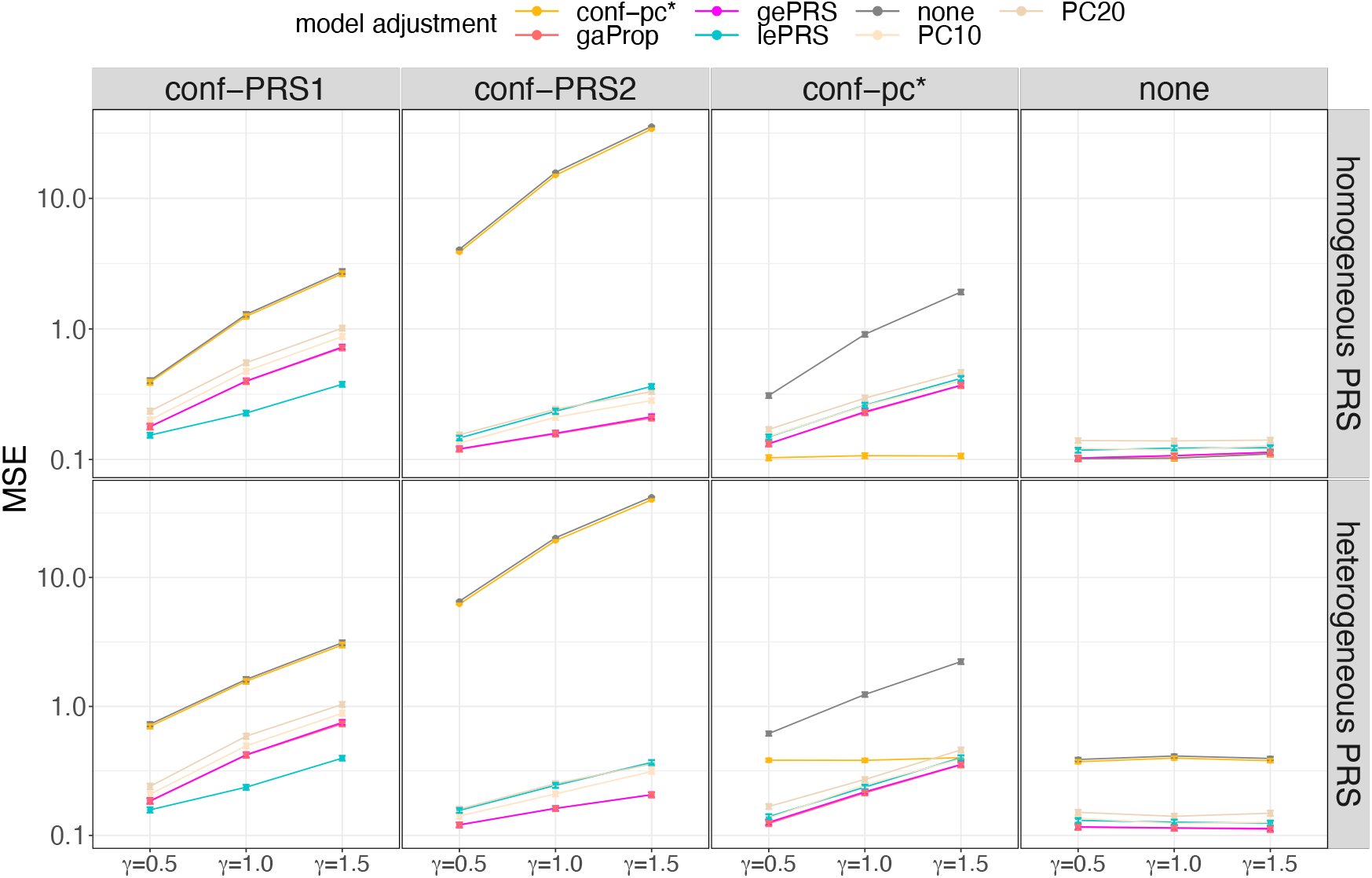
Estimation performance of the PRS effect size across increasing strength of the unknown genetic ancestry-related confounding factor. The figure provides the estimated MSE of the estimated PRS effect size 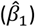 across simulation settings and analysis methods. Results on the top row correspond to data generating model with homogenous weighting PRS, and results in the bottom row correspond to heterogeneous weighting PRS. Columns correspond to four forms of genetic ancestry-related confounding factors defined in the data generating model: conf-PRS1, conf-PRS2, conf-pc*, and no confounding (from the left to the right). Each panel provides MSE (y-axis) obtained across 1000 simulation repetitions with association analyses using 7 combinations of PRS and adjustment approaches, and across 3 simulated effect sizes (*γ*) of the genetic ancestry-related confounder. In this simulation, we fixed *β*_1_ = 1.5 in the data generating model across all settings. Association analyses that estimate (standard) PRS effect while adjusting to covariates include PC10, PC20 (adjusting for top 10 and 20 genetic PCs, respectively), none (no covariate adjustment), pc* (adjustment for a known confounder, for benchmarking), and gaProp (adjustment for global proportions of ancestry). Association analyses applying the ePRS framework include gePRS and lePRS (estimation of rRPS effect adjusting for ePRS, based on global and local models, respectively). MSEs were computed over 1000 simulation repetitions. Intervals around the estimated MSE correspond to the MSE +/-one estimated standard error. Abbreviations: MSE: mean square error; conf: confounding; ePRS: expected PRS; gePRS: global ancestry expected PRS; lePRS: local ancestry expected PRS; MSA: mean squared error; PC: principal component; PRS: polygenic risk score; gaProp: global ancestry proportion.

Supplementary Note 2 provides comprehensive results from secondary simulations. Specifically, we conducted additional simulations: (a) sensitivity analyses to examine the performance of the ePRS approach under the assumption that random error exits in the global or local ancestry inference (Supplementary Figure 4); (b) simulation studies demonstrating that the estimated effect of the ePRS is related to the strength of the unknown genetic confounding effect (Supplementary Figures 5 and 6); and finally (c) simulations demonstrating the use of quantile PRS (qPRS) in binary trait risk classification analyses (Supplementary Figure 7). Each of the global and local qPRS approaches may outperform the other in some settings.

### PRS association analyses in the Trans-Omics for Precision Medicine (TOPMed) dataset

The TOPMed dataset provides aggregated whole genome sequencing (WGS) data from individuals from multiple parent studies representing genetically (and environmentally) diverse populations. We applied the ePRS framework to estimate the associations of a few PRSs with their outcomes in the TOPMed dataset. We considered five continuous phenotypes: body mass index (BMI), diastolic blood pressure (DBP), systolic blood pressure (SBP), high-density lipoprotein (HDL), low-density lipoprotein (LDL), and two binary outcomes: venous thromboembolism (VTE) and obstructive sleep apnea (OSA). Across traits, up to 49,626 individuals were included in a given analysis, with sample sizes and parent studies of participants varying across traits. Supplementary Table 2 characterizes TOPMed participants in the continuous trait analyses, Supplementary Table 3 provides summary statistics of these continuous traits in the sample. Supplementary Tables 4 and 5 characterize TOPMed participants in the VTE and OSA analyses, respectively. Supplementary Table 6 provides the number of individuals participating in each of the analyses (combined and broken down by harmonized self-reported race/ethnicity).

### Characteristics of PRS, ePRS, and rPRS across TOPMed individuals and traits

Figure 6 illustrates the impact of genetic ancestry on BMI and LDL PRSs. Panel a shows that the distribution of conventional PRS differ across self-reported race/ethnicity groups, where African American individuals have higher BMI PRS values and lower LDL PRS values, compared to other groups. This is driven by their ancestral makeup, as demonstrated by the ePRS distributions. In contrast, the rPRS distributions are similar across self-reported race/ethnicity groups and are centered around zero. Panel b provides another view of the ePRS and PRS relationship. For example, it demonstrates that the highly admixed groups, such as African American and Hispanic/Latino individuals, have large variation in ePRS values, while genomes of most Asian and European individuals have little admixture. Panel c demonstrates that while African American individuals are enriched in high BMI and low LDL PRS values, this pattern disappears when using the rPRS and the qPRS. Supplementary Figures 8-14 provide similar visualizations for other traits (SBP, DBP, HDL, OSA, and VTE).

**Figure 6.**
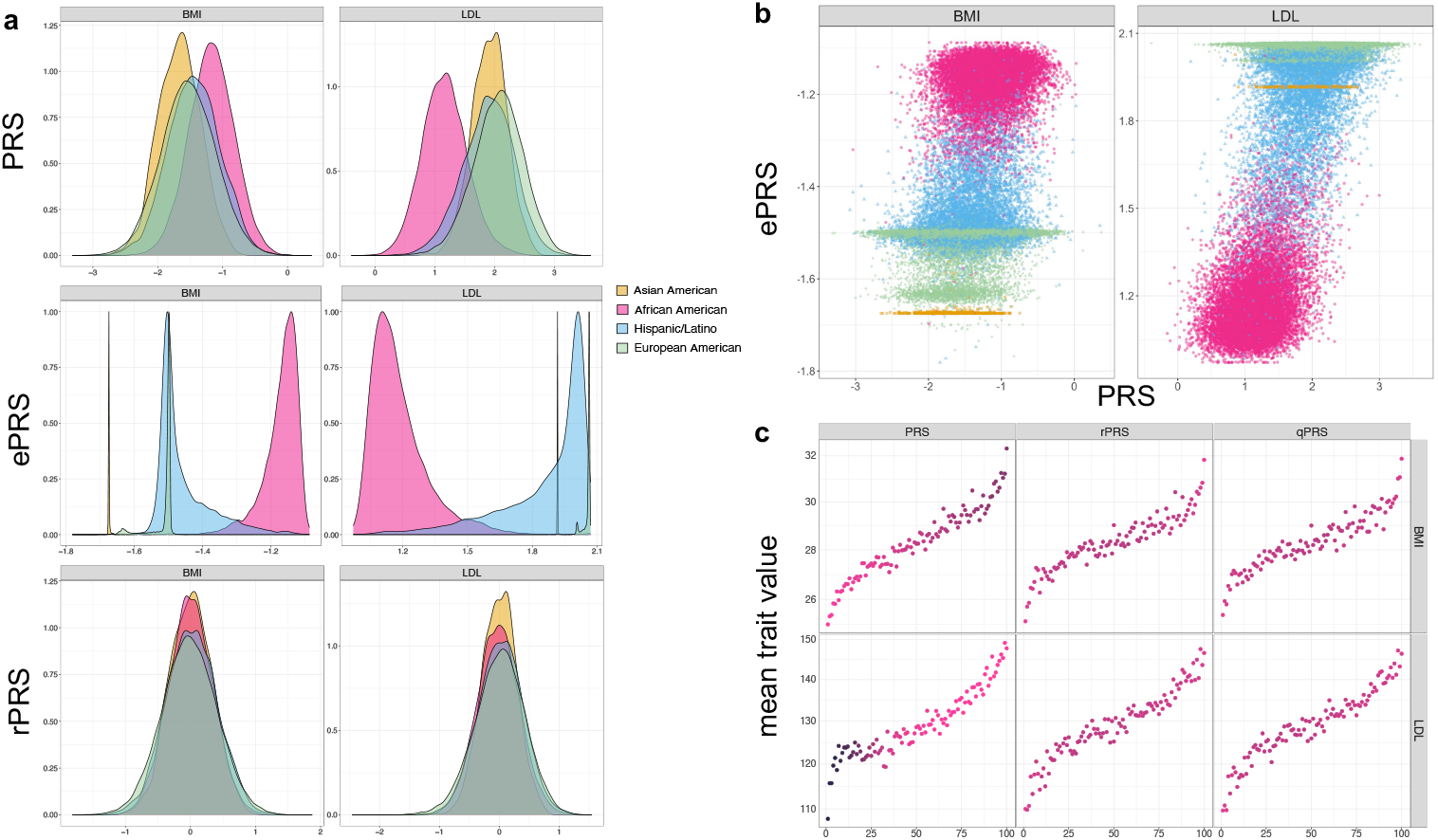
Patterns of BMI and LDL ePRS-related measures in TOPMed participants. Panel a: BMI and LDL PRS, global ePRS, and global rPRS distributions in TOPMed participants. Panel b: The relationship between BMI and LDL PRSs and global ePRSs. Each point represents an individual’s global ePRS (y-axis) and PRS (x-axis) values. Panel c: Scatterplots visualizing, on the x-axis, percentiles of PRS, global rPRS, and global qPRS, against mean values of the corresponding phenotypes (y-axis), averaged across individuals with the corresponding PRS, rPRS, or qPRS percentile. For instance, for each phenotype and PRS measure, individuals were binned into 100 strata defined by PRS measure percentiles. The color of a given point corresponds to the proportion of African American individuals among individuals in the relevant percentile stratum. Darker color reflects a higher proportion of African American individuals. Global ePRSs were constructed based on each individual’s global ancestry proportion and the ancestry-specific allele frequency estimated using GAFA. Abbreviations: TOPMed: Trans-Omics for Precision Medicine; BMI: body mass index; LDL: low-density lipoprotein; PRS: polygenic risk score; ePRS: expected PRS; rPRS: residual PRS; qPRS: quantile PRS.

### Estimation of PRS-outcome associations

We apply the ePRS framework to estimate PRS-outcome associations and compare the results to those from association models using conventional PRS and adjusting for genetic PCs or the estimated global ancestry proportions. When using the ePRS framework, we estimated the association of the rPRS while adjusting for the corresponding ePRS (global or local), computed with ancestry-specific allele frequencies calculated by GAFA.

Figure 7 shows the estimated effect sizes of the PRSs and of the rPRSs with their 95% confidence intervals. Results are provided from multi-population analysis, and stratified by selfreported race/ethnicity groups. The results from the multi-population analyses show that all four models had similar estimates, matching what we observed in simulation studies. In some cases, local ePRS analysis resulted in slightly higher estimated effect sizes (this is more apparent for DBP). The estimates obtained via multi-population analyses are close to those from the European American population, as it usually dominates other populations in sample size contribution. The estimates corresponding to the Asian American group have large standard errors due to small sample sizes. Generally, effect estimates often differ across race/ethnic groups. For example, the estimated effect sizes in the Asian American group are substantially lower than other groups for LDL and HDL traits. The estimated effect size for the African American group is higher than others for OSA (BMI adjusted) and HDL; however, the estimated effect sizes are lower than other three populations for most of the other traits. All association analysis results, including the estimated effect sizes, standard error, sample size used for each trait, are summarized in Supplementary Data 1.

**Figure 7.**
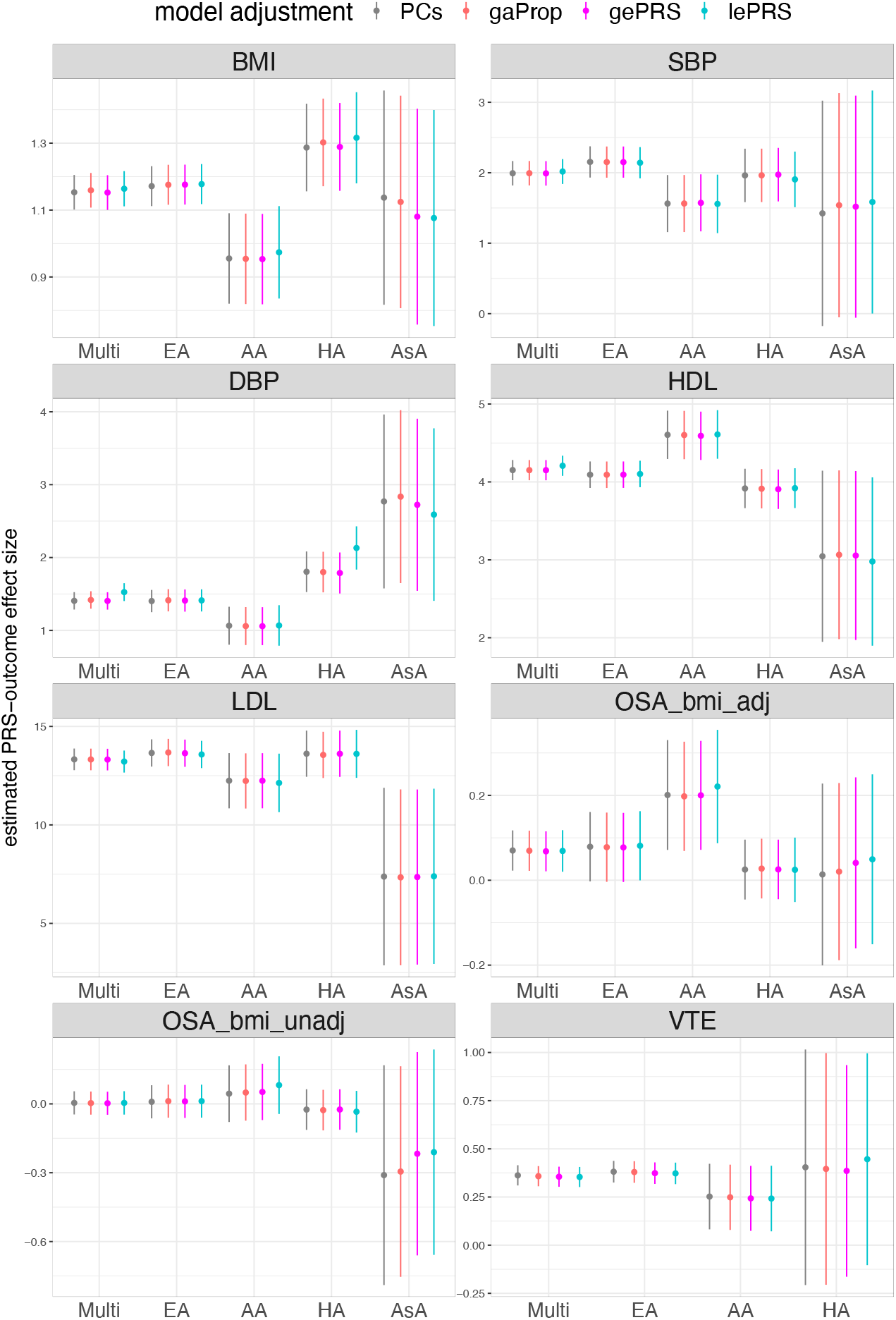
The estimated PRS-outcome effect sizes for TOPMed studies traits. Estimated PRS-outcome effect sizes (y-axis) and their corresponding 95% confidence intervals in the TOPMed dataset. Conventional PRS model estimated the PRS effect size by either adjusting for genetic PCs or for global ancestry proportions. For the ePRS model, we show the estimated rPRS effect size either adjusting for global or local ePRS. Estimated association are provided, for each trait, for the combined dataset (Multi), and stratified by self-reported race/ethnicity. For continuous traits (BMI, SBP, DBP, HDL, and LDL), effect sizes are in the original trait scale (kg/m^2^, mmHg, mmol/L). OSA and VTE are binary traits, and their estimated effect sizes are in the log odds ratio scale. For OSA, we provide results for two OSAs: OSA_bmi_adj and OSA_bmi_unadj, based on GWAS that did and did not adjust for BMI, respectively. Due to the limited sample size, we did not perform an Asian-specific analysis of VTE. More details of the analytic approaches for all the analyses can be found in the Methods section. Abbreviations: BMI: body mass index; SBP: systolic blood pressure; DBP: diastolic blood pressure; HDL: high-density lipoprotein; LDL: low-density lipoprotein; OSA: obstructive sleep apnea; VTE: venous thromboembolism; Multi: Multi-ethnic; EA: European American; AA: African American; HA: Hispanic/Latino; AsA: Asian American; PRS: polygenic risk score; PC: principal component; gaProp: global ancestry proportion; gePRS: global ancestry expected PRS; lePRS: local ancestry expected PRS.

In secondary analysis we developed genome-wide PRSs using LDpred2 (19), and constructed PRS, ePRS, and rPRS for each trait. The estimated PRS-outcome associations across all traits are similar between the ePRS framework and conventional association models (adjusting for genetic PCs or global ancestry proportions), with the same conclusion as the primary analyses. Details of secondary data analysis are summarized in Supplementary Note 4, Supplementary Figure 15, and Supplementary Data 2.

### Analysis of CVD-related traits in All of Us (AoU) research program

We applied the proposed ePRS framework to the AoU dataset using publicly-available resources. This analysis focused on six CVD-related phenotypes: atrial fibrillation (AF), coronary artery disease (CAD), chronic kidney disease (CKD), heart failure (HF), hypertension (HTN), and type 2 diabetes mellitus (T2DM), which are all binary outcomes. We used short-read wholegenome sequencing (srWGS) genetic data (version 7), restricting the analysis to variants with a population-specific allele frequency (AF) ≥ 1% or a population-specific allele count (AC) > 100. Summary statistics for PRS computation were obtained from the PGS Catalog. For computing ePRS, we used ancestry-specific allele frequencies from gnomAD (version 3.1.2), which align with the ancestry definitions in AoU. This analysis focused on computing global ePRS, which was derived using the global ancestry proportions estimated for each individual in the AoU Research Program. The characteristics of AoU participants and the selected phenotypes are summarized in Supplementary Tables 8 and 9, while details of the PGS (PGS catalog IDs, number of variants) used for PRS computation are provided in Supplementary Table 10. Additional details on the AoU analysis are available in the Methods section and Supplementary Note 5.

Figure 8 summarizes the results. We compared the estimated PRS-outcome associations obtained using the proposed approach, which adjusts for global ePRS, with the estimates derived from models either adjusting for genetic PCs or global ancestry proportions. We also included a method that uses PC-adjusted PRS (denoted as PC* in the figure) as covariate instead of PRS, following the procedure described in (4), for comparison. Overall, the PRS-outcome association estimates were consistent between the models adjusted for global ePRS and global ancestry proportions and, in most cases, were similar to those from models adjusted for PCs. However, using PC-adjusted PRS as the PRS measure, particularly for binary traits, led to underestimation of the PRS effect size estimates, as observed in the analyses of CAD and T2DM. This underestimation is because the standard deviation (SD) of the PC-adjusted PRS was sometimes very different than the SD of the PRS (prior to its regression on PCs), leading to these differences in effect sizes measured per 1 SD of the PRS measure. The characteristic of the PRSs, ePRSs, rPRSs, and qPRSs across AoU individuals and the considered traits are summarized in Supplementary Figures 16-20.

**Figure 8.**
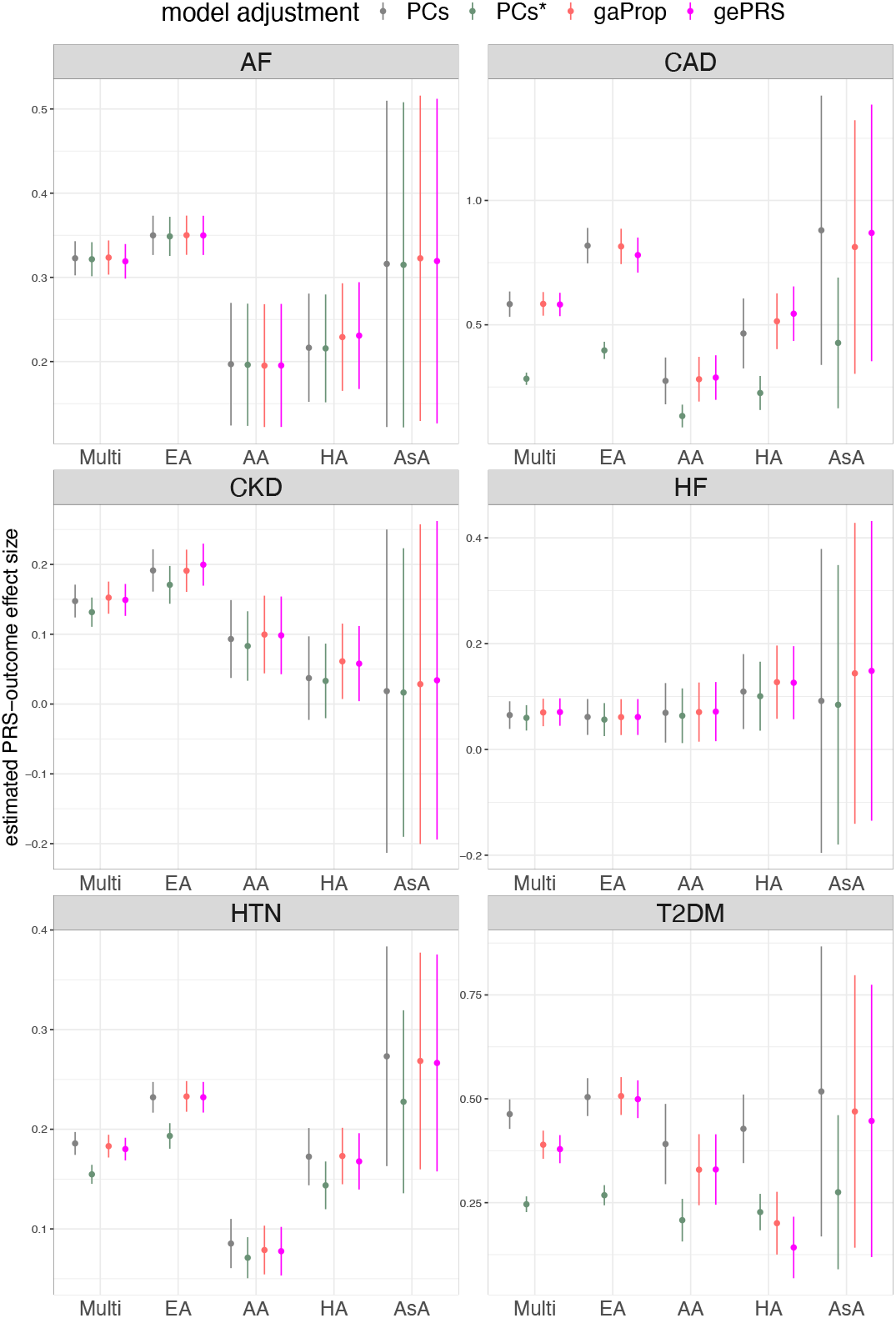
Estimated PRS-outcome effect sizes in AoU analysis. Estimated PRS-outcome effect sizes (y-axis) and their corresponding 95% confidence intervals in the AoU dataset. The conventional PRS model estimated the PRS effect size by adjusting either for genetic PCs or for global ancestry proportions. For the ePRS model, we report the estimated rPRS effect size, adjusted for global ePRS. We also include the effect size of PC-adjusted PRS (denoted as PC*), further adjusted for PCs in the association analysis, for comparison. Estimates are provided for each trait in the combined dataset (labeled “Multi”) and stratified by self-reported race/ethnicity. Effect sizes are reported on the log odds ratio scale. Further details on the analytical approaches used in all analyses can be found in the Methods section. Abbreviations: AF: atrial fibrillation; CAD: coronary artery disease; CKD: chronic kidney disease; HF: heart failure; HTN: hypertension; T2DM: type 2 diabetes mellitus; Multi: Multi-ethnic; EA: European American; AA: African American; HA: Hispanic/Latino; AsA: Asian American; PRS: polygenic risk score; PC: principal component; gaProp: global ancestry proportion; gePRS: global ancestry expected PRS.

## Discussion

We proposed an individual-level metric, the ePRS, calculated according to one’s ancestral makeup, to provide an equitable way to quantify genetic risk across diverse populations. By “equitable” we mean that an individual of a specific genetic ancestry will not be automatically annotated has “high” or “low” risk due to their ancestry, and that the same PRS would be useful independently of population descriptors such as genetic ancestry, or race and ethnicity (which are noisy correlates of genetic ancestry and have no biological meaning). By its definition, the ePRS can be considered as the baseline genetic characteristic of an individual according to their specific ancestry composition. The ePRS forms a basis for standardizing PRS across diverse and admixed individuals. We showed that adjustment for the ePRS accounts for population stratification in PRS-outcome association studies: we conducted several simulation studies and confirmed that it is sufficient to adjust for ePRS to obtain nearly unbiased estimates of the PRS-outcome associations, while adjustment for PCs (with or without the ePRS) does not improve parameter estimates. The homogeneous distribution of rPRS across different populations further demonstrates the usefulness of calibrating a PRS by the ePRS. We applied this framework to study PRS associations with 7 phenotypes in the TOPMed dataset. Across phenotypes, the estimated effect size of the rPRS adjusting for the ePRS is similar to the estimated effect of the PRS adjusting for genetic PCs or global ancestry proportion.

PRS distributions are affected by allele frequencies. As frequencies vary between groups defined by genetic ancestry patterns, PRS distributions may differ between groups, limiting the interpretation of an observed PRS value of an individual who is compared to other individuals with different genetic makeup. One of the novelties of our ePRS framework is the conceptualization of personalized PRS distribution based on one’s genetic ancestry makeup. Thus, observed PRS values are contextualized based on an individual’s ancestry composition, which determines the potential attained PRS values. Consequently, “high” or “low” PRS values are determined in relation to this potential. Individuals can be classified into at-risk groups regardless of ancestral makeup, preventing a situation where all individuals with high proportion of one genetic ancestry all appear to be at a high (or low) risk compared to individuals with high proportion of a different genetic ancestry.

Our proposed method has a few notable strengths. First, ePRS accounts for population stratification in a similar fashion to genetic PCs, however, because the ePRS is constructed such that it is specific to the PRS of interest, it has more intuitive interpretation. Further, the ePRS represents the same quantity across datasets, in contrast to PCs which are typically constructed independently in each dataset (though some approaches have been proposed for unifying PCs across datasets (e.g., (20)). While these authors also showed that PCs are used for prediction models when aligned across datasets, this alignment procedure requires joint quality control across multiple datasets, which is difficult, and critically, the ePRS framework is not inconsistent with this approach, because PCs can be incorporated to prediction models that include the ePRS. Second, the ePRS framework obviate the potential use of race and ethnicity in clinical use of PRS. While race/ethnic classification can be seen as proxies of genetic ancestry (e.g., because self-reported White individuals usually have large proportions of European genetic ancestries, etc.), two individuals self-identifying with the same race/ethnicity group may have different mixes of genetic ancestry proportions (21,22). Therefore, the distributions of PRS may arbitrarily differ across groups, potentially leading to wrong interpretations when stratifying individuals into risk groups. In contrast, genetic ancestry is defined or estimated by admixture pattern, which is a more accurate characteristic than the ambiguous definition of self-reported race ethnicity (23). How to explicitly use race/ethnicity self-reported information in biomedical or genetics research still debated, given that it is often unclear what such variables measure (22). For that reason, we expect our ePRS framework to be widely adopted and refined in order to limit potential use of self-reported race/ethnicity in clinical use of PRS while still providing accurate, well defined, measures of genetic risk.

A major consideration and limitation of the proposed ePRS framework is the computational burden and estimation accuracy of the global/local ancestry as well as the ancestry-specific allele frequency. We conducted simulation studies to examine the sensitivity of the ePRS to accuracy of both global and local ancestry inference, and showed the robustness of both global and local ePRS. Still, the performance of the ePRS framework must be influenced by the accuracy of estimated global/local ancestry pattern. In our simulation studies, although local ePRS utilizes more information than global ePRS, and, if accurate, should theoretically perform better than global ePRS, there were settings in which global ePRS outperformed the local ePRSs. Many algorithms have been proposed to conduct local ancestry inference in the past few years, including LAMP (24) and RFMix (25)(26). Among these approaches, RFMix, the approach used to estimate global/local ancestry patterns in the TOPMed dataset, showed high accuracy in estimating ancestry (27,28). We expect a more accurate and efficient algorithm to be proposed to conduct local ancestry inference and, ultimately, incorporate it into the ePRS framework to increase the estimation performance.

The ePRS is proposed to calibrate the PRS to obtain the rPRS and qPRS, which do not depend on ancestry. Another approach that have been used to “remove” the effect of ancestry on PRS is the “ancestry-adjusted PRS” method, where PCs are regressed out of the PRS via a linear model, and then the PRS-outcome association is estimated with the residuals from this regression used in lieu of the PRS (4,12,13). Khera et al. (4) showed that the ancestry-adjusted PRS had similar distribution across diverse populations, which is the same as what we found in the distribution of rPRS. However, the conventional problems of utilizing PCs, including the number of PCs to use and their generalization across datasets, are still unsolved. Moreover, local ancestry patterns specific to the PRS are not captured by standard PCs. Even though the homogenous distribution of ancestry-adjusted PRS highlights the success of adjusting PCs to account for population stratification, we think that a PRS-specific adjustment is useful. Yet, the ePRS is currently limited, compared to the PCs-adjustment method, in that it is harder to use given the computational complexity and the choices that have to be made regarding ancestry inference. Global ePRS is very easy to compute, it only requires the computation of ancestry-specific ePRS, followed by weighting according to individual’s ancestry proportion. Still, this requires choosing the level of ancestry to use (e.g., continental ancestry? more refined ancestry levels?), and inference of that ancestry, while PC-adjustment does not. Notably, PC adjustment does not account for dependency of PRS variance on admixture patterns, as it only removes mean effects. While the rPRS does not account for the variance of the PRS distribution according to genetic makeup, the qPRS does. However, currently we did not account for linkage disequilibrium between variants in the computation of the variance, and therefore only used PRSs that rely on a limited number of highly significant, independent, trait loci. It is an important extension of the ePRS framework to account for LD when computing individual PRS distribution. This will allow for computing qPRS for a PRS that is constructed based on hundreds of thousands or millions of variants. The challenge is mostly computational.

A limitation of our framework is that it uses reference populations to defined ancestry. While this is very much a standard, an important direction of current research encourages the expansion of methodology to the realm of continuous ancestry (29). When using reference populations to define ancestry, allele frequencies need to be available for the same reference populations as the ones used to quantify ancestry in the data in which ePRS is to be computed. In TOPMed, we used continental ancestries as reference populations, for local ancestry inferences, which was then averaged to provide global ancestry fractions. We computed ancestry-specific allele frequencies using the TOPMed WGS data. AoU used a different method to compute genetic ancestry fractions, and the reference populations are a bit different as Admixed Latinos are included in one category with Native American individuals (30). The AoU categories match those of gnomAD, which we used for ancestry-specific allele frequencies in ePRS computation. In all, this demonstrates that our method can be applied with existing PRS “instructions” (defined by variants and weights, independent of method used to develop them), and using available frequencies and global ancestry proportions provided by AoU. Importantly, the results in AoU are consistent with those from TOPMed, where the ePRS framework prevents population stratification bias in effect estimates in association analyses, with no need for further adjustment with genetic PCs.

In addition to clarifying the meaning of rPRS when using the ePRS, we demonstrated in our simulation studies that the estimated effect size of the ePRS is also meaningful, to some extent (Supplementary Note 2). We showed that the effect size of the ePRS can reflect the strength of unknown genetic factors based on ancestry composition. In contrast, the estimated effect sizes of PCs usually are not interpreted.

In this work, we use an individual-specific PRS distribution, assuming the variant effect sizes are fixed, admixture patterns are known, and the source of randomness is the variant frequencies. Other publications studied uncertainty in PRS quantifications by incorporating uncertainty in estimating variant weights. For example, Ding et al. (31) used a Bayesian framework to account for the potential distribution of variant weights. An exciting potential extension of this work is incorporating genetic ancestry information to simultaneously update the ancestry-specific effect size and the admixed pattern of genetics composition. It is a promising future direction for providing comprehensive PRS analysis results.

In summary, the proposed ePRS approach provides a strategy to differentiate individual genetic risk from differences in PRS distributions due to ancestral makeup, computed based on reference populations. The ePRS improves over PCs in its interpretation and transferability across datasets while protecting from population stratification bias in association analysis.

## Methods

### Computing expected PRS (ePRS)

The polygenic risk score (PRS) of an individual *i, i* = 1, 2, …, *n*, is calculated by the weighted sum of alleles, with weights being the alleles’ estimated effect sizes. This can be written as 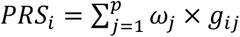. We use the clumping and thresholding method, e.g., as implemented in the PRSice software (32), in order to have a set of independent genetic variants, to enable efficient computation of the PRS variance, conditional on ancestry, as later described. Here we treat the weighting parameter *ω*_*j*_ as a fixed and known value. We define the expected PRS (ePRS) for an individual *i* as

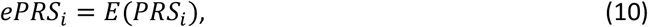

where the *E*(·) is the expectation, over the random sample of alleles. By the definition of PRS and the calculation of expectation, we can write the ePRS in the following form,

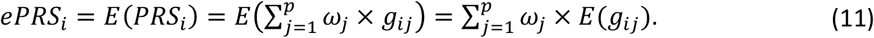

In equation (11), we assume that *g*_*ij*_ is a random variable and the estimated effect size *ω*_*j*_ is fixed value. When focusing on a homogenous population, one can assume the genetic variant follows a binomial distribution *g*_*ij*_~*Bin*(2, *f*_*j*_), where *f*_*j*_ is the allele frequency of the allele *g*_*j*_. Therefore, the expected value of the variant *j* can be computed as *E*(*g*_*ij*_) = 2 × *f*_*j*_ and the resulting ePRS is 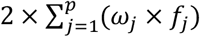for individual *i*. In addition, by assuming that the *p* variants are independent, we can apply the Binomial model to compute the variance of the PRS of individuals *i* as

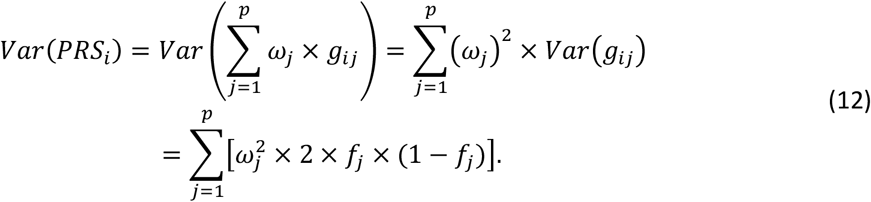

We define the residual PRS (rPRS) for individual *i* as the difference between *PRS*_*i*_ and the corresponding *ePRS*_*i*_, which is

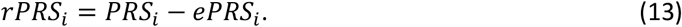

The rPRS is an index indicating the additive deviation of the PRS value from the underlying ePRS value. In a homogenous population, each individual has the same ePRS value, while they likely have different rPRS values.

Consider the “mixed population” case, in which we assume the genome for each individual is inherited from one ancestry. Assume that in the studied population there are *K* ancestries *a*_*G*_ ∈ {*a*_1_, *a*_2_, …, *a*_*k*,_}, and the allele frequency for each genetic variant *j* in ancestry *k* is 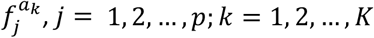. The ePRS for each individual conditioned on their ancestry can be computed as

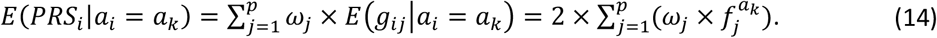

By equation (14), the ePRS for two individuals from the same ancestry is identical. Extending the idea of modeling mixed populations to admixed populations, in which the genome consists of mosaic segments inherited from different genetic ancestries, the Binomial distribution 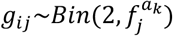 needs to be revised. In the next section, we illustrate how to calculate ePRS for admixed populations according to global and local ancestry patterns.

### Construction of ePRS using global ancestry proportions and local ancestries

#### Global ancestry ePRS

We use global ancestry proportion to calculate global ancestry ePRS (gePRS) and later extend the idea to local ancestries ePRS (lePRS) using local ancestry inference. First, we assume that for each person, the proportions of their entire genomes inherited from each ancestry is known, which are the global ancestry proportion (example is shown in Figure 1 b). We further assume that the various ancestries are uniformly distributed across the genome. Let *π*_*Gk*_ represent the proportion of the entire genome inherited from ancestry *k* for the participant *i*, where *k* = 1, 2, …., *K*. To model the genetic pattern of an admixed population, we assume that a genetic variant *j* follows a mixture model

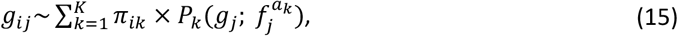

where 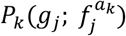 is the Binomial distribution 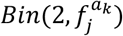 with an ancestry-specific allele frequency 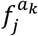. Using the mixture model (15), we can now compute the gePRS for each individual as

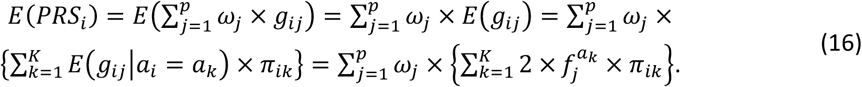

From equation (16), the gePRS can be interpreted as a weighted combination of ancestry-specific ePRSs, with weights being the global ancestry proportion. We can extend equation (12) to calculate the variance of a PRS based on equation (15). Assuming that the *p* variants are independent, the variance of the PRS conditional on global ancestry proportions is:

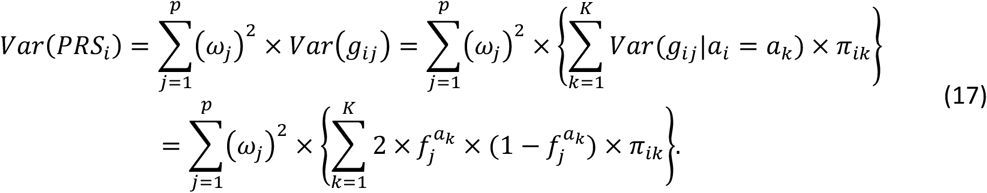

#### Local ePRS

Next, we show how to use local ancestries to construct ePRS, which we call lePRS. Suppose we know the ancestry of each locus and each chromosomal copy. Alleles for each variant are counted by the sum of two chromosomal copies, *g*_*ij*_ = *g*_*ij*1_ + *g*_*ij*2_. Assume 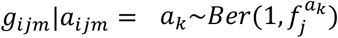, where *m* = 1, 2 denotes the two copies. The lePRS can be computed as:

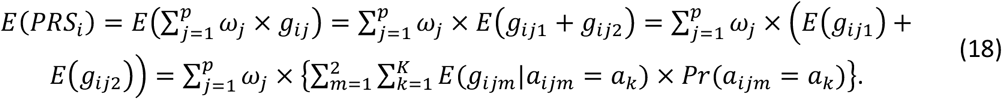

Distinct from gePRS, we assume that each copy’s local ancestry is known and fixed. Hence, by conditioning on *a*_*ijm*_ = *a*_*k*_, the last term of equation (18) can be simplified as *Pr*(*a*_*ijm*_ = *a*_*k*_) = 1 and *Pr*(*a*_*ijm*_ = *a*_*t*_; *t* ≠ *k*) = 0. Therefore, the lePRS can be expressed as followed

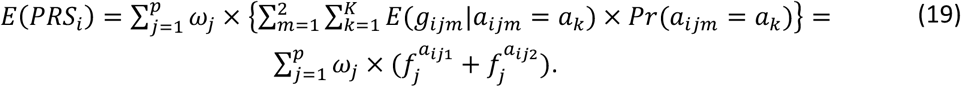

We can also compute the variance of the PRS based on local ancestry information but add additional assumptions. Here, assuming that the *p* variants are independent, and that the two chromosomal copies are also independent, the variance of the PRS can be derived as:

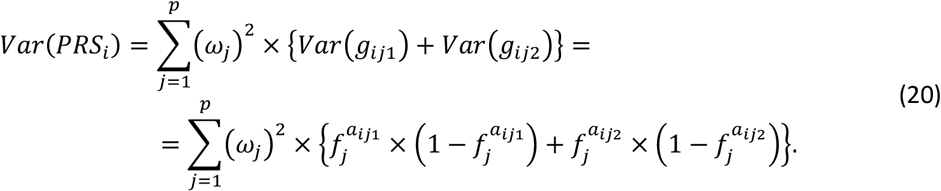

The technical considerations of computing ePRS and the related metrics are summarized in Supplementary Note 3.

#### Quantile PRS

Based on the ePRS and the PRS variance, we can construct a third metric: the quantile PRS (qPRS). We assume that given the gePRS or lePRS and the corresponding variance of PRS, the distribution of the PRS for individual *i* follows the Normal distribution with mean *E*(*PRS*_*G*_) and variance Var(*PRS*_*i*_), i.e.,

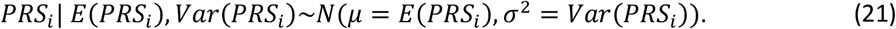

The qPRS is computed as the percentile of the PRS value conditional on an individual’s ancestral makeup, which can be written as

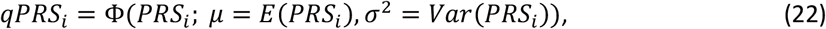

where Φ is the cumulative Normal distribution function.

### Simulations

#### Generating global ancestry proportions, local ancestries, and allele counts

Without loss of generality, we assume that the genome of each individual in the simulation study is inherited from three ancestries {*a*_1_, *a*_2_, *a*_1_}. The model could be readily extended to a higher number of ancestries. Three ancestries may represent Hispanic/Latino admixed individuals in the U.S., where, for example, *a*_1_ represents European, *a*_2_ represents African, and *a*_1_ represents American ancestry. We assume that *a*_1_ has the highest proportion across the entire genome, and generated the global ancestry proportions by sampling the three proportions of ancestry sequentially from uniform distributions, as follows:

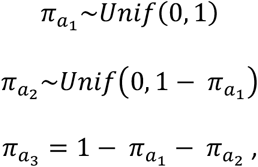

where *π*_*ak*_, *k* = 1,2,3 denotes the global ancestry proportion across the genome from ancestries *a*_*k*_ and *Unif*(*a, b*) is the uniform distribution with range (*a, b*). After generating *π*_*i*_ for each observation, we next generate local ancestry *a*_*ijm*_. Assuming that each SNP is independent, and the two chromosomal copies are also independent, we generate the local ancestry for each variant and each chromosome separately based on 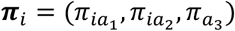. Thus, we assume that *a*_*ijm*_ follows multinomial distribution, where 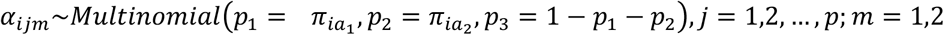. For each simulated variant, allele counts are then generated with the local ancestry guiding its ancestry-specific allele frequency.

#### Generating PRS

To select the parameters for the simulations, i.e., ancestry-specific allele frequencies and SNP effect sizes, we used summary statistics from a GWAS of systolic blood pressure (SBP), from UK biobank and the ICBP consortium (17). We preprocessed the summary statistics using PLINK with the standard parameters setting (genome-wide significance threshold (5×10^−8^), clumping parameter R^2^=0.1, and the distance was set to 1000 kb), and took 100 SNPs and their estimated effect sizes. Their ancestry-specific frequencies 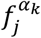 were taken to be the estimated frequencies from the GAFA procedure (18) applied over the TOPMed dataset (33). For each person and each SNP, we first sampled the local ancestry as described above, followed by sampling of allele count using the ancestry-specific allele frequency corresponding to that SNP and that local ancestry. We finally calculated the PRS as the weighted sum of SNP alleles with weights being the GWAS-estimated effect sizes. The flow chart to generate simulation data is illustrated in Figure 2.

We considered two PRS models. In the primary model, the PRS was assumed to be homogenous across genetic ancestries, i.e., the SNP effects are assumed the same regardless of ancestry, which is the homogeneous weighting PRS. The second model allowed for some heterogeneity by assuming that the SNP effects are sometimes different by ancestry. When simulating heterogeneous weighting PRS settings, we selected the top 10 most frequent SNPs in each *a*_2_ and *a*_3_ ancestries and set different effect sizes for these selected SNPs. Specifically, we set the effect sizes as 1.5 for the selected SNPs in *a*_2_ (African) ancestry and 2 for *a*_3_ (Native American) ancestry.

#### Generating the genetic ancestry confounder *U*_*G*_

We generated an ancestry-dependent genetic variable potentially confounding the PRS-outcome association. Details are provided in Supplementary Table 1. Briefly, we considered a few types of unobserved confounder, namely conf-PRS1, conf-PRS2, with computation generally following the same procedure used to generate PRS (i.e., sampling of local ancestry followed by sampling of alleles). Each confounder type was used in separate simulations. For benchmarking, we also used in simulations a variable representing a genetic PC (conf-pc*), which is observed, as a confounder.

For the first genetic confounder (conf-PRS1), we assume the generated genetic variants shared the same local ancestry interval as the variants that computed the main PRS of interest. Based on the local ancestry interval, we then used ancestry-specific allele frequencies of 100 randomly selected variants from the UKBB-ICBP SBP GWAS to generate genetic data, which is the same procedure for generating observed PRS. The only difference is that the weighting parameters of computing conf-PRS1 are generated from a standard normal distribution.

A second type of genetic confounder (conf-PRS2) is generated based on a selection of SNPs from the MVP SBP GWAS. These SNP were selected to be enriched in frequency (MAF > 0.4) in the African ancestry population while having MAF<0.1 in the European ancestral population, as estimated by GAFA. Notably, the distribution of Conf-PRS2 is quite different from that of the observed PRS, leading to strong unknown genetic confounding effect in the simulation settings.

The third continuous confounder, conf-pc*, intends to mimic a scenario where the true confounding is based on only one genetic principal component score. To guide the construction of this variable we randomly select 100 SNPs from MVP GWAS and generate the weighting parameters from standard Normal distribution. To emphasize, conf-pc* is not calculated via a principal component analysis procedure.

#### Generating outcomes via a data generating model

We generate a continuous outcome from a linear model summing the effect of the PRS of interest and the genetic confounder through the following model:

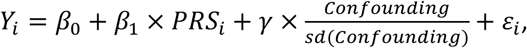

Where the genetic confounder is standardized in each simulated dataset so that effect sizes are comparable across simulation settings. We set the PRS effect *β*_1_ to 1.5 and varied the effect of the genetic confounder to be *γ* = 0.5, 1, 1.5. The intercept was set to *β*_0_ = 1 across all simulations. Finally, the errors were sampled from a Normal distribution with ε_*i*_~*N*(0,1). We sampled *N* = 10,000 observations in each simulation repetition and repeated each simulation setting 1000 times.

#### Estimation of PRS effect in simulations via a working association model

We compare the estimation performance of *β*_1_ via seven working association model, M1-M7. In all models we evaluate the estimated 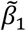 as an estimator of *β*_1_, i.e., as 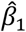. Also, all association models use homogeneous weighting PRS.

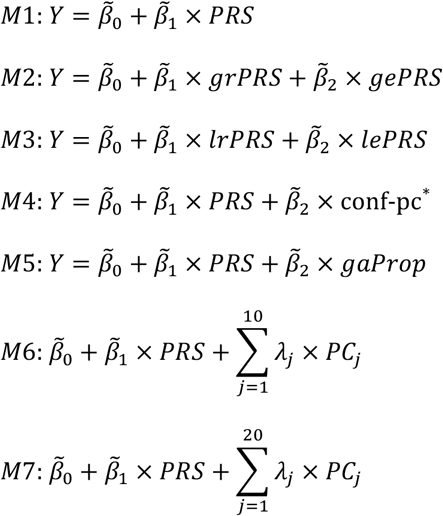

In M1, we estimate PRS association without adjusting for other covariates (we named it as “none” in our results and figures). The proposed ePRS-based approaches are provided in M2 and M3, which adjust for global and local ePRS with the corresponding rPRS. M4-M7 models again estimate the effect of the PRS, and further adjust for different ancestry-related measures. M4 uses conf-pc*, which is a benchmark when the true confounding factor is simulated as conf-pc*. M5 adjusts for the global ancestry proportion (“gaProp” in our results and Figures). M6 and M7 use 10 and 20 PCs, respectively, where the PCs were calculated based on the genetic dataset used for computing the PRS.

#### The TOPMed dataset

The TOPMed project aggregates individual-level data from multiple parent studies. Supplementary Note 6 describes TOPMed parent studies contributing to the analyses.

#### Whole-genome sequencing

We used genetic data from whole genome sequencing via the TOPMed program (34) freeze 8 released. Information about genome sequencing, allele calling, and quality control in TOPMed is publicly available in https://www.nhlbiwgs.org/topmed-whole-genome-sequencing-methods-freeze-8. The TOPMed Data Coordinating Center constructed a kinship matrix estimating recent genetic relatedness, the corresponding sparse kinship matrix, where values were set to zero when the genetic relationship was estimated to be more distant than 4^th^ degree relatedness, as well as providing genetic principal components (PCs), using the PC-Relate algorithm (35). In the TOPMed data analysis, we adjusted for 11 genetic PCs in the standard PRS model (except the VTE analysis, in which we adjusted seven genetic PCs according to the previously published paper (36)).

#### Genetic ancestry inference in TOPMed

Ancestry inference was performed by the TOPMed Informatics Research Center (IRC). First, local ancestry was inferred using RFMix (25), with default parameter settings except the following option: --node-size=5. Then, global ancestry was computed as for each participant as a weighted average of the ancestries in inferred local ancestry intervals. The reference panel used was the Human Genome Diversity Panel (HGDP) downloaded from the Stanford HGDP website http://hagsc.org/hgdp/files.html. Genomic coordinates were lifted over from genome build 37 to build 38. The 53 HGDP populations were merged into 7 super-populations: Sub-Saharan Africa, Central and South Asia, East Asia, Europe, Native America, Oceania, Middle East. Local ancestry inference was performed in two versions. First, for samples available in TOPMed freeze 6, RFMix V1 was used, and local ancestry was inferred for the autosomes only. Later, for samples participating only in freeze 8 (but not in freeze 6), and for the X-chromosome, local ancestry inference was performed using RFMix V2. Global ancestry proportions of an individual were defined as the proportions of the inferred ancestries (accounting for interval lengths) of each of the ancestries, genome-wide. Because the levels of Oceania ancestry were very low in the sample, we did not use it, and instead rescaled, for each person, the other ancestries so that they some to 1. So, if based on the RFMix global ancestry proportions we had 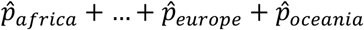 with 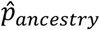 is the estimated proportion of an ancestry for an individual, we summed the non-oceania proportions for each individual to get 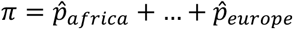. The scaled proportions were set to 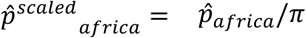 etc.

#### Computation of ancestry-specific allele frequencies

Ancestry-specific allele frequencies were computed for all common variants as we previously described (33), using the GAFA algorithm (18) that uses global genetic ancestries to deconvolute ancestry-specific allele frequencies. We also computed ancestry-specific allele frequencies using the LAFA algorithm, i.e. using local genetic ancestry patterns at each variant location, however the results are similar, so we chose to move forward with the GAFA-estimated frequencies.

#### Harmonization of self-reported race/ethnicity in TOPMed

Most of the participating studies recruited individuals from pre-defined race/ethnicity categories, which were not always the same. For example, CARDIA uses the descriptor “Black” while JHS uses “African American”. Here, we used the race/ethnicity harmonized categorized prepared by the Data Coordinating Center of TOPMed as part of a phenotype harmonization effort (37), and as descriptors we use White (non-Hispanic White or European American), Black (non-Hispanic Black or African American), Asian (Asian or Chinese American), and Hispanic/Latino. We did not use genetic ancestry to categorize race/ethnic categories, as they are not biological and reflect sociopolitical and demographic patterns, though the distribution of genetic ancestry patterns generally differs by groups defined by race and ethnicity

#### Phenotype harmonization

We used 5 continuous phenotypes, SBP, DBP, HDL, LDL, and BMI, all harmonized by the TOPMed DCC (37). To account for medication use, SBP and DBP values were increased in antihypertensive medication users by 15 and 10mmHg, respectively. For LDL, the value was adjusted via dividing by 0.7 if the individual took lipid-lowering medication.

Two binary variables used were VTE and OSA. VTE was also harmonized by the TOPMed DCC, as reported in the Seyerle et al. 2023 (36). We harmonized OSA as follows. COPDGene control participants (i.e., individuals without COPD) self-reported doctor-diagnosed OSA. Other participating studies used home sleep apnea testing device to measure the Apnea Hypopnea Index (AHI; measured in ARIC, CARDIA, CHS, and FHS as part of the sleep heart health study (38), and in MESA (39) and JHS (40)), or the Respiratory Event Index (REI; measured in HCHS/SOL (41)). For these cases we categorized OSA based on AHI/REI≥ 15 (i.e., moderat or more severe OSA). Lower AHI/REI values were categorized as no OSA.

#### Selecting SNPs and effect sizes for PRS for the studied traits

We used summary statistics from published GWAS to develop PRS (and ePRS) for each considered trait. These are reported in Supplementary Table 7, which includes information about accessing these summary statistics, the sample sizes of each GWAS population, and the number of variants used to compute PRSs. Briefly, for most traits we used summary statistics from MVP GWAS (42–45), which are multi-population GWAS. For BMI, we used the GWAS summary statistics from the Genetic Investigation of Anthropometric Traits (GIANT) consortium meta-analyzed with a UK Biobank (UKBB) BMI GWAS (46). The GIANT + UKBB study is mostly based on European ancestry. For each trait and its PRS, in primary analysis we focused on independent SNPs to allow for calculating the variance of PRS. Thus, we performed clumping using plink (47). The p-value thresholding parameter is set as the genome-wide significance threshold (5×10^−8^). The clumping parameter was set to R^2^=0.1, and the distance was set to 1000 kb. In secondary analysis we also developed PRSs and corresponding ePRSs based on the same GWAS summary statistics using LDpred2 (19). These are secondary analyses because we currently cannot compute the PRS variance when using highly correlated SNPs in the PRS. Information about these PRSs and the association analyses is provided in Supplementary Note 4.

#### PRS-outcome association analyses

PRS associations with continuous traits were estimated via linear mixed model, and with binary traits via logistic mixed models. All models used sparse kinship matrix to model a random effect to account for relatedness. Standard PRS model were adjusted for (a) 11 genetic PCs or (b) global ancestry proportions. EPRS models estimated the global or local rPRS association while adjusting for the corresponding ePRS. Other covariates were often trait-specific: BMI PRS associations were adjusted for sex, age, and squared age term, while SBP, DBP, HDL, and LDL associations were further adjusted for BMI. Logistic mixed model analysis of VTE used a similar approach to that in the published VTE GWAS (36): we adjusted sex, age, and *lnratio* (log of the ratio between the size of the case stratum and the matched control; to address the differences of case-control matched ratio). Because the model did not converge, we did not include “sample set” as a covariate. OSA analysis adjusted for sex, age, BMI, and the square of the BMI term as fixed effect covariates. All analyses were conducted via the R package *GENESIS* (version 2.32.0; (48)) with the fitNullModel function.

#### Assessment of qPRS as an equitable metric for genetic disease risk

We assessed the use of qPRSs for ranking genetic liability quantified by PRS, instead of using the conventional PRSs which are highly affected by ancestral makeup. To this end, we visualized patterns of trait values and qPRS percentiles. For each trait, we stratified individuals to 100 bins defined by values of PRS, rPRS, and qPRS (each separately), and computed the mean value of the trait or proportion of the binary outcome, and visualized these as scatterplots. To demonstrate the impact of ancestral makeup on the PRS metric ranking, we colored each point in the scatterplot by the proportion of African American individuals in the strata (or bin). We chose this population group because the sample is sufficiently large.

#### The AoU data analysis

We used short-read whole-genome sequencing (srWGS) data (version 7) from the AoU study to compute PRS and performed PRS-outcome association analyses. To minimize the memory storage requirements, we used the genomics data pre-filtered by the following criteria: population-specific allele frequency ≥ 1% or population-specific allele count > 100 (data from: gs://fc-aou-datasets-controlled/v7/wgs/short_read/snpindel/acaf_threshold_v7.1). Only unrelated individuals were included, with related individuals excluded based on information in: gs://fc-aou-datasets-controlled/v7/wgs/short_read/snpindel/aux/relatedness/relatedness_flagged_samples.tsv.

Detailed quality control (QC) procedures, including both genomic and sample QC, can be found in the Genomic Research Data Quality Report: https://support.researchallofus.org/hc/article_attachments/27634053350292.

The analysis focused on adults aged 18 to 95 with BMI values ranging from 17 to 55, consistent with the selection procedure used in the TOPMed analysis. Sample sizes for self-reported race/ethnicity groups varied slightly depending on the phenotype to analyze. Six binary CVD-related phenotypes were considered: atrial fibrillation (AF), coronary artery disease (CAD), cardiovascular disease (CVD), heart failure (HF), hypertension (HTN), and type 2 diabetes mellitus (T2DM). Selection criteria for clinical outcomes, including associated SNOMED codes and OMOP Concept IDs in the AoU study, are detailed in Supplementary Table 9 and Supplementary Note 5.

#### Genetic ancestry inference and ancestry-specific allele frequency

The global ancestry proportion for each individual with srWGS data from the AoU is available at: gs://fc-aou-datasets-controlled/v7/wgs/short_read/snpindel/aux/ ancestry/ancestry_preds.tsv. Six ancestry populations are reported: African/African American (afr), American Admixed/Latino (amr), East Asian (eas), European (Eeur), Middle Eastern (mid), and South Asian (sas). These categories align with the ancestry definitions used in gnomAD, the Human Genome Diversity Project, and the 1000 Genomes Project. Additional details on computing categorical ancestry for individuals in AoU are provided in the Genomic Quality Report (https://support.researchallofus.org/hc/article_attachments/27634053350292)

The ancestry-specific allele frequency used for computing ePRS were obtained from gnomAD (version 3.1.2). The data can be access at: gs://gcp-public-data--gnomad/release/3.1.2/ht/genomes/gnomad.genomes.v3.1.2.hgdp_1kg_subset_variant_annota tions.ht using Hail procedures. The genetic ancestry group labels from gnomAD are harmonized with those from Human Genome Diversity Project and the 1000 Genomes Project, aligning with the definitions used in AoU. Further details about genetic ancestry inference in gnomAD are available at: https://gnomad.broadinstitute.org/news/2023-11-genetic-ancestry/

#### PRS-outcome association analysis

Since only unrelated individuals were included in this analysis, logistic regression was used to conduct PRS-outcome associations. All models were adjusted for sex at birth, BMI, age, and the square term of age. For the conventional PRS model, either 16 genetic PCs or global ancestry proportions were included to account for population stratification bias. For the ePRS framework, the outcome of interest was regressed on the rPRS with the global ePRS included as an adjustment in the model. Additionally, PC-adjusted PRS were analyzed instead of using unadjusted PRS as covariates, and the 16 genetic PCs were also adjusted in the model for the comparison purposes.

## Data availability

TOPMed freeze 8 WGS data and harmonized BP and lipid phenotypes are available by application to dbGaP according to the study specific accessions: Amish: “phs000956”, ARIC: “phs001211”, CARDIA: “phs001612”, CFS: “phs000954”, CHS: “phs001368”, COPDGene: “phs000951”, FHS: “phs000974”, GENOA: “phs001345”, HCHS/SOL: “phs001395”, HVH: “phs000993”, JHS: “phs000964”, Mayo VTE: “phs001402”, MESA: “phs001211”, WHI: “phs001237”. Summary statistics from MVP GWAS are available from dbGaP by application to study accession “phs001672”. Summary statistics from GIANT + UKBB GWAS were downloaded from https://portals.broadinstitute.org/collaboration/giant/index.php/GIANT_consortium_data_files. Data needed to construct the reported PRSs in this study include variants, alleles, and weights for each of the PRS are deposited on fighsare, and will be deposited on the PGS catalog. A dataset with ancestry-specific allele frequencies computed using GAFA on the TOPMed dataset for Europe, Africa, Middle East, East Asia, South Asia, and America ancestries for HapMap3 variants, which are recommended for use by the LDpred2 software, are available on the figshare repository: https://figshare.com/articles/dataset/ePRS_project_summary_statistics_and_ancestry-specific_allele_frequency/25336294. The summary statistics and ancestry-specific allele frequency used in AoU analysis can also be found in our figshare repository. Data from the NIH AoU study are available via institutional data access for researchers who meet the criteria for access to confidential data. To register as a researcher with AoU, researchers may use the following URL and complete the laid-out steps: https://www.researchallofus.org/register/. The srWGS genomic data were available on: gs://fc-aou-datasets-controlled/v7/wgs/short_read/.

The ancestry-specific allele frequency can be downloaded from gnomAD Google Cloud Public Datasets: gs://gcp-public-data--gnomad/release/3.1.2/ht/genomes/gnomad.genomes.v3.1.2.hgdp_1kg_subset_variant_annota tions.ht.

## Code availability

R codes used for simulation studies and for constructing ePRSs are available on the GitHub repository: https://github.com/Gene-Huang/Expected_PRS

## Acknowledgements

This work is supported by National Heart, Lung, and Blood Institute (NHLBI) grant R01HL161012 and National Aging Institute grant R01AG080598 grant to TS, and National Human Genome Research Institute (NHGRI) grant R56HG013163. SZ is supported by NHGRI grant R01HG011031. NLS is supported by NHLBI grants R01HL139553 and R01HL154385. GMP is support by R01HL142711 from NHLBI. This analysis was approved by the Beth Israel Deaconess Medical Center Committee on Clinical Investigations, protocol #2023P000279, and by the Mass General Brigham IRB, protocol #2021P001928. We gratefully acknowledge All of Us participants for their contributions and also thank the National Institutes of Health’s All of Us Research Program for making available the participant data examined in this study. The All of Us Research Program is supported by the National Institutes of Health, Office of the Director: Regional Medical Centers: 1 OT2 OD026549; 1 OT2 OD026554; 1 OT2 OD026557; 1 OT2 OD026556; 1 OT2 OD026550; 1 OT2 OD 026552; 1 OT2 OD026553; 1 OT2 OD026548; 1 OT2 OD026551; 1 OT2 OD026555; IAA #: AOD 16037; Federally Qualified Health Centers: HHSN 263201600085U; Data and Research Center: 5 U2C OD023196; Biobank: 1 U24 OD023121; The Participant Center: U24 OD023176; Participant Technology Systems Center: 1 U24 OD023163; Communications and Engagement: 3 OT2 OD023205; 3 OT2 OD023206; and Community Partners: 1 OT2 OD025277; 3 OT2 OD025315; 1 OT2 OD025337; 1 OT2 OD025276. The All of Us Research Program would not be possible without the partnership of its participants.

## Author contributions

Y-JH and TS conceptualized the method. Y-JH, NK, MOG, and TS brainstormed the method development and application. Y-JH performed simulation studies. Y-JH, NK, and BWS performed data analysis related to PRS development, phenotype harmonization, and association analyses. Y-JH performed data analysis in AoU platform. JW and SZ performed local and global ancestry inference in TOPMed. AS and CL performed phenotype harmonization on behalf of TOPMed Data Coordinating Center. Y-JH and TS drafted the manuscript. PSdV, HC, Y-IM, MS, GMP, LL, XG, JCB, JAB, LMR, JAS, WZ, JIR, SSR, SR, MF, RK, NF, DL, ACM, EB, NLS, CK, and BMP contributed to design and data curation in TOPMed studies they represent. NK, MOG, BWS, JW, AS, CL, PSdV, HC, Y-IM, MS, GMP, LL, XG, JCB, JAB, LMR, JAS, WZ, JIR, SSR, SR, MF, RK, NF, DL, ACM, EB, NLS, CK, BMP, and SZ critically reviewed the manuscript.

