## Supplementary information for "The expected polygenic risk score (ePRS) framework: an equitable metric for quantifying polygenetic risk via modeling of ancestral makeup"

Supplementary Figure 19: Density plots of PC-adjusted PRSs in AoU. ... 48

Supplementary Figure 20: Associations between percentiles of PRS-related measurements and six CVD-related outcomes' proportions. ... 49

*Supplementary Note 6: Descriptions of TOPMed parent studies ... 50*

Amish ... 50

ARIC ... 50

CARDIA ... 51

CFS ... 52

CHS ... 53

COPDGene ... 55

FHS ... 55

GENOA ... 56

HCHS/SOL ... 57

HVH ... 58

JHS ... 59

Mayo VTE ... 61

MESA ... 61

WHI ... 63

*Supplementary Note 7: TOPMed and CCDG acknowledgements ... 64*

*Supplementary Note 8: TOPMed consortium investigators ... 66*

*References ... 74*

### Supplementary Note 1: Primary simulation studies

#### Overview of simulation settings

We compared the estimation performance of the PRS effect size $\beta_1$ using ePRS approaches with other alternative approaches. Our simulation studies rely on (a) a data generating model, and (b) an association analysis model (which we compare). The association model is typically different than the data generating model. For the data generating model, we considered two types of PRS settings. First, the PRS is the same regardless of ancestry, i.e., the variant weights are the same across ancestries. This is the homogeneous weighting PRS setting. Another setting has a subset of the variants where the weights differ by ancestry. We refer to this as the heterogenous weighting PRS setting. We also considered a few types of genetic ancestry-related unobserved confounding factors. Supplementary Table 1 summarizes the ways we simulated unknown genetic confounding factors and the goals (hypothesis to examine) of each simulation setting. One construction of an unknown confounding variable was PRSs, i.e. weighted sum of alleles. One such PRS shared the same local ancestry intervals with the computed observed PRS of interest (conf-PRS1), such that a given SNP used in conf-PRSs was on the same local ancestry interval as another SNP used in the PRS of interest. In another confounder setting, conf-PRS2, the local ancestry intervals used for the confounder were sampled independently of those of the primary PRS of interest. In addition, we used simplified confounder variable using only either a single or two variants that are more enriched in a specific ancestral population. For benchmarking, we also generated another continuous PRS-type variable, the conf-pc*, in which address the potential extreme scenario were all the unknown genetic founding can be captured via one principal component. In all simulations the true PRS effect size was fixed at $\beta_1 = 1.5$ while we varied the effect of the genetic confounder, to assess the impact of the strength of confounding on the estimation of $\beta_1$.

We compared 7 association analysis models with different PRS measures and covariate adjustments. All association models relied on the assumption of homogeneous weighting PRS, mimicking the accepted/desired PRS association analysis approach. The ePRS approach used global and local ePRS with the corresponding rPRS in the model. These approaches were compared to "conventional" PRS approaches, were the homogeneous PRS was used, with the following adjusting covariates (separately): (a) top 20 or top 10 genetic PCs that were calculated based on the genetic data that generated the PRS of interest; (b) each individual's global ancestry proportion; (c) the conf-pc*, which is computed as the benchmarking purpose in the conf-pc* scenario; (d) no covariate-adjusted.

#### Simulating the observed PRS

The observed PRS (i.e., the PRS of interest) was designed as a weighted sum of 100 variant alleles. We used summary statistics from real data to guide the (a) ancestry-specific allele frequencies for the simulated alleles, and (b) the weighting of the alleles. In homogeneous weighting settings, we identified the top 100 SNPs (after pruning) from the UKBB+ICBP SBP GWAS. For each allele, we identified its pre-computed ancestry-specific allele frequency (computed by applying GAFA on the TOPMed dataset), and used these frequencies to sample alleles, as described in Figure 2 in the main manuscript. Similarly, alleles were summed using the corresponding weights from the UKBB+ICBP SBP GWAS. In heterogeneous weighting settings, we set ancestry-specific weights for some of the alleles to increase potential bias/error due to population stratification. Specifically, out of the 100 variants identified from the UKBB+ICBP SBP GWAS, we identified 10 variants with the highest allele frequency in African ancestry, and the 10 variants with highest MAF in the Amerindian ancestry. We set the African-specific and Amerindian-specific effect sizes for these alleles to be 1.5 and 2, respectively. These values tend to be higher compared to the estimated effect sizes of the top 100 SNPs in the GWAS.

#### Details summary of the primary simulation results (from main manuscript)

Figure 3 (main manuscript) shows the distribution of estimated $\beta_1$ from each model under 1000 simulation replicated. The corresponding mean square error (MSE) values and their standard error are shown in Figure 4 (main manuscript). In these simulations, the true value of $\beta_1$ was set as 1.5 and the effect size of unknown confounding ($\gamma$) was set as 0.5. For homogeneous weighting PRS with conf-PRS1 scenario, all association models had similar, unbiased, $\beta_1$ estimates (on average across simulations), but the MSE of ePRS-based approaches is much smaller than other methods. Compared with the model adjusted with global ancestry proportions, which used the true simulated global ancestry proportions in model fitting, both global and local rPRS showed similar or slightly better efficiency (MSE) of parameter estimates. Notably, the local ePRS model performs better than global ePRS (in terms of smaller MSE) in the conf-PRS1 setting. This is due to the data-generating process, where each SNP in conf-PRS1 shared the same local ancestry interval as a SNP in the observed PRS. Therefore, because the local ePRS, which leverages information at the local ancestry interval, performs better than the global version, which only utilizes the aggregated ancestry information.

Considering the heterogeneous weighting PRS settings, where all models are misspecified, ePRS-based approaches and the standard adjustment of PCs approach still have nearly unbiased estimation and smaller variance compared to other models. In contrast, the model which only considers PRS without covariate-adjustment or pc* (that does not include variants used in the PRS, but does include information about ancestry in that it was generating while accounting for each individual's ancestry makeup) tend to produce biased estimates.

In the Conf-PRS2 confounding setting, in which we generated the confounding in a way that prioritized higher MAF for one of the ancestries, the ePRS-based approaches and PCs adjustment provided nearly unbiased estimation and much smaller MSE compared with others. In the confounding pc* setting (conf-pc*), where the exact confounding factor is conf-pc*, the ePRS-based methods perform similarly to the ground truth model (PRS with conf-pc* adjustment). However, if the underlying outcome is generated by the heterogeneous weighting PRS, the model with conf-pc* adjustment still resulted in biased estimation, while the ePRS framework is unbiased and have smaller MSE than PC-adjusted analyses.

To examine how sensitive each model is to the change of confounding effect size, we fix $\beta_1$ as 1.5 and consider three different values of $\gamma$ $(0.5, 1, 1.5)$ of the confounding factor. These results are presented in Figure 5 (main manuscript). Across all settings, the MSE value of each method increases when the confounding effect size gets larger. However, the increase is lower when using ePRS-based and PC-based approaches, and the ePRS-framework has lower MSE than PC-based approaches when conf-PRS1 is the confounder.

##### Results from simulations using single/two variants as confounders

We consider another two scenarios where the confounding factors are (a) a single genetic variant and (b) two genetic variants as described before, but the results are not shown in the main manuscript. The rationale of these two settings is to create relatively simple scenarios in which we could easily confirm hypotheses about effects of ancestry-specific allele frequencies on estimation. Specifically, the variant(s) were simulated to have ancestry-specific frequencies as those of variants where the African frequency ranged from 0.45 to 0.55 and lower than 0.1 in the European population (see Supplementary Table 1). The results are shown in Supplementary Figures 1, 2, and 3. Here, analysis without adjusting for any covariate ("none"), performs poorly. In contrast, the ePRS and PC-adjusted approaches still had a nearly unbiased estimation, and the ePRS approach performed better than the conventional estimation of a PRS effect while adjusting for PCs in terms of MSE.

#### Supplementary Note 2: secondary simulations studies

##### Sensitivity analysis considering random errors in ancestry inference

We conducted additional simulation studies to check the robustness of the ePRS approach by adding random estimation error to the local and global ancestries used in the association model (compared to the data generation procedure), mimicking a random error in the ancestry estimation process. For global ePRS implementation, we assumed that there are errors in the global ancestry proportions, which are used for global ePRS construction. Specifically, for each of the individuals, we identified the ancestries with the highest and lowest proportions. We reduced the "inferred" global ancestry proportion of the ancestry with the higher (true simulated) proportion by 0.1 and increased the "inferred" global ancestry proportion of the ancestry with the lowest (true) proportion by 0.1. For local ePRS implementation, we assumed that there are errors in the local ancestry inference, where the ancestry of some of the intervals is misidentified. To induce such errors, we randomly selected 30 SNPs (out of the 100 used in the simulated observed PRS) and permuted their local ancestries in all individuals. We then compare the estimated effect size of the rPRS when constructed based on the accurate underlying ancestry information and based on the one with added random errors. The results are illustrated in Supplementary Figure 4. For global rPRS, the results were not sensitive to the perturbed global ancestry information, while estimation results when using local rPRS with perturbed local ancestry information are more affected, likely because the level of error is higher.

#### Interpretation of ePRS effect size

We investigated whether the effect size of the ePRS is associated with the strength of the ancestry-related genetic effect. We used the simulation settings with a single variant unobserved confounder and estimated the effect size of the ePRS while increasing the effect size of the unobserved confounder. The results are shown in Supplementary Figure 5. The effect size estimates of the both global and local ePRSs increase in absolute value as the confounding effect size increases. The effect size of rPRS remains still unbiased, but its estimation variability increase. Thus, the effect size of the ePRS is related to the strength of the unobserved confounding effects.

In a second scenario, we compared the estimated ePRS effect size from homogeneous weighting PRS and heterogeneous weighting PRS settings— both without adding in any unknown confounding in the data-generating models. The results are shown in Supplementary Figure 6. Despite the working association model always using a PRS with the same weights across ancestries (even when the data generating model is heterogeneous PRS), the rPRS effect size estimates are nearly unbiased. However, the estimated ePRS effect indeed changes between the settings: the estimated global ePRS effect size was close to zero in the homogeneous PRS settings, but was, had effect size estimates were further from zero in the heterogeneous PRS settings. The patterns were opposite in the local and global ePRSs. We cannot confidently interpret these effect sizes from these results.

#### Assessment of the potential use of the qPRS for disease risk classification

We assessed the power of disease status classification based on qPRS and compare this approach to approaches that use standard PRS through simulations. We simulated data as follows. First, we used the following logistic model to generate the disease probability of each individual.

$$P(Y=1)=\frac{\exp\left(-0.84+3\times G+1.5\times\frac{confounding}{sd(confounding)}\right)}{1+\exp\left(-0.84+3\times G+1.5\times\frac{confounding}{sd(confounding)}\right)}$$

Next, we sampled disease status from Bernoulli distribution with this probability. As causal genetic effect $G$, we consider four PRS measures: homogeneous and heterogeneous PRS as previously defined, and qPRS, based on either global or local ancestry. We considered three confounding factors, conf-PRS1, conf-PRS2, and conf-pc* (for benchmarking), used in previous simulation studies. Note here that having the causal genetic measure $G$ being the qPRS encapsulates the assumption that it is the deviation of the distribution of genetic effects based on one's ancestral make up that is important for disease risk.

As in the other simulation studies, we compared a few working association models. Differently from previous simulations that focused on effect estimation, we here assessed classification performance and thus, for each simulation repetition, used a training dataset, with 80% of the simulated data, to fit the working association model, and evaluated classification accuracy on a separate testing dataset based on the remaining 20% of the simulated data. The working association models were the five logistic models M1-M5 below, and another two strategies which directly use qPRS values to classify individuals without fitting a logistic regression model.

$$M1: log(\frac{p(Y=1)}{1-p(Y=1)}) = \beta_o + \beta_1 \times PRS$$

$$M2: log\left(\frac{p(Y=1)}{1-p(Y=1)}\right) = \beta_o + \beta_1 \times gqPRS$$

$$M3: log\left(\frac{p(Y=1)}{1-p(Y=1)}\right) = \beta_o + \beta_1 \times gqPRS + gEPRS$$

$$M4: log\left(\frac{p(Y=1)}{1-p(Y=1)}\right) = \beta_o + \beta_1 \times lqPRS$$

$$M5: log\left(\frac{p(Y=1)}{1-p(Y=1)}\right) = \beta_o + \beta_1 \times lqPRS + lEPRS$$

The classification performance was measured by the area under the receiver operating curve (AUC) evaluated over the held-out testing dataset, based on the fitted values from the trained models.

The resulting computed AUCs across 1000 replications are shown in Supplementary Figure 7. In the scenarios using PRS in the data generating model (either homogeneous or heterogeneous weighting), all the association models perform similarly, with association models using both qPRS and ePRS (M3, M5) having slightly higher AUCs. The improvement of these models, relative to other association models, was most apparent in the conf-PRS2 confounding settings. When the underlying data-generating model used the qPRS, the qPRS-based models had higher AUCs than the standard PRS model. The AUCs of the logistic models with qPRS (M2, M4) are identical to AUCs from analyses that directly utilize qPRS (as outcome probabilities) to conduct classification. Overall, models M3 and M5 have the highest AUCs.

#### Supplementary Note 3: Technical consideration in the computation of ePRS and related metrics

##### Global ePRS

Construction of global ePRS is relatively simple, as it is a weighted sum of ancestry-specific ePRSs, with weights being an individual's estimated global ancestry proportions. To compute the ancestry-specific global ePRS, we need the expected counts, or dosages, of the variants in the PRS. This can be done relatively easily by constructing a "genetic data file" with a few "individuals" representing each of the genetic ancestries used. The genetic dosage of each individual and variant is the expected dosage, i.e., twice the allele frequency, of the modelled allele in the population (genetic ancestry) represented by the individual. The first few rows of such a file are, for example:

| Variant | Chr | Position | rsID | A1 | A2 | Person_AFR | Person_EUR | … | Person_EAS |
|---|---|---|---|---|---|---|---|---|---|
| 1 | 1 | 64931 | rs62639104 | G | A | $2 \times f_1^{afr}$ | $2 \times f_1^{eur}$ | | $2 \times f_1^{eas}$ |
| 2 | 1 | 732994 | rs138476838 | G | A | $2 \times f_2^{afr}$ | $2 \times f_2^{eur}$ | | $2 \times f_2^{eas}$ |
| 3 | 1 | 758351 | rs12238997 | A | G | $2 \times f_3^{afr}$ | $2 \times f_3^{eur}$ | | $2 \times f_3^{eas}$ |
| 4 | 1 | 758443 | rs61769351 | G | C | $2 \times f_4^{afr}$ | $2 \times f_4^{eur}$ | | $2 \times f_4^{eas}$ |

A challenge is that this file will always have complete data (no missing values of ancestry-specific allele frequencies) while an individual may have missing values (i.e. missing genotypes). Therefore, the ePRS constructed using this approach may not perfectly correspond to an individual's PRS. Using genome-wide PRS and the "average" approach for PRS construction, where the PRS (and here, ePRS) value is divided by the number of variants used in the PRS derivation, will result in ePRS approximately corresponding to the PRS as long as the proportion of missing alleles is low.

#### Local ePRS

Construction of the local ePRS requires accounting for the local ancestries of each individual at each variant. We address this using genetic files where instead of allele dosages, we store the expected dosage, computed based on individuals' local ancestry patterns. The expected dosage for one allele on one chromosomal copy for a given individual is simply the ancestry-specific allele frequency corresponding to the local ancestry at the locus. Accounting for two chromosomal copies: it is the sum of ancestry-specific allele frequencies (summing over the two ancestries across the two chromosomal copies, which could be identical) at the locus. Note that when using expected dosage at individual, single variant level, missingness patterns are likely, or could be set, to be identical between the allele counts and expected allele counts.

#### Supplementary Note 4: Secondary analysis using LDPred2 PRSs

In the secondary data analysis, we used the same GWAS summary statistics to develop PRSs using the LDPred2 algorithm (1) implemented in the bigsnpr R package (2). We used the HapMap3 SNP list provided in the package, the TOPMed data as an LD reference panel (because the GWAS are mostly multi-ethnic). We computed correlation matrices for these SNPs (excluding SNPs not passing quality control filters and with MAF<1%), and subsetted the correlation matrices as appropriate based on overlap with the trait-specific GWAS. We used the "auto" option for selecting tuning parameters in an unsupervised manner. The number of variants used to compute LDPred2 PRSs and ePRSs are summarized in the last column of Supplementary Table 7. Because LDpred2 updates the weights jointly (by incorporating LD information between variants), the variance of PRS, which was computed based on the assumption of independent variants (equation (5) and (7) in main manuscript), cannot be applied to this analysis. Therefore, this analysis computed PRS, ePRS, and rPRS, but not qPRS.

Supplementary Figure 15 shows the estimated PRS-outcome association effect sizes and their corresponding 95% confidence intervals. The full results are summarized in Supplementary Data 2. Due to the high computational cost of calculating local ePRS and the results usually similar to global ePRS in primary analyses, we only compare the results from global ePRS to the standard PRS model here. Across all analyzed traits, the estimates of PRS-outcome association are similar between the ePRS framework and the model adjusting for genetic PCs or global ancestry proportion, matching what we found in primary analyses. Compared to the primary analyses that used only genome-wide significant SNPs, LDPred2 PRS estimated association effect sizes are usually higher, other than for LDL and VTE. Some of the race/ethnic-specific patterns are different in the LDPred2 PRS analysis. For instance, compared to other populations, the African American group have lower estimated effect sizes for HDL and OSA in LDPred2 PRS analysis but higher in the primary analyses that focuses on genome-wide significant SNPs. Conversely, in LDPred2 PRS analysis, the VTE effect sizes are higher for the African American population compared to other population groups, while in primary analysis the pattern is reversed.

#### Supplementary Note 5: All of Us data analysis

We used short-read whole-genome sequencing (srWGS) data (version 7) and phenotype information, including outcomes of interest as well as covariates, from the All of Us (AoU) Research Program to (1) compute PRS and (2) perform PRS-outcome associations. We focused on six cardiovascular disease (CVD)-related outcomes, all of which are binary traits. Details of the sequencing and quality control procedures are comprehensively documented in the AoU Research Hub (https://support.researchallofus.org/hc/article_attachments/27634053350292). AoU provided genetic PCs, global ancestry proportion estimates, and also conduct relatedness analysis. The genetic PCs and global ancestry proportion information for each individual can be found in: gs://fc-aou-datasets-controlled/v7/wgs/short_read/snpindel/aux/ancestry/ancestry_preds.tsv. The relatedness metrics, including estimated kinship values, are available in gs://fc-aou-datasets-controlled/v7/wgs/short_read/snpindel/aux/relatedness/relatedness.tsv. We used the information provided in gs://fc-aou-datasets-controlled/v7/wgs/short_read/snpindel/aux/relatedness/relatedness_flagged_samples.tsv to remove related individuals in our analysis. In other words, only unrelated individuals were used for PRS computation and for further performing PRS-outcome association analysis.

##### Preprocessing of clinical variables and CVD-related outcomes in AoU

We began by loading basic clinical information (covariates) into the Jupyter notebook, including BMI, date of birth, sex assigned at birth, and race and ethnicity for all 245,394 individuals with srWGS data. Our analysis focused on six binary CVD-related outcomes: atrial fibrillation (AF), coronary artery disease (CAD), cardiovascular disease (CVD), heart failure (HF), hypertension (HTN), and type 2 diabetes mellitus (T2DM). Outcome data were retrieved using the corresponding SNOMED and OMOP Concept IDs, as detailed in Supplementary Table 9. Individuals without a diagnosis for the analyzed outcome were classified as controls. For individuals diagnosed with a CVD-related condition, we defined their age based on the first recorded diagnosis date and used the closest BMI measurement to that date for further analysis. For control individuals, age was defined as "age in 2024".

For data preprocessing, we included only individuals who self-reported their sex assigned at birth as either "Female" or "Male". The analysis was further restricted to adults aged 18 to 95 with BMI values between 17 and 55, consistent with the preprocess procedure done in TOPMed analysis. Individuals with documented deaths in the electronic health record were excluded. After preprocessing, the overall dataset included approximately 7,000 Asian, 42,000 Black, 40,000 Hispanic/Latino, and 114,000 White individuals, with slight variations in sample size depending on the specific outcome analyzed. The characteristics of the samples used in the association analysis for this study are summarized in Supplementary Table 8.

#### Computation of PRS and global ePRS

The summary statistics used for computing PRS were obtained from the PGS Catalog (https://www.pgscatalog.org/). Details of the summary statistics utilized are provided in Supplementary Table 10. Most of the summary statistics were derived from multi-ethnic GWASs. For preprocessing the srWGS data in AoU, we first load in the genetics data using Hail procedure, and the variants with a minor allele frequency (MAF) <1% in the AoU dataset were excluded. The resulting Hail table was then converted into a PLINK file (bed/bim/fam triplets, version 1.9), and the PRS were computed using PLINK --score comment.

To compute the global ePRS, two key pieces of information are required: (1) global ancestry proportions for each individual and (2) ancestry-specific allele frequencies for each selected variant. The global ancestry proportion data for individuals in the AoU study can be accessed at: gs://fc-aou-datasets-controlled/v7/wgs/short_read/snpindel/aux/ancestry/ancestry_preds.tsv. Since the ancestry categories in the AoU study directly correspond to categorical ancestry definitions used in gnomAD, the Human Genome Diversity Project (HGDP), and the 1000 Genomes Project, we used ancestry-specific allele frequencies computed in gnomAD for the ePRS calculation. These ancestry-specific allele frequencies are accessible via the Cloud Public Datasets at gs://gcp-public-data--gnomad/release/3.1.2/ht/genomes/gnomad.genomes.v3.1.2.hgdp_1kg_subset_variant_annotations.ht. Our analysis focused on variants that are mapped to both the HGDP and the 1000 Genomes Project. We used the ancestry-specific allele frequency information for those variants passed quality control (QC), which were performed within the gnomAD platform, to ensure the accuracy and reliability of the computed ePRS. We computed global ePRS by accounting for ancestry composition using the global ancestry proportions provided by the AoU study for the following six ancestries: African/African American (afr), American Admixed/Latino (amr), East Asian (eas), European (eur), Middle Eastern (mid), and South Asian (sas). The corresponding ancestry-specific allele frequencies were then extracted from gnomAD. The distributions of PRS, ePRS, and rPRS, stratified by self-reported race and ethnicity, are shown in Supplementary Figures 16, 17, and 18. The distribution of PC-adjusted PRS is presented in Supplementary Figure 19. A scatterplot comparing different percentiles of PRS-related measures with the proportion of disease outcomes is shown in Supplementary Figure 20.

#### Ethics statement

The All of Us research program was approved by a single IRB, the "All of Us IRB", which is charged with reviewing the protocol, informed consent, and other participant-facing materials for the All of Us Research Program. The IRB follows the regulations and guidance of the Office for Human Research Protections (https://www.hhs.gov/ohrp/index.html) for all studies, ensuring that the rights and welfare of research participants are overseen and protected uniformly. More information is provided online https://allofus.nih.gov/about/who-we-are/institutional-review-board-irb-of-all-of-us-research-program and in the All of Us design paper.

#### Acknowledgements

We gratefully acknowledge All of Us participants for their contributions and also thank the National Institutes of Health's All of Us Research Program for making available the participant data examined in this study. The All of Us Research Program is supported by the National Institutes of Health, Office of the Director: Regional Medical Centers: 1 OT2 OD026549; 1 OT2 OD026554; 1 OT2 OD026557; 1 OT2 OD026556; 1 OT2 OD026550; 1 OT2 OD 026552; 1 OT2 OD026553; 1 OT2 OD026548; 1 OT2 OD026551; 1 OT2 OD026555; IAA #: AOD 16037; Federally

Qualified Health Centers: HHSN 263201600085U; Data and Research Center: 5 U2C OD023196; Biobank: 1 U24 OD023121; The Participant Center: U24 OD023176; Participant Technology Systems Center: 1 U24 OD023163; Communications and Engagement: 3 OT2 OD023205; 3 OT2 OD023206; and Community Partners: 1 OT2 OD025277; 3 OT2 OD025315; 1 OT2 OD025337; 1 OT2 OD025276. The All of Us Research Program would not be possible without the partnership of its participants.

### Supplementary Tables

#### Supplementary Table 1: Simulating the unknown genetic confounding factors

| Confounding variable | Sampling of genetic variants | Weighting | Goal |
|---|---|---|---|
| **Conf-PRS1** | We used ancestry-specific allele frequencies of 100 randomly-selected variants from the UKBB-ICBP SBP GWAS. | Allele weights sampled from a $N(0,1)$ distribution. | Assess and compare the estimation performance of PRS-outcome association between local ePRS and global ePRS. |
| **Conf-PRS2** | We used ancestry-specific allele frequencies of the top 100 SNPs (after pruning) identified in the MVP SBP GWAS, further requiring that the variant have African-specific MAF>0.4 and European-specific MAF<0.1. | Allele weights taken from the MVP GWAS (corresponding to the SNPs selected to guide the sampling of alleles). | Generate stronger confounding compared to conf-PRS1; a reasonable scenario whether under-represented genetic ancestry (e.g., African ancestry) has unknown genetic effects from variants that are rare in other ancestries. |
| **Single variant** | We used ancestry-specific allele frequencies based on a randomly-selected variant from the top 100 SNPs described in Conf-PRS2. | No weighting. | Assess the effect of strong unknown genetic effect that is easily described. |
| **Two variants** | We used ancestry-specific allele frequencies based on a two randomly-selected variants from the top 100 SNPs described in Conf-PRS2. | No weighting (two separate variables). | Assess the effect of strong unknown genetic effect that is easily described. |
| **Conf-pc*** | We used ancestry-specific allele frequencies of 100 randomly-selected variants from the MVP SBP GWAS. | Allele weights sampled from a $N(0,1)$ distribution. | Benchmarking. |
| **None** | -- | -- | Compare model misspecification when using homogeneous PRS when the underlying PRS is heterogeneous. |

The goal (hypothesis to examine) of each simulation settings are listed in the last column of Supplementary Table 1

#### Supplementary Table 2: Characteristics of TOPMed participants used in PRS-outcome associations analyses of continuous phenotypes

| | **African American (N=12,010)** | **Asian American (N=799)** | **European American (N=27,824)** | **Hispanic/Latino (N=8,993)** | **Overall (N=49,626)** |
|---|---|---|---|---|---|
| **Sex** | | | | | |
| Female | 7,622 (63.5%) | 498 (62.3%) | 19,027 (68.4%) | 5,284 (58.8%) | 32,431 (65.4%) |
| Male | 4,388 (36.5%) | 301 (37.7%) | 8,797 (31.6%) | 3,709 (41.2%) | 17,195 (34.6%) |
| **Age** | | | | | |
| Mean (SD) | 53.7 (15.0) | 62.5 (9.79) | 57.4 (14.8) | 48.8 (14.3) | 55.0 (15.1) |
| Median [Min, Max] | 55.0 [18.0, 93.1] | 63.0 [44.0, 84.0] | 60.0 [18.0, 98.0] | 50.0 [18.0, 86.0] | 57.0 [18.0, 98.0] |
| **Current smoke** | | | | | |
| No | 8,544 (71.1%) | 759 (95.0%) | 21,948 (78.9%) | 7,050 (78.4%) | 38,301 (77.2%) |
| Yes | 3,402 (28.3%) | 40 (5.0%) | 5,788 (20.8%) | 1,937 (21.5%) | 11,167 (22.5%) |
| Missing | 64 (0.5%) | 0 (0%) | 88 (0.3%) | 6 (0.1%) | 158 (0.3%) |
| **Hypertension** | | | | | |
| No | 3,838 (32.0%) | 340 (42.6%) | 12,314 (44.3%) | 4,458 (49.6%) | 20,950 (42.2%) |
| Yes | 8,138 (67.8%) | 459 (57.4%) | 15,478 (55.6%) | 4,420 (49.1%) | 28,495 (57.4%) |
| Missing | 34 (0.3%) | 0 (0%) | 32 (0.1%) | 115 (1.3%) | 181 (0.4%) |
| **Study** | | | | | |
| Amish | 0 (0%) | 0 (0%) | 1,102 (4.0%) | 0 (0%) | 1,102 (2.2%) |
| ARIC | 1,321 (11.0%) | 0 (0%) | 5,988 (21.5%) | 0 (0%) | 7,309 (14.7%) |
| CARDIA | 1,373 (11.4%) | 0 (0%) | 1,688 (6.1%) | 0 (0%) | 3,061 (6.2%) |
| CFS | 311 (2.6%) | 0 (0%) | 244 (0.9%) | 1 (0.0%) | 556 (1.1%) |
| CHS | 696 (5.8%) | 3 (0.4%) | 2,744 (9.9%) | 38 (0.4%) | 3,481 (7.0%) |
| COPDGene | 1,423 (11.8%) | 0 (0%) | 2,191 (7.9%) | 0 (0%) | 3,614 (7.3%) |
| FHS | 3 (0.0%) | 0 (0%) | 3,079 (11.1%) | 11 (0.1%) | 3,093 (6.2%) |
| GENOA | 1,070 (8.9%) | 0 (0%) | 0 (0%) | 0 (0%) | 1,070 (2.2%) |
| HCHS/SOL | 0 (0%) | 0 (0%) | 0 (0%) | 7,618 (84.7%) | 7,618 (15.4%) |
| JHS | 3,295 (27.4%) | 0 (0%) | 0 (0%) | 0 (0%) | 3,295 (6.6%) |
| MESA | 1,100 (9.2%) | 595 (74.5%) | 1,856 (6.7%) | 1,022 (11.4%) | 4,573 (9.2%) |
| WHI | 1,418 (11.8%) | 201 (25.2%) | 8,932 (32.1%) | 303 (3.4%) | 10,854 (21.9%) |

#### Supplementary Table 3: Summary statistics characterizing continuous traits

| | African American (N=12,010) | Asian American (N=799) | European American (N=27,824) | Hispanic/Latino (N=8,993) | Overall (N=49,626) |
|---|---|---|---|---|---|
| **SBP** | | | | | |
| Mean (SD) | 135 (22.9) | 131 (24.7) | 129 (21.8) | 127 (21.2) | 130 (22.2) |
| Median [Min, Max] | 132 [73.0, 258] | 129 [77.5, 219] | 126 [61.0, 236] | 124 [77.0, 246] | 127 [61.0, 258] |
| Missing | 38 (0.3%) | 0 (0%) | 35 (0.1%) | 115 (1.3%) | 188 (0.4%) |
| **DBP** | | | | | |
| Mean (SD) | 80.4 (12.2) | 76.8 (12.3) | 75.8 (11.5) | 76.1 (12.1) | 77.0 (12.0) |
| Median [Min, Max] | 80.0 [0, 144] | 76.0 [43.5, 118] | 75.0 [8.00, 139] | 75.0 [0, 135] | 76.5 [0, 144] |
| Missing | 39 (0.3%) | 0 (0%) | 42 (0.2%) | 115 (1.3%) | 196 (0.4%) |
| **HDL** | | | | | |
| Mean (SD) | 53.4 (15.5) | 51.2 (13.8) | 53.1 (15.8) | 49.2 (13.3) | 52.3 (15.3) |
| Median [Min, Max] | 51.0 [15.4, 194] | 49.0 [23.0, 138] | 51.0 [9.63, 156] | 47.0 [13.0, 153] | 50.0 [9.63, 194] |
| Missing | 1675 (13.9%) | 50 (6.3%) | 4913 (17.7%) | 20 (0.2%) | 6658 (13.4%) |
| **LDL** | | | | | |
| Mean (SD) | 134 (44.0) | 124 (32.5) | 138 (41.0) | 130 (41.2) | 135 (41.7) |
| Median [Min, Max] | 130 [4.40, 505] | 121 [45.2, 261] | 134 [13.8, 505] | 125 [23.8, 431] | 131 [4.40, 505] |
| Missing | 3413 (28.4%) | 73 (9.1%) | 7339 (26.4%) | 301 (3.3%) | 11126 (22.4%) |
| **BMI** | | | | | |
| Mean (SD) | 30.0 (6.47) | 24.4 (3.55) | 27.2 (5.28) | 29.9 (6.02) | 28.3 (5.88) |
| Median [Min, Max] | 29.1 [17.0, 54.9] | 23.9 [17.2, 42.3] | 26.4 [17.0, 54.9] | 29.0 [17.1, 55.0] | 27.4 [17.0, 55.0] |

Abbreviation: SBP: systolic blood pressure; DBP: diastolic blood pressure; HDL: high-density lipoprotein; LDL: low-density lipoprotein; BMI: body mass index

#### Supplementary Table 4: Characteristics of TOPMed participants used in PRS-outcome associations analyses of venous thromboembolism (VTE) dataset

| | African American (N=1,551) | Asian American (N=21) | European American (N=9,296) | Hispanic/ Latino (N=126) | other (N=68) | Overall (N=11,062) |
|---|---|---|---|---|---|---|
| **Sex** | | | | | | |
| Female | 1,197 (77.2%) | 20 (95.2%) | 5,869 (63.1%) | 116 (92.1%) | 36 (52.9%) | 7,238 (65.4%) |
| Male | 354 (22.8%) | 1 (4.8%) | 3,427 (36.9%) | 10 (7.9%) | 32 (47.1%) | 3,824 (34.6%) |
| **age** | | | | | | |
| Mean (SD) | 73.2 (9.65) | 68.5 (10.6) | 66.3 (15.5) | 72.6 (10.7) | 47.4 (16.2) | 67.2 (15.0) |
| Median [Min, Max] | 74.3 [23.0, 92.9] | 64.9 [44.0, 84.4] | 69.8 [18.0, 96.8] | 74.4 [20.0, 90.9] | 47.5 [18.0, 82.0] | 70.7 [18.0, 96.8] |
| **Study** | | | | | | |
| ARIC | 716 (46.2%) | 0 (0%) | 4,160 (44.8%) | 0 (0%) | 0 (0%) | 4,876 (44.1%) |
| CHS | 112 (7.2%) | 1 (4.8%) | 277 (3.0%) | 2 (1.6%) | 0 (0%) | 392 (3.5%) |
| FHS | 1 (0.1%) | 0 (0%) | 574 (6.2%) | 2 (1.6%) | 0 (0%) | 577 (5.2%) |
| HVH | 25 (1.6%) | 2 (9.5%) | 560 (6.0%) | 10 (7.9%) | 7 (10.3%) | 604 (5.5%) |
| Mayo VTE | 3 (0.2%) | 0 (0%) | 1,254 (13.5%) | 3 (2.4%) | 61 (89.7%) | 1,321 (11.9%) |
| WHI | 694 (44.7%) | 18 (85.7%) | 2,471 (26.6%) | 109 (86.5%) | 0 (0%) | 3,292 (29.8%) |
| **VTE status** | | | | | | |
| Matched control | 1,189 (76.7%) | 15 (71.4%) | 6,093 (65.5%) | 85 (67.5%) | 0 (0%) | 7,382 (66.7%) |
| Case | 362 (23.3%) | 6 (28.6%) | 3,203 (34.5%) | 41 (32.5%) | 68 (100%) | 3,680 (33.3%) |

Abbreviation: VTE: venous thromboembolism

#### Supplementary Table 5: Characteristics of TOPMed participants used in PRS-outcome associations analyses of obstructive sleep apnea (OSA) dataset

| | African American (N=3,298) | Asian American (N=243) | European American (N=7,066) | Hispanic/Latino (N=8,153) | Overall (N=18,760) |
|---|---|---|---|---|---|
| **Sex** | | | | | |
| Female | 1,683 (51.0%) | 119 (49.0%) | 3,712 (52.5%) | 4,712 (57.8%) | 10,226 (54.5%) |
| Male | 1,615 (49.0%) | 124 (51.0%) | 3,350 (47.4%) | 3,441 (42.2%) | 8,530 (45.5%) |
| Missing | 0 (0%) | 0 (0%) | 4 (0.1%) | 0 (0%) | 4 (0.0%) |
| **Age** | | | | | |
| Mean (SD) | 57.7 (12.8) | 67.8 (9.06) | 62.2 (11.0) | 48.0 (14.7) | 55.3 (14.6) |
| Median [Min, Max] | 55.3 [18.1, 92.0] | 67.0 [54.0, 93.0] | 62.0 [18.0, 93.0] | 49.0 [18.0, 93.0] | 56.0 [18.0, 93.0] |
| **BMI** | | | | | |
| Mean (SD) | 30.3 (6.41) | 24.3 (3.20) | 28.5 (5.41) | 30.0 (6.09) | 29.4 (5.95) |
| Median [Min, Max] | 29.4 [17.1, 54.8] | 24.1 [17.1, 37.9] | 27.7 [17.1, 54.0] | 29.1 [17.1, 55.0] | 28.6 [17.1, 55.0] |
| **Study** | | | | | |
| ARIC | 0 (0%) | 0 (0%) | 1,105 (15.6%) | 0 (0%) | 1,105 (5.9%) |
| CFS | 403 (12.2%) | 0 (0%) | 410 (5.8%) | 0 (0%) | 813 (4.3%) |
| CHS | 200 (6.1%) | 0 (0%) | 688 (9.7%) | 0 (0%) | 888 (4.7%) |
| COPDGene | 1,561 (47.3%) | 0 (0%) | 2,459 (34.8%) | 0 (0%) | 4,020 (21.4%) |
| FHS | 0 (0%) | 0 (0%) | 1630 (23.1%) | 1 (0.0%) | 1,631 (8.7%) |
| HCHS | 0 (0%) | 0 (0%) | 0 (0%) | 7,663 (94.0%) | 7,663 (40.8%) |
| JHS | 609 (18.5%) | 0 (0%) | 0 (0%) | 0 (0%) | 609 (3.2%) |
| MESA | 525 (15.9%) | 243 (100%) | 774 (11.0%) | 489 (6.0%) | 2,031 (10.8%) |
| **OSA status** | | | | | |
| Control | 2,425 (73.5%) | 114 (46.9%) | 4,277 (60.5%) | 6,088 (74.7%) | 12,904 (68.8%) |
| Case | 798 (24.2%) | 114 (46.9%) | 1,568 (22.2%) | 1,050 (12.9%) | 3,530 (18.8%) |
| Missing | 75 (2.3%) | 15 (6.2%) | 1,221 (17.3%) | 1,015 (12.4%) | 2,326 (12.4%) |

Notes: In studies with overnight sleep studies, OSA was defined based on the Apnea Hypopnea Index (AHI) or the Respiratory Event Index (REI), requiring AHI/REI≥15. In COPDGene it was defined using self-reported doctor diagnosis (COPDGene).
Abbreviation: OSA: obstructive sleep apnea

#### Supplementary Table 6: Sample sizes from analyzes of each trait (primary analysis)

| Trait | African American | Asian American | European American | Hispanic/Latino |
|---|---|---|---|---|
| SBP | 11,944 | 787 | 27,787 | 8,866 |
| DBP | 11,943 | 787 | 27,780 | 8,866 |
| HDL | 10,323 | 738 | 22,909 | 8,961 |
| LDL | 8,585 | 715 | 20,483 | 8,680 |
| BMI | 11,982 | 787 | 27,822 | 8,981 |
| VTE | 1,520 | X | 7,482 | 113 |
| OSA | 3,222 | 228 | 5,835 | 7,137 |

Note: For VTE, due to the sample sizes being too small in Asian-Americans (N = 21), we did not perform Asian-American-specific analysis.
Abbreviations: SBP: systolic blood pressure; DBP: diastolic blood pressure; HDL: high-density lipoprotein; LDL: low-density lipoprotein; BMI: body mass index; VTE: venous thromboembolism; OSA: obstructive sleep apnea.

#### Supplementary Table 7: Characteristics of trait GWASs and constructed PRSs

| Trait | GWAS summary statistics | PIMD reference | Sample sizes | Study population of GWAS | Number of SNPs used (primary analysis) | Number of SNPs used (secondary analysis) |
|---|---|---|---|---|---|---|
| SBP | MVP | 30578418 | N = 318,492 | Multi-ethnic | 107 | 574,384 |
| DBP | MVP | 30578418 | N = 318,891 | Multi-ethnic | 98 | 574,294 |
| HDL | MVP | 30275531 | N = 297,626 | Multi-ethnic | 482 | 524,956 |
| LDL | MVP | 30275531 | N = 297,626 | Multi-ethnic | 245 | 521,113 |
| BMI | GIANT+UKBB | 30124842 | N ~ 700,000 | European ancestry | 1,440 | 434,712 |
| VTE | MVP | 31676865 | N = 263,795 (11,844 cases; 251,951 controls) | Multi-ethnic | 46 | 516,802 |
| BMI-unadj OSA | MVP | 36989840 | N = 568,576 (121,332 cases; 447,244 controls) | Multi-ethnic | 17 | 307,286 |
| BMI-adj OSA | MVP | 36989840 | N = 559,070 | Multi-ethnic | 6 | 323,621 |

SBP and DBP: 69.1% non-Hispanic white, 18.8% non-Hispanic black, 6.7% Hispanic, 0.77% non-Hispanic Asian, and 0.85% non-Hispanic Native American (0.85%). HDL and LDL: 72.4% non-Hispanic white, 19.3% non-Hispanic black, 8.3% Hispanic. VTE: 72.1% non-Hispanic white, 19.6% non-Hispanic black, 8.3% Hispanic. OSA: 72% non-Hispanic white, 19% non-Hispanic black, 8% Hispanic.
Abbreviations: MVP: Million veteran program; GIANT: Genetic Investigation of Anthropometric Traits; SBP: systolic blood pressure; DBP: diastolic blood pressure; HDL: high-density lipoprotein; LDL: low-density lipoprotein; BMI: body mass index; VTE: venous thromboembolism; OSA: obstructive sleep apnea; BMI-unadj OSA: OSA GWAS without BMI adjusted; BMI-adj OSA: OSA GWAS with BMI adjusted

#### Supplementary Table 8: Characteristics of AoU participants used in PRS-outcome associations analyses

| AF | Asian (N=6,933) | Black (N=42,467) | Hispanic/Latino (N=40,595) | White (N=114,021) |
|---|---|---|---|---|
| Female | 4,068 (58.7%) | 23,423 (55.2%) | 26,879 (66.2%) | 67,610 (59.3%) |
| Male | 2,865 (41.3%) | 19,044 (44.8%) | 13,716 (33.8%) | 46,411 (40.7%) |
| Age | | | | |
| Mean (SD) | 48.3 (16.9) | 54.4 (14.4) | 49.7 (15.6) | 59.9 (16.6) |
| Median [Min, Max] | 45.0 [21.0, 95.0] | 57.0 [19.0, 95.0] | 49.0 [20.0, 95.0] | 63.0 [18.0, 95.0] |
| BMI | | | | |
| Mean (SD) | 25.1 (4.72) | 30.6 (7.62) | 30.1 (6.59) | 28.8 (6.56) |
| Median [Min, Max] | 24.3 [17.0, 55.0] | 29.4 [17.0, 55.0] | 29.1 [17.0, 55.0] | 27.5 [17.0, 55.0] |
| AF = No | 6,830 (98.5%) | 41,343 (97.4%) | 39,626 (97.6%) | 106,774 (93.6%) |
| AF = Yes | 103 (1.5%) | 1,124 (2.6%) | 969 (2.4%) | 7,247 (6.4%) |
| **CAD** | **Asian (N=6,933)** | **Black (N=42,469)** | **Hispanic/Latino (N=40,591)** | **White (N=114,012)** |
| Female | 4,068 (58.7%) | 23,427 (55.2%) | 26,873 (66.2%) | 67,604 (59.3%) |
| Male | 2,865 (41.3%) | 19,042 (44.8%) | 13,718 (33.8%) | 46,408 (40.7%) |
| Age | | | | |
| Mean (SD) | 48.3 (16.8) | 54.3 (14.4) | 49.7 (15.5) | 59.9 (16.7) |
| Median [Min, Max] | 45.0 [21.0, 95.0] | 57.0 [18.0, 95.0] | 49.0 [20.0, 95.0] | 63.0 [20.0, 95.0] |
| BMI | | | | |
| Mean (SD) | 25.1 (4.72) | 30.6 (7.62) | 30.1 (6.59) | 28.8 (6.57) |
| Median [Min, Max] | 24.3 [17.0, 55.0] | 29.4 [17.0, 55.0] | 29.1 [17.0, 55.0] | 27.5 [17.0, 55.0] |
| CAD = No | 6,816 (98.3%) | 41,334 (97.3%) | 3,9870 (98.2%) | 10,9142 (95.7%) |
| CAD = Yes | 117 (1.7%) | 1,135 (2.7%) | 721 (1.8%) | 4,870 (4.3%) |
| **CKD** | **Asian (N=6,931)** | **Black (N=42,470)** | **Hispanic/Latino (N=40,584)** | **White (N=113,964)** |
| Female | 4,066 (58.7%) | 23,420 (55.1%) | 26,872 (66.2%) | 67,573 (59.3%) |
| Male | 2,865 (41.3%) | 19,050 (44.9%) | 13,712 (33.8%) | 46,391 (40.7%) |
| Age | | | | |
| Mean (SD) | 48.3 (16.9) | 54.2 (14.4) | 49.6 (15.6) | 60.1 (16.8) |
| Median [Min, Max] | 45.0 [19.0, 95.0] | 57.0 [18.0, 95.0] | 49.0 [18.0, 95.0] | 63.0 [19.0, 95.0] |
| BMI | | | | |
| Mean (SD) | 25.1 (4.72) | 30.6 (7.62) | 30.1 (6.59) | 28.8 (6.57) |
| Median [Min, Max] | 24.3 [17.0, 55.0] | 29.4 [17.0, 55.0] | 29.1 [17.0, 55.0] | 27.5 [17.0, 55.0] |
| CKD = No | 6,809 (98.2%) | 40,093 (94.4%) | 39,093 (96.3%) | 108,990 (95.6%) |
| CKD = Yes | 122 (1.8%) | 2,377 (5.6%) | 1,491 (3.7%) | 4,974 (4.4%) |
| **HF** | **Asian (N=6,932)** | **Black (N=42,461)** | **Hispanic/Latino (N=40,588)** | **White (N=113,962)** |
| Female | 4,067 (58.7%) | 234,17 (55.1%) | 26,872 (66.2%) | 67,582 (59.3%) |
| Male | 2,865 (41.3%) | 19,044 (44.9%) | 13,716 (33.8%) | 46,380 (40.7%) |
| Age | | | | |
| Mean (SD) | 48.4 (17.0) | 54.4 (14.4) | 49.7 (15.6) | 60.2 (16.8) |
| Median [Min, Max] | 45.0 [21.0, 95.0] | 57.0 [20.0, 95.0] | 49.0 [19.0, 95.0] | 63.0 [20.0, 95.0] |
| BMI | | | | |
| Mean (SD) | 25.1 (4.73) | 30.6 (7.62) | 30.1 (6.59) | 28.8 (6.57) |
| Median [Min, Max] | 24.3 [17.0, 55.0] | 29.4 [17.0, 55.0] | 29.1 [17.0, 55.0] | 27.5 [17.0, 55.0] |
| HF = No | 6,867 (99.1%) | 40,619 (95.7%) | 39,644 (97.7%) | 110,023 (96.5%) |
| HF = Yes | 65 (0.9%) | 1842 (4.3%) | 944 (2.3%) | 3,939 (3.5%) |
| **HTN** | **Asian (N=6,939)** | **Black (N=42,484)** | **Hispanic/Latino (N=40,579)** | **White (N=114,170)** |
| Female | 4,074 (58.7%) | 23,438 (55.2%) | 26,872 (66.2%) | 67,680 (59.3%) |
| Male | 2,865 (41.3%) | 19,046 (44.8%) | 13,707 (33.8%) | 46,490 (40.7%) |
| Age | | | | |

| | | | | |
|---|---|---|---|---|
| Mean (SD) | 47.0 (16.0) | 51.3 (13.6) | 47.5 (14.5) | 57.0 (15.8) |
| Median [Min, Max] | 44.0 [18.0, 95.0] | 53.0 [18.0, 94.0] | 47.0 [18.0, 95.0] | 59.0 [18.0, 95.0] |
| BMI | | | | |
| Mean (SD) | 25.1 (4.74) | 30.6 (7.64) | 30.1 (6.60) | 28.8 (6.59) |
| Median [Min, Max] | 24.3 [17.0, 55.0] | 29.4 [17.0, 55.0] | 29.2 [17.0, 55.0] | 27.5 [17.0, 55.0] |
| HTN = No | 5,793 (83.5%) | 27,507 (64.7%) | 30,006 (73.9%) | 74,236 (65.0%) |
| HTN =Yes | 1,146 (16.5%) | 14,977 (35.3%) | 10,573 (26.1%) | 39,934 (35.0%) |
| **T2DM** | **Asian (N=6,931)** | **Black (N=42,480)** | **Hispanic/Latino (N=40,594)** | **White (N=113,925)** |
| Female | 4,064 (58.6%) | 23,436 (55.2%) | 26,874 (66.2%) | 67,564 (59.3%) |
| Male | 2,867 (41.4%) | 19,044 (44.8%) | 13,720 (33.8%) | 46,361 (40.7%) |
| Age | | | | |
| Mean (SD) | 48.2 (16.9) | 54.1 (14.3) | 49.4 (15.5) | 60.0 (16.8) |
| Median [Min, Max] | 45.0 [21.0, 95.0] | 56.0 [18.0, 95.0] | 49.0 [18.0, 95.0] | 63.0 [18.0, 95.0] |
| BMI | | | | |
| Mean (SD) | 25.1 (4.74) | 30.6 (7.63) | 30.1 (6.61) | 28.8 (6.57) |
| Median [Min, Max] | 24.3 [17.0, 55.0] | 29.4 [17.0, 55.0] | 29.1 [17.0, 55.0] | 27.5 [17.0, 55.0] |
| T2DM = No | 6,706 (96.8%) | 39,066 (92.0%) | 37,933 (93.4%) | 10,7915 (94.7%) |
| T2DM = Yes | 225 (3.2%) | 3,414 (8.0%) | 2,661 (6.6%) | 6,010 (5.3%) |

Sex was based on sex assigned at birth. Summary statistics are stratified by self-reported race/ethnicity group.

Abbreviations: BMI: body mass index; AF: atrial fibrillation; CAD: coronary artery disease; CKD: chronic kidney disease; HF: heart failure; HTN: hypertension; T2DM: type 2 diabetes mellitus.

#### Supplementary Table 9: Standard concept names used to define outcomes in the AoU study

| Outcome | OMOP concept ID | SNOMED ID | Number of cases |
|---|---|---|---|
| Atrial fibrillation (AF) | 313217 | 49436004 | 11,342 |
| Coronary arteriosclerosis (CAD) | 317576 | 53741008 | 8,321 |
| Chronic Kidney disease (CKD) | 46271022 | 709044004 | 11,094 |
| Essential hypertension (HTN) | 320128 | 59621000 | 79,612 |
| Heart failure (HF) | 316139 | 84114007 | 8,688 |
| Type 2 diabetes mellitus (T2DM) | 201826 | 44054006 | 14,786 |

Note: number of cases was summed across all self-reported race/ethnicity groups.

#### Supplementary Table 10: Publicly available PGS used for constructing PRSs in AoU

| Outcome | Polygenic score ID | PGS publication ID | PubMed ID | Number of variants used |
|---|---|---|---|---|
| Atrial fibrillation (AF) | PGS002814 | PGP000392 | 36653681 | 2,783 |
| Coronary arteriosclerosis (CAD) | PGS004698 | PGP000602 | 38380516 | 156,064 |
| Chronic Kidney disease (CKD) | PGS004016 | PGP000517 | 38908374 | 45,419 |
| Essential hypertension (HTN) | PGS004234 | PGP000531 | 35729114 | 136,347 |
| Heart failure (HF) | PGS000709 | PGP000128 | 33462484 | 111,505 |
| Type 2 diabetes mellitus (T2DM) | PGS000036 | PGP000023 | 30297969 | 50,816 |

Note: The PGS variant and weights were downloaded from the PGS Catalog, with the PGS publication ID number shown in the third column. The last column summarizes the number of variants used for computing the PRS

#### Supplementary Figures

##### Supplementary Figure 1: Estimated PRS effect size $\beta_1$ in simulations comparing genetic confounding from one or two variants.

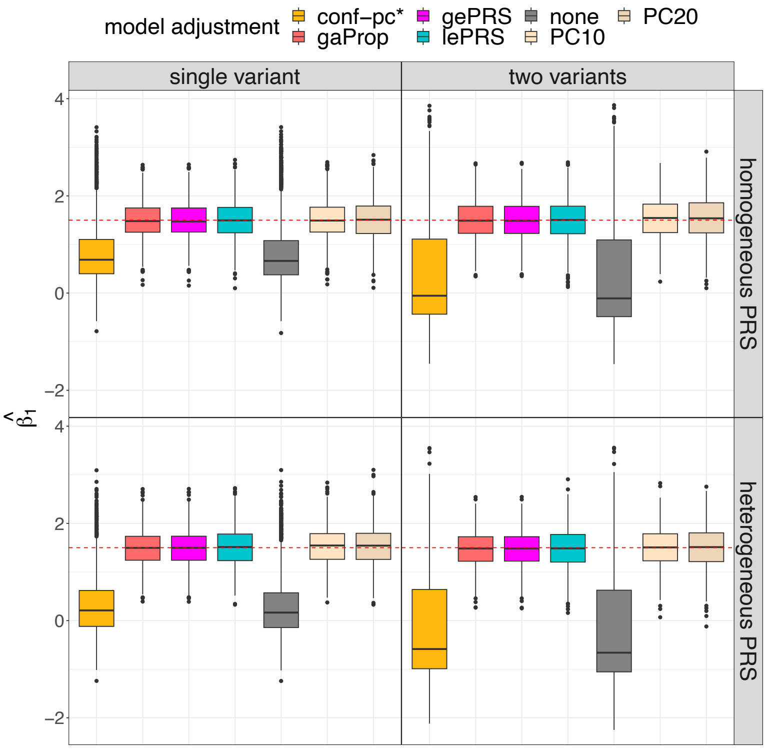

The figure provides the estimated PRS-outcome association (shown in the y-axis) from each association models for the single variant (left panel) and two variants (right panel) unknown genetic confounding settings. Results on the top row correspond to data generating model with homogenous weighting PRS, and results in the bottom row correspond to heterogeneous weighting PRS. The dashed red line is the underlying true $\beta_1$ value. For the PRS model, the covariates to adjust includes: PC10, PC20 (adjusting for top 10 and 20 genetic PCs, respectively), none (no covariate adjustment), conf-pc* (adjustment for a known confounder, for benchmarking), and gaProp (adjustment for global proportions of ancestry). For ePRS model, the covariates to adjust are global ePRS (gePRS) and local ePRS (lePRS), respectively. Distributions are provided from 1000 simulation repetitions.

Abbreviations: conf: confounding; ePRS: expected PRS; gePRS: global ancestry expected PRS; lePRS: local ancestry expected PRS; PC: principal component; PRS: polygenic risk score; gaProp: global ancestry proportion.

#### Supplementary Figure 2: Mean squared error from estimation of $\beta_1$ in simulations comparing genetic confounding from one or two variants.

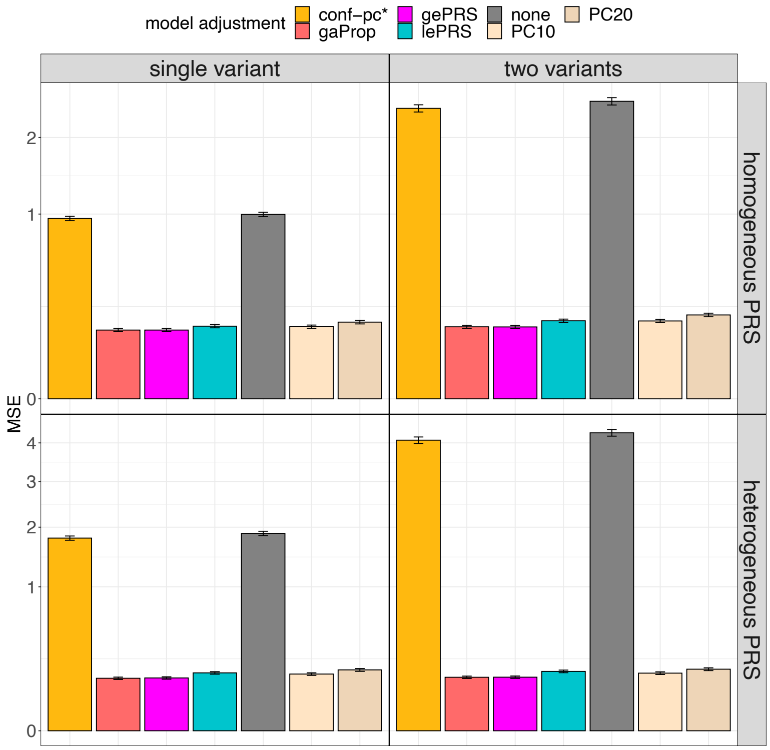

The figure provides bar plots of the visualizing the MSE of estimated effect sizes of the PRS ($\hat{\beta}_1$) for single variant (left panel) and two variants (right panel) unknown genetic confounding settings. Results on the top row correspond to data generating model with homogenous weighting PRS, and results in the bottom row correspond to heterogeneous weighting PRS. The y-axis shows the MSE value. For the PRS model, the covariates to adjust includes: PC10, PC20 (adjusting for top 10 and 20 genetic PCs, respectively), none (no covariate adjustment), conf-pc* (adjustment for a known confounder, for benchmarking), and gaProp (adjustment for global proportions of ancestry). For ePRS model, the covariates to adjust are global ePRS (gePRS) and local ePRS (lePRS), respectively. MSEs were computed over 1000 simulation repetitions. Intervals around the estimated MSE correspond to the MSE +/- one estimated standard error.
Abbreviations: MSE: mean square error; conf: confounding; ePRS: expected PRS; gePRS: global ancestry expected PRS; lePRS: local ancestry expected PRS; PC: principal component; PRS: polygenic risk score; gaProp: global ancestry proportion.

#### Supplementary Figure 3: MSE of the estimation of $\beta_1$ in simulations comparing genetic confounding from one or two variants, by increasing confounding effect.

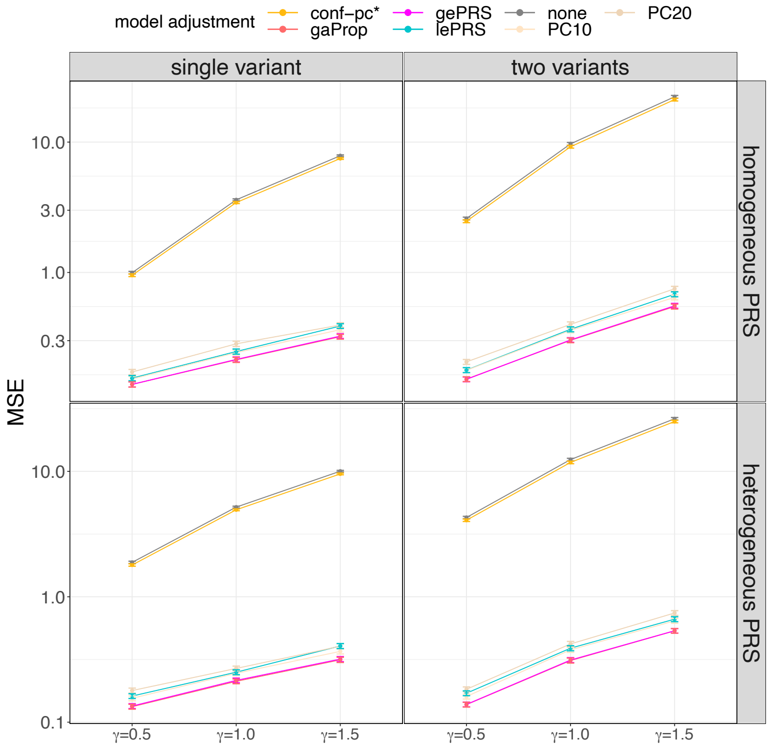

The figure provides the estimated MSE of the estimated PRS effect size ($\hat{\beta}_1$) for single variant (left panel) and two variants (right panel) unknown genetic confounding settings. Results on the top row correspond to data generating model with homogenous weighting PRS, and results in the bottom row correspond to heterogeneous weighting PRS. Each panel provides MSE (y-axis) obtained across 1000 simulation repetition with association analyses using 7 combinations of PRS and adjustment approaches, and across 3 simulated effect sizes ($\gamma$) of the genetic ancestry-related confounder. In this simulation, we fixed $\beta_1 = 1.5$ in the data generating model across all settings. Association analyses that estimate (standard) PRS effect while adjusting to covariates include PC10, PC20 (adjusting for top 10 and 20 genetic PCs, respectively), none (no covariate adjustment), pc* (adjustment for a known confounder, for benchmarking), and gaProp (adjustment for global proportions of ancestry). Association analyses applying the ePRS framework include gePRS and lePRS (estimation of rRPS effect adjusting for ePRS, based on local and local models, respectively). MSEs were computed over 1000 simulation repetitions. Intervals around the estimated MSE correspond to the MSE +/- one estimated standard error.

Abbreviations: MSE: mean square error; conf: confounding; ePRS: expected PRS; gePRS: global ancestry expected PRS; lePRS: local ancestry expected PRS; MSA: mean squared error; PC: principal component; PRS: polygenic risk score; gaProp: global ancestry proportion.

#### Supplementary Figure 4: Results from simulations applying ePRS under random errors in ancestry.

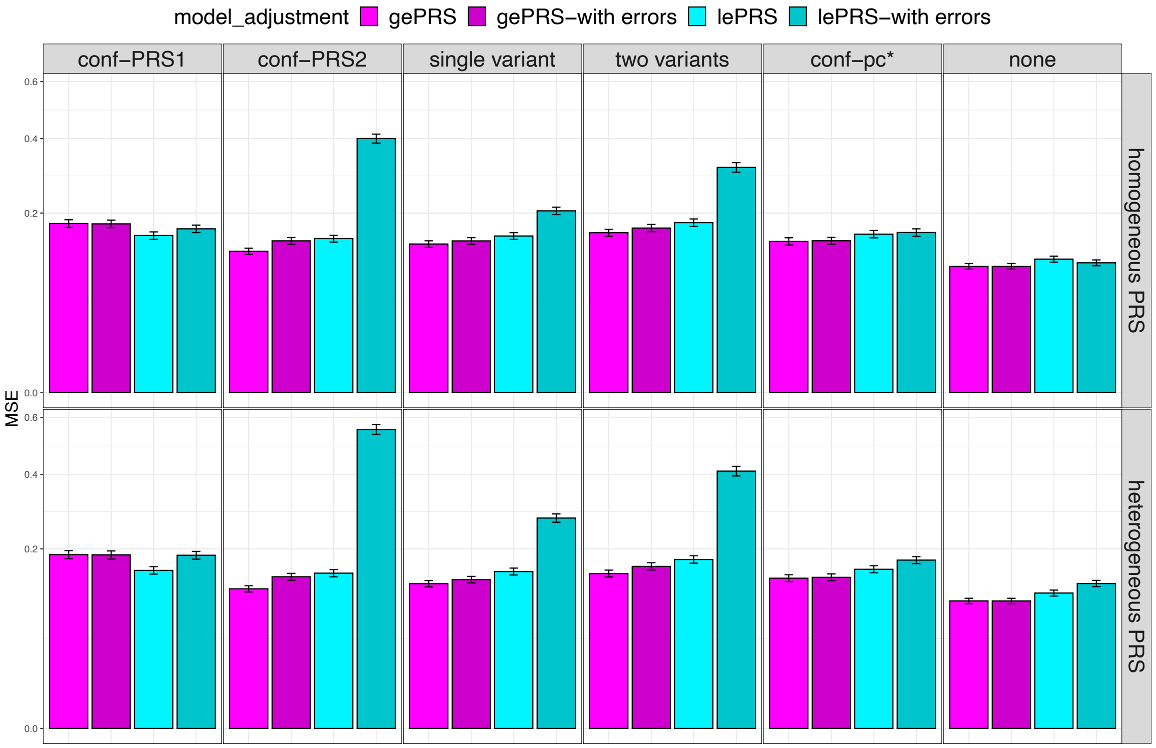

The MSE from the estimation of $\beta_1$ are shown in the y-axis. Results from ePRS calculated by using global or local ancestry information with random error are shown in gePRS-with errors and lePRS-with errors. Results on the top row correspond to data generating model with homogenous weighting PRS, and results in the bottom row correspond to heterogeneous weighting PRS. Columns correspond to different genetic ancestry-related confounding factors defined in supplementary table 1: conf-PRS1, conf-PRS2, single variable, two variables, conf-pc*, and no confounding (from the left to the right). The MSE (y-axis) was obtained across 1000 simulation repetition and the intervals around the estimated MSE correspond to the MSE +/- one estimated standard error. Abbreviations: MSE: mean square error; conf: confounding; gePRS: global expected PRS; lePRS: local expected PRS; PRS: polygenic risk score.

Supplementary Figure 5: Estimated effect sizes of ePRSs and rPRSs in simulations when varying confounding strengths.

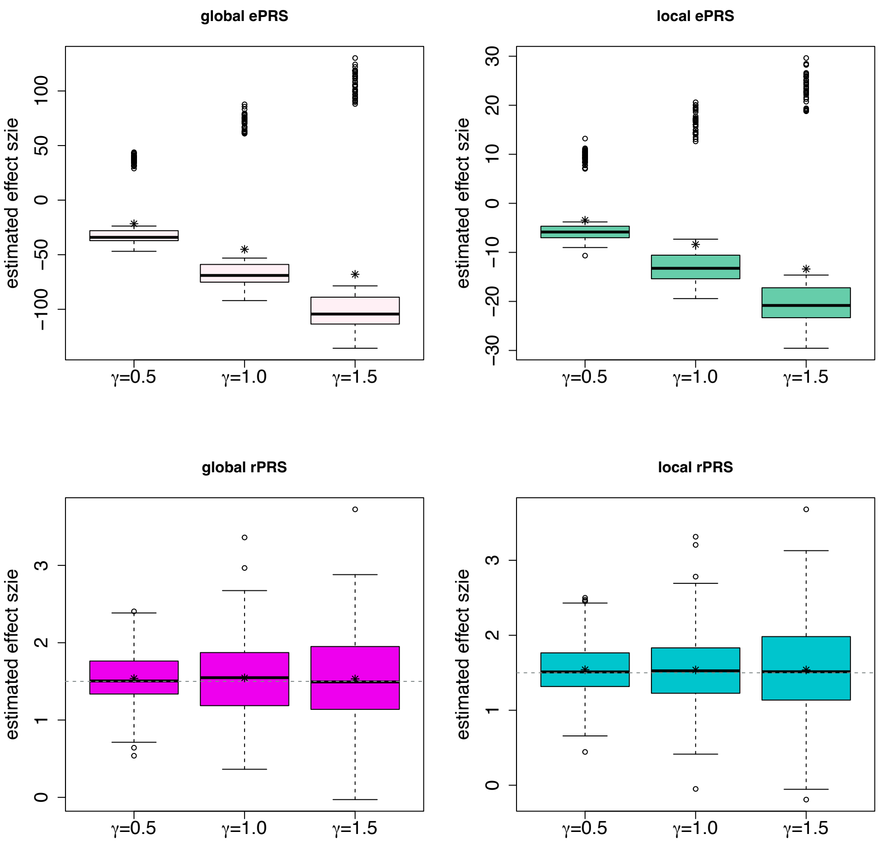

The figure visualizes the estimated effect sizes of the ePRS (top) and rPRS (bottom) in simulations, for different confounding effect sizes. The confounding used in this analysis is the single genetic variant setting (third row of Supplementary Table 1). The results of global ePRS and rPRS are shown in the left panel. The results of local ePRs and rPRS are shown in the right panel. The y-axis of each panel is the estimated effect size across 1000 replications.

Abbreviation: ePRS: expected PRS; rPRS: residual PRS; PRS: polygenic risk score.

#### Supplementary Figure 6: Estimated effect sizes of the ePRS and rPRS in simulations comparing homogeneous and heterogeneous ancestry weighting PRSs.

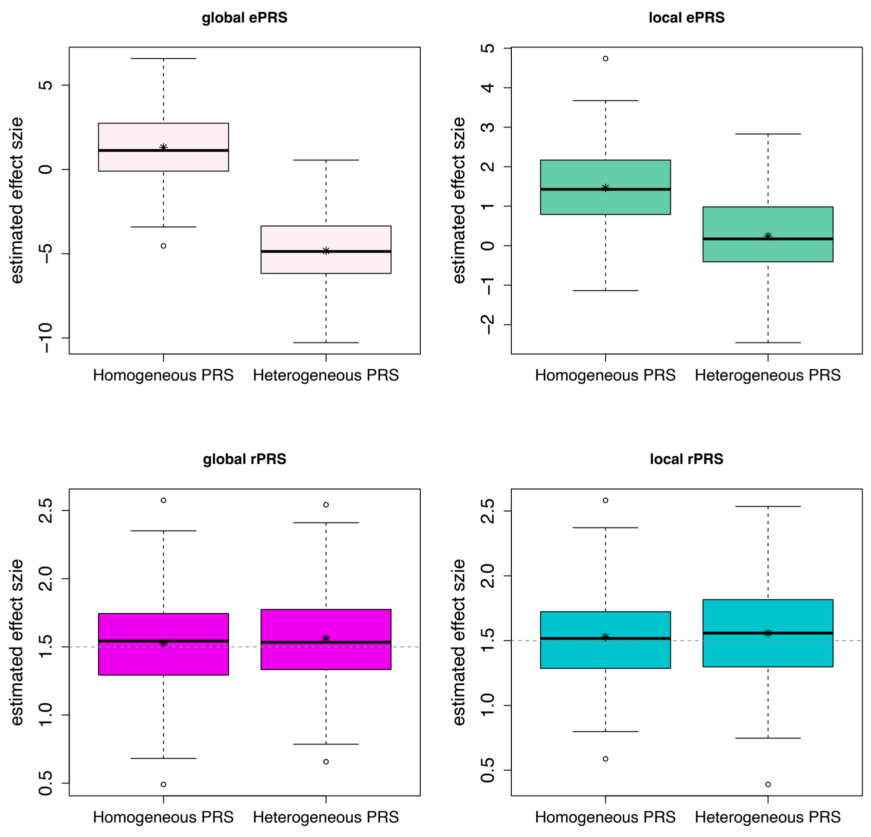

The figure visualizes the estimated effect sizes of the ePRS (top) and rPRS (bottom) in simulations when considering different types of PRSs in the data generating model. No confounding was considered in this analysis. Box plots of effects estimates of global ePRS and rPRS are shown in the left panels, while the boxplots on the right panels who results from estimation using local ePRs nad rPRS. The y-axis of each panel is the estimated effect size across 1000 replications.

Abbreviation: ePRS: expected PRS; rPRS: residual PRS; PRS: polygenic risk score.

#### Supplementary Figure 7: Estimated AUCs from simulation studies applying PRS approaches for disease risk classification.

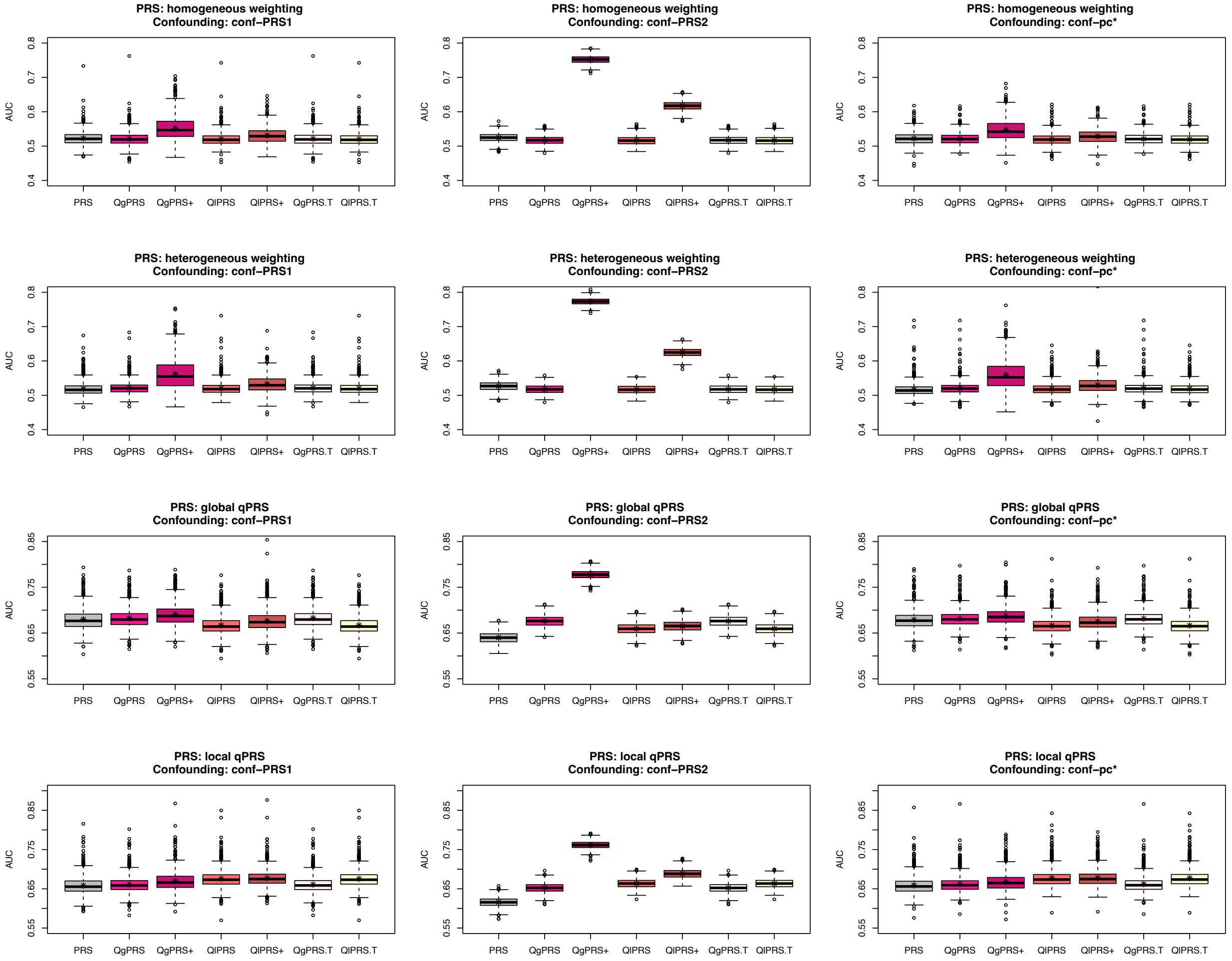

The figure provides boxplots visualizing computed AUCs across 1000 simulation replicates from each data generating and working association model. Each panel corresponds to a specific data generating model, defined by combinations of causal PRS (rows) and genetic confounding (columns). Ancestry-related confounding factors were set to conf-PRS1, conf-PRS2, or conf-pc*, and causal PRS measures were homogeneous or heterogeneous weighting PRS, global qPRS or local qPRS. Each panel provides results from working association models with various PRS measures used: QgPRS+ and QlPRS+ represent models adjusted for both qPRS and ePRS (global and local, respectively). The QgPRS and QlPRS represent the model only adjust qPRS. The QgPRS.T and QlPRS.T present the method directly using the qPRS to conduct classification without fitting a prediction model.
Abbreviations: AUC: area under the ROC curve; QgePRS: quantile global ePRS; QlePRS: quantile local ePRS; ePRS: expected PRS; PRS: polygenic risk score.

#### Supplementary Figure 8: Scatterplots visualizing PRS versus global ePRS values in TOPMed analysis.

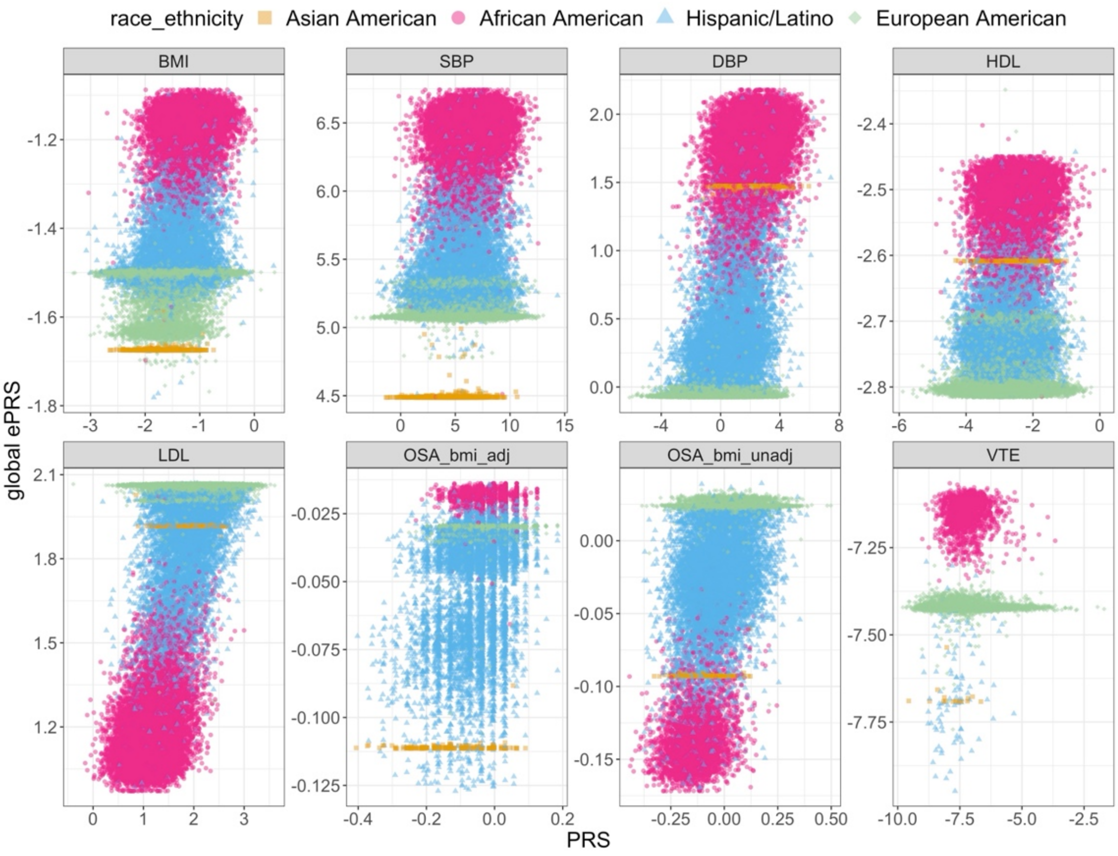

The figure visualizes the relationship between PRS and global ePRS using BMI, SBP, DBP, HDL, LDL, OSA, and VTE PRSs from the main analysis (i.e., PRSs based on genome-wide significant variants). In each panel, each point represents an individual's global ePRS (y-axis) and PRS (x-axis) values. Each point's color and style represents self-reported race/ethnicity. OSA_bmi_adj and OSA_bmi_unadj PRSs are based on OSA that either adjusted for, or did not adjust for, BMI in the association model, respectively. Global ePRSs were constructed based on each individual's global ancestry proportion and the ancestry-specific allele frequency estimated using GAFA. Abbreviations: BMI: body mass index; SBP: systolic blood pressure; DBP: diastolic blood pressure; HDL: high-density lipoprotein; LDL: low-density lipoprotein; OSA: obstructive sleep apnea; VTE: venous thromboembolism; TOPMed: Trans-Omics for Precision Medicine; PRS: polygenic risk scores; ePRS: expected polygenic risk scores.

Supplementary Figure 9: Scatterplots visualizing PRS versus global rPRS values in TOPMed.

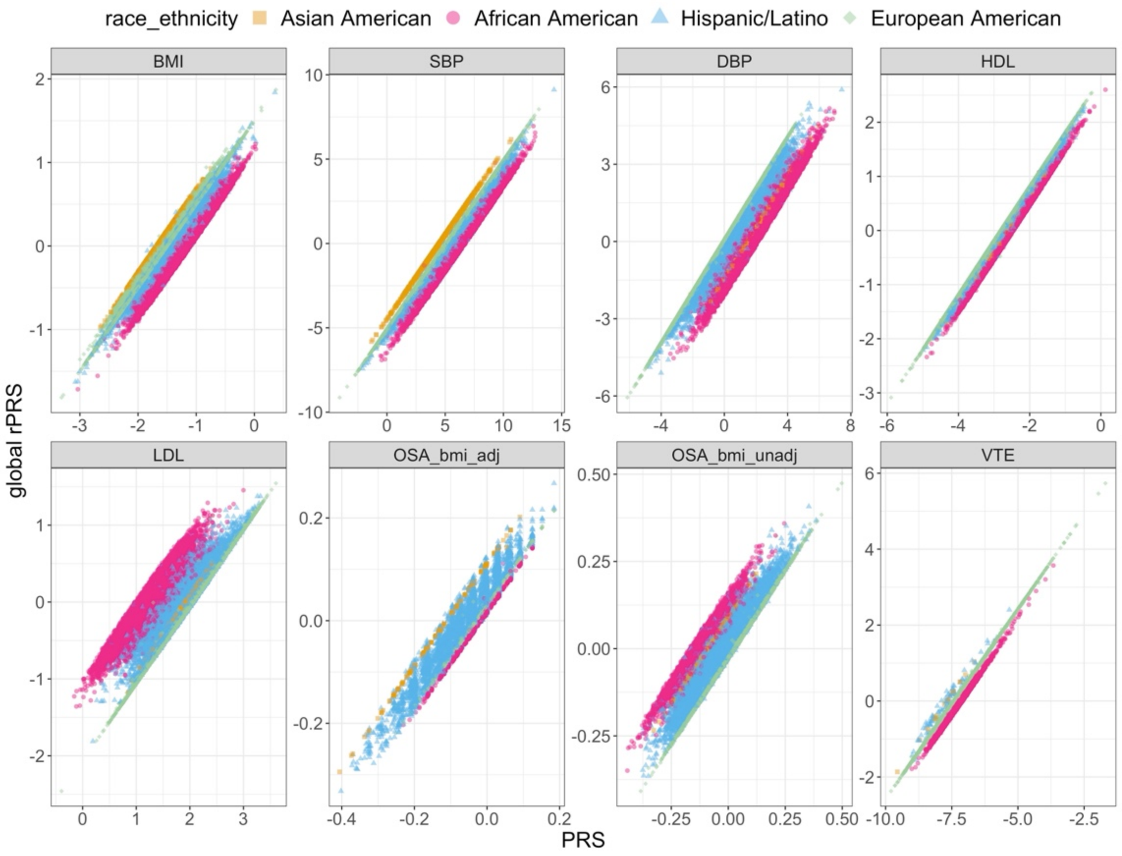

The figure visualizes the relationship between PRS (x-axis) and global rPRS (y-axis) values in TOPMed individuals, with PRS measures corresponding to BMI, SBP, DBP, HDL, LDL, OSA_BMI_adj, OSA_bmi_unadj, and VTE. The global rPRS was calculated as the difference between the PRS and global ePRS. The points' color and style represent self-reported race/ethnicity groups. OSA_bmi_adj and OSA_bmi_unadj PRSs are based on OSA that either adjusted for, or did not adjust for, BMI in the association model, respectively.
Abbreviations: BMI: body mass index; SBP: systolic blood pressure; DBP: diastolic blood pressure; HDL: high-density lipoprotein; LDL: low-density lipoprotein; OSA: obstructive sleep apnea; VTE: venous thromboembolism; TOPMed: Trans-Omics for Precision Medicine; PRS: polygenic risk scores; ePRS: expected polygenic risk scores; rPRS: residual PRS.

#### Supplementary Figure 10: Density plots of PRSs.

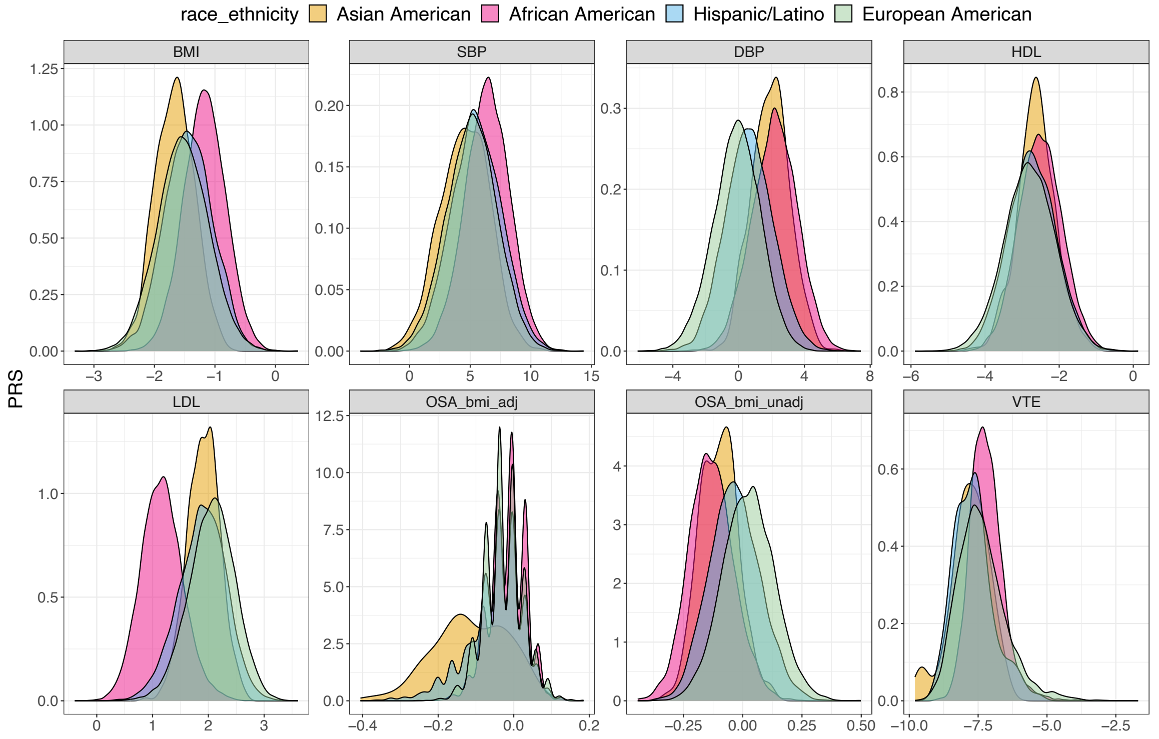

The figure visualizes PRS distributions in TOPMed participants, with PRS corresponding to BMI, SBP, DBP, HDL, LDL, OSA_BMI_adj, OSA_bmi_unadj, and VTE. Density plots are stratified by self-reported race/ethnicity groups. OSA_bmi_adj and OSA_bmi_unadj PRSs are based on OSA that either adjusted for, or did not adjust for, BMI in the association model, respectively.

Abbreviations: BMI: body mass index; SBP: systolic blood pressure; DBP: diastolic blood pressure; HDL: high-density lipoprotein; LDL: low-density lipoprotein; OSA: obstructive sleep apnea; VTE: venous thromboembolism; TOPMed: Trans-Omics for Precision Medicine; PRS: polygenic risk scores.

#### Supplementary Figure 11: Density plots of global ePRSs.

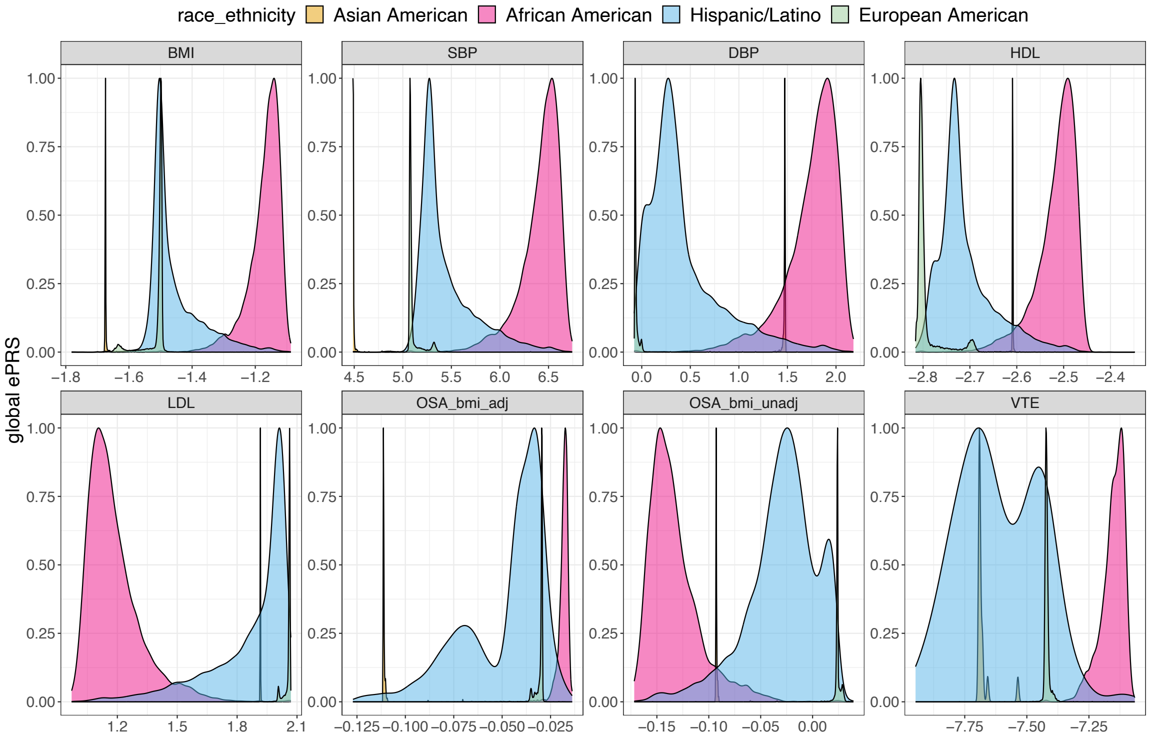

The figure visualizes global ePRS distributions in TOPMed participants, with ePRSs corresponding to BMI, SBP, DBP, HDL, LDL, OSA_BMI_adj, OSA_bmi_unadj, and VTE. Density plots are stratified by self-reported race/ethnicity groups. OSA_bmi_adj and OSA_bmi_unadj PRSs are based on OSA that either adjusted for, or did not adjust for, BMI in the association model, respectively. To better visualize the density plot for ePRS across populations, we re-scaled the density and let the y-axis values range from 0 to 1. Global ePRSs were constructed based on each individual's global ancestry proportion and the ancestry-specific allele frequency estimated using GAFA.

Abbreviations: BMI: body mass index; SBP: systolic blood pressure; DBP: diastolic blood pressure; HDL: high-density lipoprotein; LDL: low-density lipoprotein; OSA: obstructive sleep apnea; VTE: venous thromboembolism; TOPMed: Trans-Omics for Precision Medicine; ePRS: expected PRS; PRS: polygenic risk scores.

#### Supplementary Figure 12: Density plot of global rPRSs.

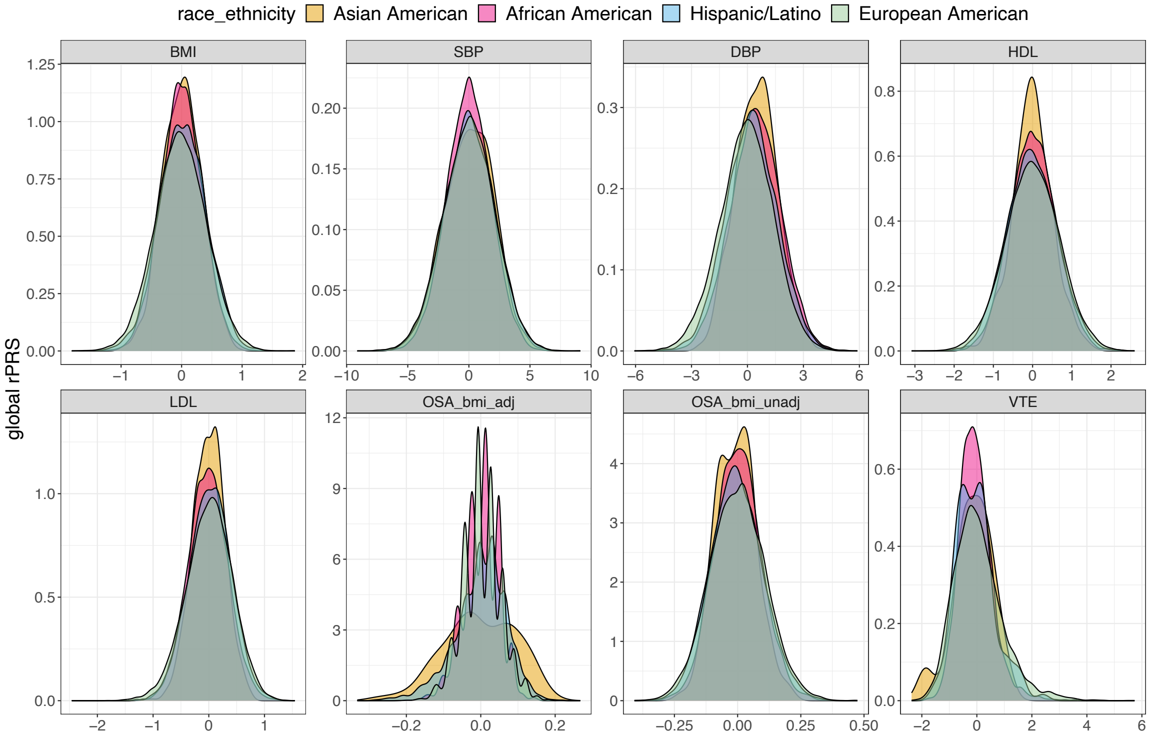

The figure visualizes global rPRS distributions in TOPMed participants, with rPRSs corresponding to BMI, SBP, DBP, HDL, LDL, OSA_BMI_adj, OSA_bmi_unadj, and VTE. Each rPRS was constructed as the difference between an individual's PRS and their global ePRS. Density plots are stratified by self-reported race/ethnicity groups. OSA_bmi_adj and OSA_bmi_unadj PRSs are based on OSA that either adjusted for, or did not adjust for, BMI in the association model, respectively.

Abbreviations: BMI: body mass index; SBP: systolic blood pressure; DBP: diastolic blood pressure; HDL: high-density lipoprotein; LDL: low-density lipoprotein; OSA: obstructive sleep apnea; VTE: venous thromboembolism; TOPMed: Trans-Omics for Precision Medicine; rPRS: expected PRS; PRS: polygenic risk scores.

#### Supplementary Figure 13: The association between PRS, rPRS, and qPRS percentiles and SBP, DBP, and HDL values.

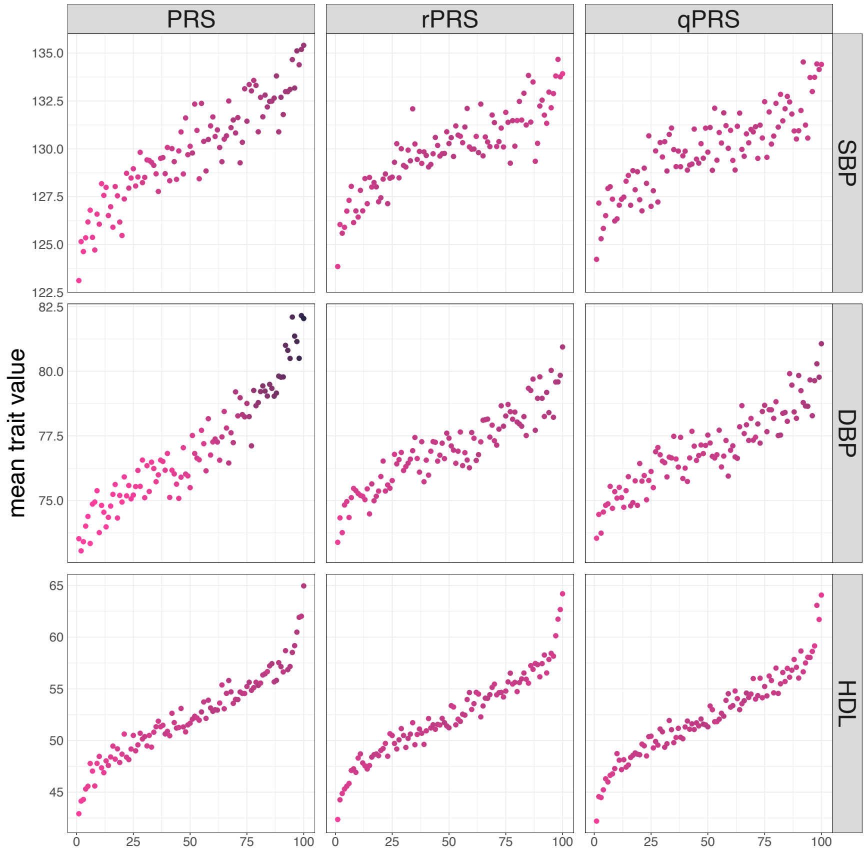

Each panel is a scatterplot visualizing, on the x-axis, percentiles of PRS (left column panels), global rPRS (middel column panels), and global qPRS (right column panels), against mean values of the corresponding phenotypes (y-axis), averaged across individuals with the corresponding PRS, rPRS, or qPRS percentile. For instance, for each phenotype and PRS measure, individuals were binned into 100 strata defined by PRS/qPRS percentiles. The color of a given point corresponds to the proportion of African American individuals among individuals in the PRS, rPRS, or qPRS percentile stratum. Darker color reflects a higher proportion of African American individuals. Abbreviations: TOPMed: Trans-Omics for Precision Medicine; SBP: systolic blood pressure; DBP: diastolic blood pressure; HDL: high-density lipoprotein; PRS: polygenic risk score; rPRS: residual PRS; qPRS: quantile PRS.

#### Supplementary Figure 14: The association between PRS, rPRS, and qPRS percentiles and OSA and VTE proportions.

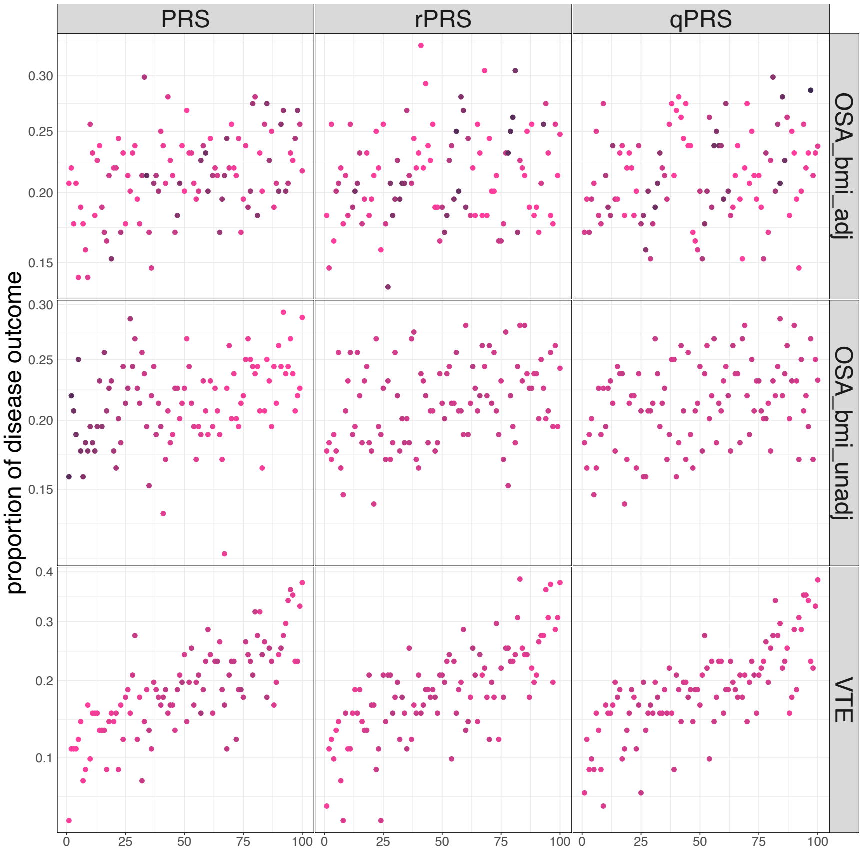

Each panel is a scatterplot visualizing, on the x-axis, percentiles of PRS (left column panels), rPRS (middle column panels), and global qPRS (right column panels), against proportions of individuals with the corresponding disease outcome (y-axis), computed across individuals with the corresponding PRS, rPRS, or qPRS percentile. For instance, for each phenotype and PRS measure, individuals were binned into 100 strata defined by PRS, rPRS or qPRS percentiles. The color of a given point corresponds to the proportion of African American individuals among individuals in the PRS, rPRS, or qPRS percentile stratum. Darker color reflects a higher proportion of African American individuals. Results based on PRS calculated based on OSA GWAS with BMI adjustment are shown in the first row, and the results based on PRS calculated based on OSA GWAS without BMI adjustment are shown in the second row.
Abbreviations: TOPMed: Trans-Omics for Precision Medicine; OSA: obstructive sleep apnea; VTE: venous thromboembolism PRS: polygenic risk score; rPRS: residual PRS; qPRS: quantile PRS.

### Supplementary Figure 15: Estimated LDpred 2 PRS-outcome effect sizes.

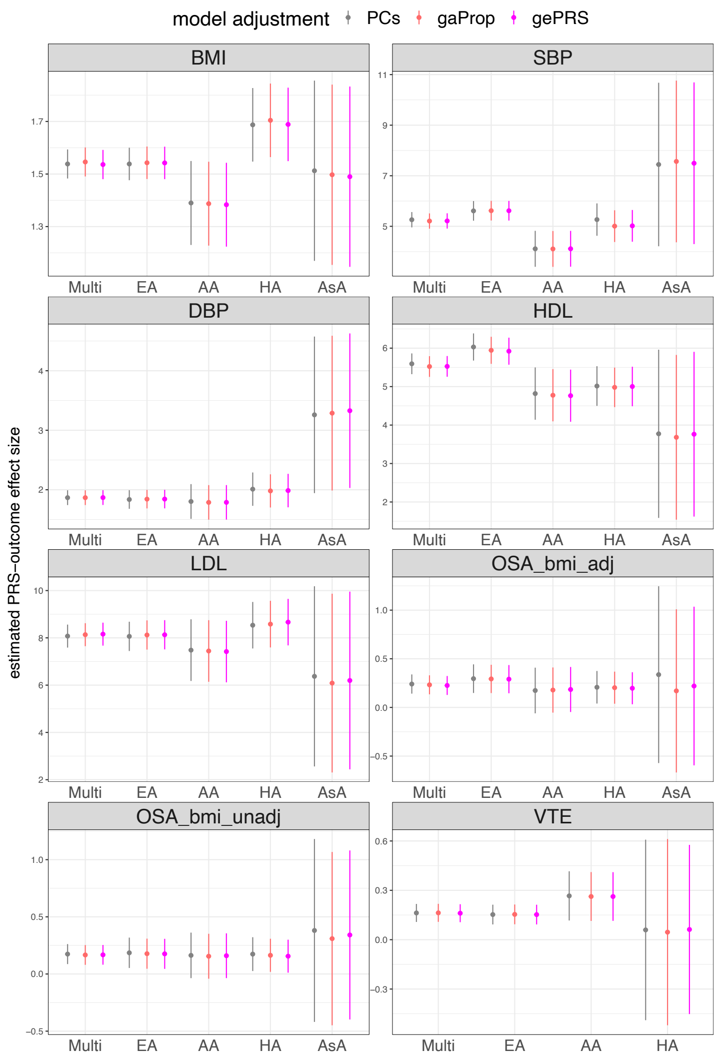

Estimated PRS-outcome effect sizes (y-axis) and their corresponding 95% confidence intervals in the TOPMed dataset. The PRS and ePRS was constructed by using the weights generated from LDPred2. Conventional PRS model estimated the PRS effect size by either adjusting for genetic PCs or for global ancestry proportions. For the ePRS model, we show the estimated rPRS effect size adjusting for global ePRS. Estimated association are provided, for each trait, for the combined dataset (Multi), and stratified by self-reported race/ethnicity. For continuous traits (BMI, SBP, DBP, HDL, and LDL), effect sizes are in the original trait scale (kg/m$^2$, mmHg, mmol/L). OSA and VTE are binary traits, and their estimated effect sizes are in the log odds ratio scale. For OSA, we provide results for two OSAs: OSA_bmi_adj and OSA_bmi_unadj, based on GWAS that did and did not adjust for BMI, respectively. Due to the limited sample size, we did not perform an Asian-specific analysis of VTE. Abbreviations: BMI: body mass index; SBP: systolic blood pressure; DBP: diastolic blood pressure; HDL: high-density lipoprotein; LDL: low-density lipoprotein; OSA: obstructive sleep apnea; VTE: venous thromboembolism; Multi: Multi-ethnic; EA: European American; AA: African American; HA: Hispanic/Latino; AsA: Asian American; PRS: polygenic risk score; PC: principal component; gaProp: global ancestry proportion; gePRS: global ancestry expected PRS.

#### Supplementary Figure 16: Density plots of PRSs in AoU.

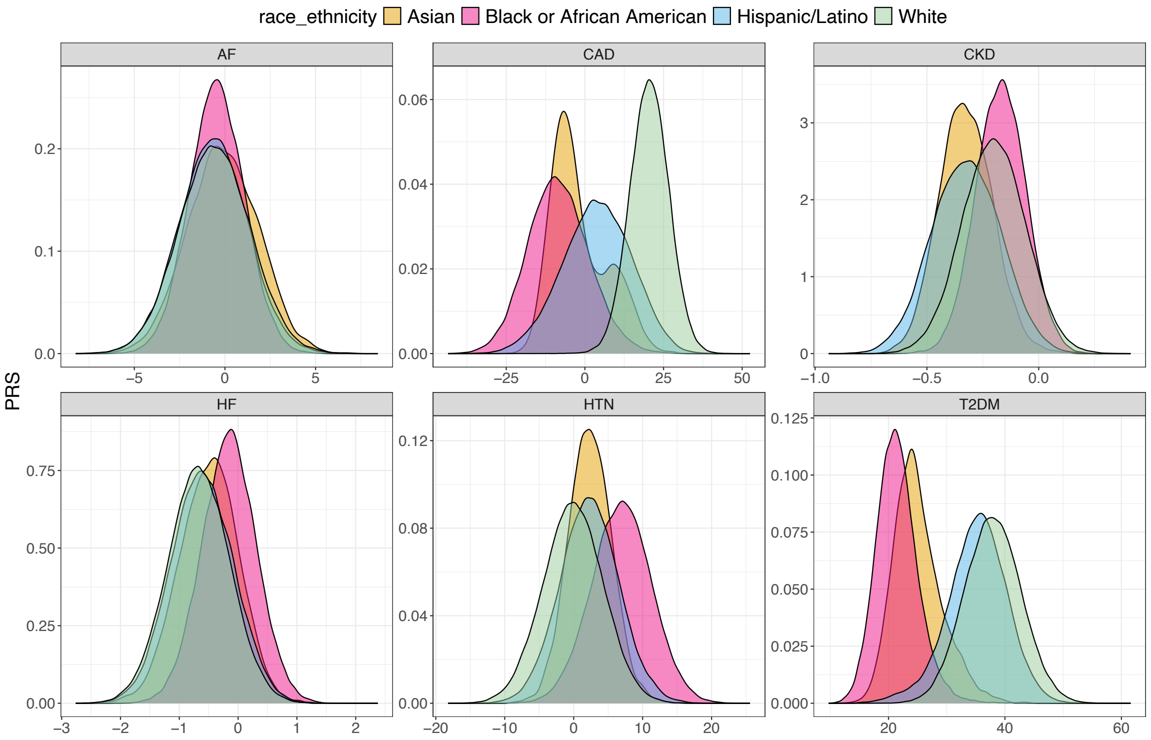

The figure visualizes PRS distributions in AoU participants, with PRS corresponding to AF, CAD, CKD, HF, HTN, and T2DM. Density plots are stratified by self-reported race/ethnicity groups.
Abbreviations: AoU: All of Us; PRS: polygenic risk score; AF: atrial fibrillation; CAD: coronary artery disease; CKD: chronic kidney disease; HF: heart failure; HTN: hypertension; T2DM: type 2 diabetes mellitus.

### Supplementary Figure 17: Density plots of global ePRSs in AoU.

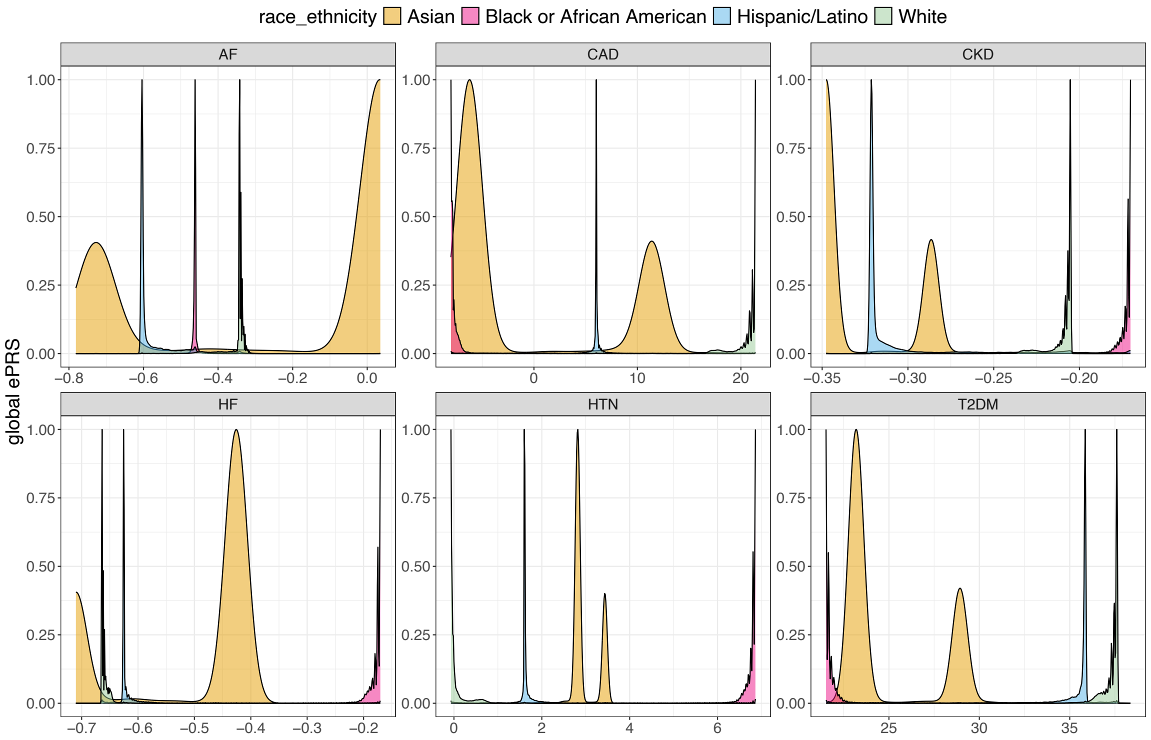

The figure visualizes global ePRS distributions in AoU participants, with ePRSs corresponding to AF, CAD, CKD, HF, HTN, and T2DM. Density plots are stratified by self-reported race/ethnicity groups. To better visualize the density plot for ePRS across populations, we re-scaled the density and let the y-axis values range from 0 to 1. Abbreviations: AoU: All of Us; ePRS: expected PRS; PRS: polygenic risk score; AF: atrial fibrillation; CAD: coronary artery disease; CKD: chronic kidney disease; HF: heart failure; HTN: hypertension; T2DM: type 2 diabetes mellitus.

#### Supplementary Figure 18: Density plots of rPRSs in AoU.

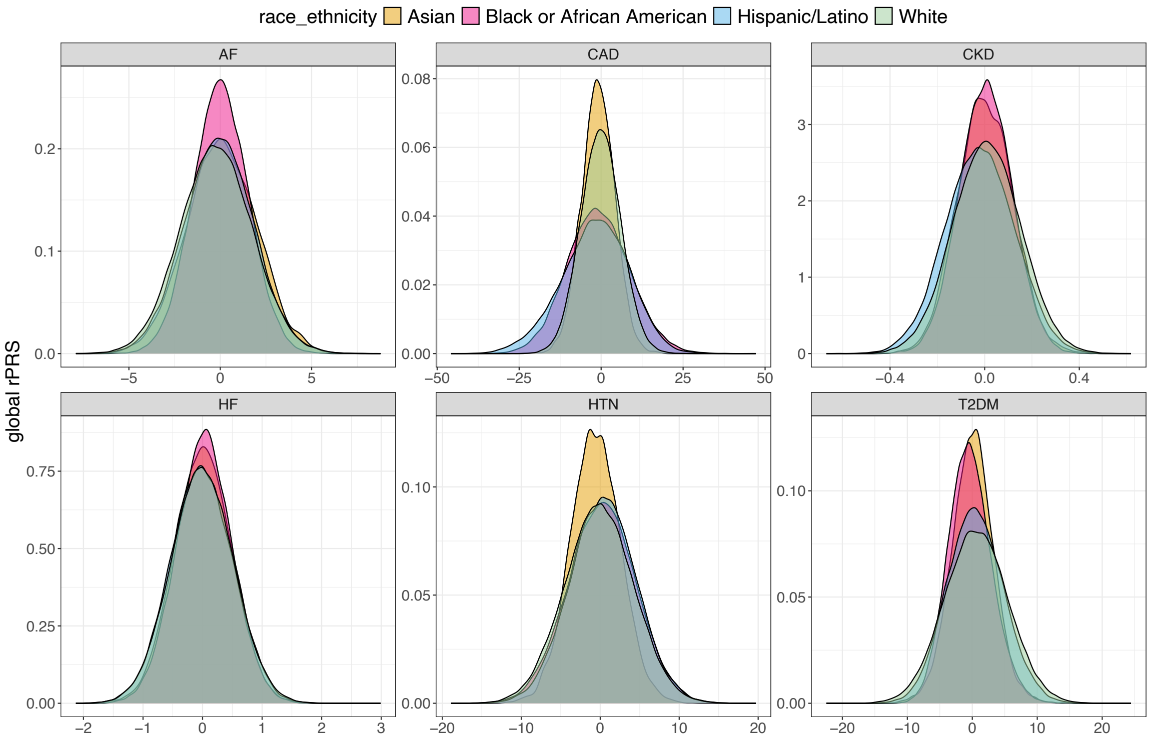

The figure visualizes global rPRS distributions in AoU participants, with rPRSs corresponding to AF, CAD, CKD, HF, HTN, and T2DM. Each rPRS was constructed as the difference between an individual's PRS and their global ePRS. Density plots are stratified by self-reported race/ethnicity groups.
Abbreviations: AoU: All of Us; rPRS: residual PRS; PRS: polygenic risk score; AF: atrial fibrillation; CAD: coronary artery disease; CKD: chronic kidney disease; HF: heart failure; HTN: hypertension; T2DM: type 2 diabetes mellitus.

#### Supplementary Figure 19: Density plots of PC-adjusted PRSs in AoU.

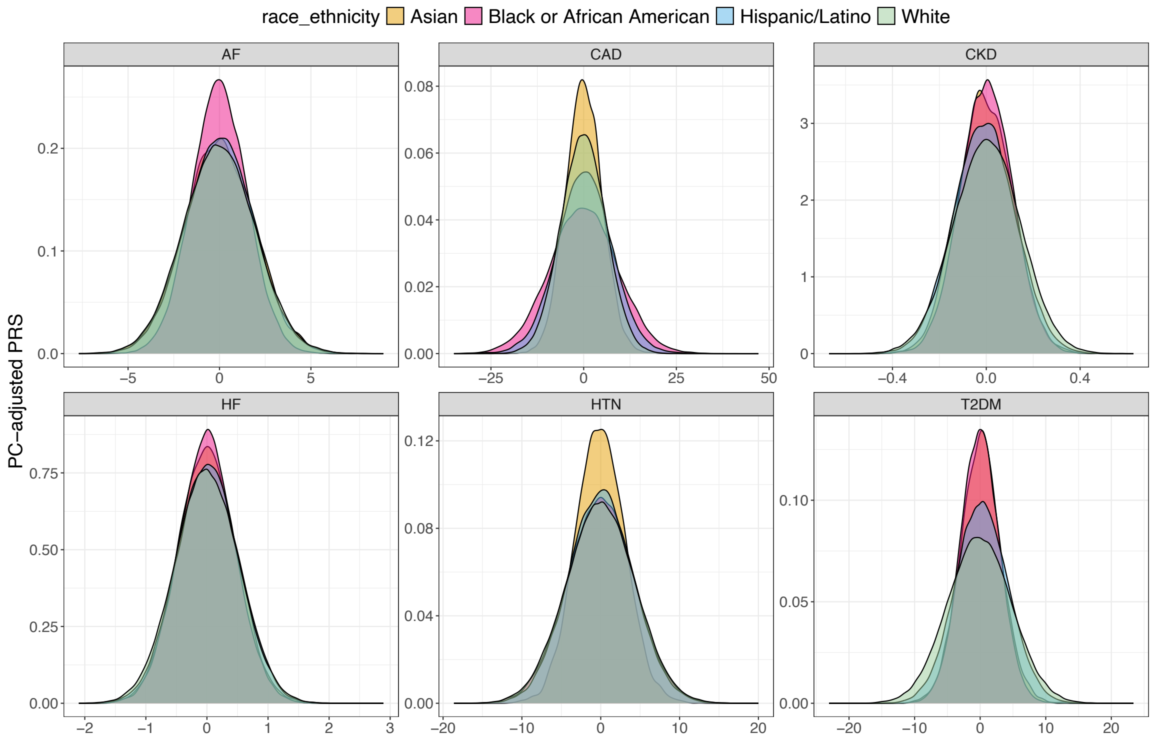

The figure visualizes distributions of PC-adjusted PRSs in AoU participants, with PRS corresponding to AF, CAD, CKD, HF, HTN, and T2DM. The PC-adjusted PRSs represent the residual values obtained from a model that regresses the PRS on genetic PCs. Density plots are stratified by self-reported race/ethnicity groups. Abbreviations: AoU: All of Us; PRS: polygenic risk score; AF: atrial fibrillation; CAD: coronary artery disease; CKD: chronic kidney disease; HF: heart failure; HTN: hypertension; T2DM: type 2 diabetes mellitus.

### Supplementary Figure 20: Associations between percentiles of PRS-related measurements and six CVD-related outcomes' proportions.

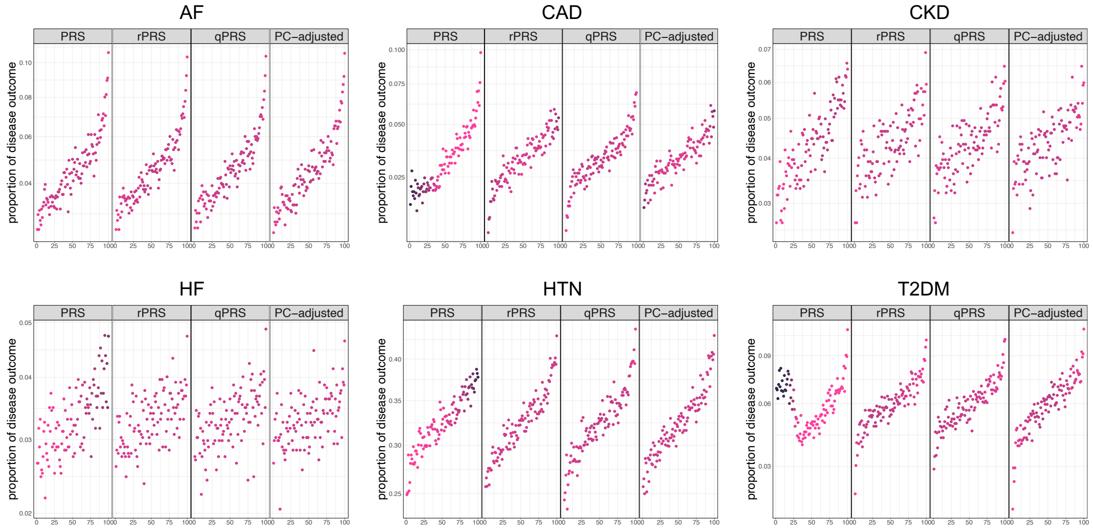

This figure illustrates the association between bins defined by percentiles of PRS-related measurements and the proportion of CVD-related outcomes. Each panel presents a scatterplot, with the x-axis representing the percentiles of PRS, rPRS, qPRS, and PC-adjusted PRS (from left to right), and the y-axis showing the proportion of the disease outcome. These proportions are computed for individuals within each corresponding PRS, rPRS, qPRS, or PC-adjusted PRS percentile. For example, for each phenotype and PRS measure, individuals were grouped into 100 strata based on their percentiles of PRS, rPRS, qPRS, or PC-adjusted PRS. The color of each point in the scatterplot represents the proportion of African American individuals in that percentile stratum. Darker colors indicate a higher proportion of African American individuals within the respective stratum. Abbreviations: AoU: All of Us; CVD: cardiovascular disease; PRS: polygenic risk score; rPRS: residual PRS; qPRS: quantile PRS; PC-adjusted: PC-adjusted PRS; AF: atrial fibrillation; CAD: coronary artery disease; CKD: chronic kidney disease; HF: heart failure; HTN: hypertension; T2DM: type 2 diabetes mellitus.

### Supplementary Note 6: Descriptions of TOPMed parent studies

#### Amish

**Ethics statement:**

All study protocols were approved by the institutional review board at the University of Maryland Baltimore. Informed consent was obtained from each study participant.

**Amish acknowledgements:**

We gratefully acknowledge our Amish liaisons, research volunteers, field workers, and Amish Research Clinic staff and the extraordinary cooperation and support of the Amish community without which these studies would not have been possible. The Amish studies are supported by grants and contracts from the NIH, including U01 HL072515, U01 HL84756, U01 HL137181, and P30 DK72488. The TOPMed component of the Amish Research Program was supported by NIH grants R01 HL121007, U01 HL072515, and R01 AG18728.

#### ARIC

The Atherosclerosis Risk in Communities study (dbGaP accession phs000090) is a population-based prospective cohort study of cardiovascular disease sponsored by the National Heart, Lung, and Blood Institute (NHLBI). ARIC included 15,792 individuals, predominantly European American and African American, aged 45-64 years at baseline (1987-89), chosen by probability sampling from four US communities. Cohort members completed three additional triennial follow-up examinations, a fifth exam in 2011-2013, a sixth exam in 2016-2017, a seventh exam in 2018-2019, an eighth exam in 2020, a ninth exam in 2021-2022, and tenth exam in 2023. The ARIC study has been described in detail previously (3,4).

**Ethics statement:**

The ARIC study has been approved by a single Institutional Review Board (sIRB) at Johns Hopkins School of Medicine and Institutional Review Boards (IRB) at all participating institutions: University of North Carolina at Chapel Hill IRB, Johns Hopkins University School of

Public Health IRB, University of Minnesota IRB, Wake Forest University Health Sciences IRB, and University of Mississippi Medical Center IRB. Study participants provided written informed consent at all study visits.

**ARIC acknowledgements:**

The Atherosclerosis Risk in Communities study has been funded in whole or in part with Federal funds from the National Heart, Lung, and Blood Institute, National Institutes of Health, Department of Health and Human Services (contract numbers 75N92022D00001, 75N92022D00002, 75N92022D00003, 75N92022D00004, 75N92022D00005). The authors thank the staff and participants of the ARIC study for their important contributions.

WGS for "NHLBI TOPMed: Atherosclerosis Risk in Communities (ARIC)" (phs001211) was performed at the Baylor College of Medicine Human Genome Sequencing Center (HHSN268201500015C and 3U54HG003273-12S2) and the Broad Institute for MIT and Harvard (3R01HL092577- 06S1). The Genome Sequencing Program (GSP) was funded by the National Human Genome Research Institute (NHGRI), the National Heart, Lung, and Blood Institute (NHLBI), and the National Eye Institute (NEI). The GSP Coordinating Center (U24 HG008956) contributed to cross program scientific initiatives and provided logistical and general study coordination. The Centers for Common Disease Genomics (CCDG) program was supported by NHGRI and NHLBI, and whole genome sequencing was performed at the Baylor College of Medicine Human Genome Sequencing Center (UM1 HG008898).

#### CARDIA

The Coronary Artery Risk Development in Young Adults study (dbGaP accession phs000285) is a prospective multicenter study with 5,115 adults Caucasian and African American participants of the age group 18-30 years at baseline, recruited from four centers at the baseline examination in 1985-1986 (5). The recruitment was done from the total community in Birmingham, AL, from selected census tracts in Chicago, IL and Minneapolis, MN; and from the Kaiser Permanente health plan membership in Oakland, CA. Nine examinations have been completed in the years

0, 2, 5, 7, 10, 15, 20, 25 and 30, with high retention rates (91%, 86%, 81%, 79%, 74%, 72%, 72%, and 71%, respectively) and written informed consent was obtained in each visit.

**Ethics statement:**

All CARDIA participants provided informed consent, and the study was approved by the Institutional Review Boards of the University of Alabama at Birmingham and the University of Texas Health Science Center at Houston.

**CARDIA acknowledgements:**

The Coronary Artery Risk Development in Young Adults Study (CARDIA) is conducted and supported by the National Heart, Lung, and Blood Institute (NHLBI) in collaboration with the University of Alabama at Birmingham (HHSN268201800005I & HHSN268201800007I), Northwestern University (HHSN268201800003I), University of Minnesota (HHSN268201800006I), and Kaiser Foundation Research Institute (HHSN268201800004I). CARDIA was also partially supported by the Intramural Research Program of the National Institute on Aging (NIA) and an intra-agency agreement between NIA and NHLBI (AG0005).

#### CFS

The Cleveland Family Study (CFS) was designed to examine the genetic basis of sleep apnea in 2,534 African-American and European-American individuals from 356 families. Index probands with confirmed sleep apnea were recruited from sleep centers in northern Ohio, supplemented with additional family members and neighborhood control families (6). Four visits occurred between 1990 and 2006; in the first 3, data were collected in participants' homes while the last occurred in a clinical research center (2000 - 2006). Measurements included sleep apnea monitoring, blood pressure, anthropometry, spirometry and other related phenotypes. Blood samples (overnight fasting, before bed and following an oral glucose tolerance test), nasal and oral ultrasound, and ECG were also obtained during the 4th exam. Institutional Review Board approval and signed informed consent was obtained for all participants.

**Ethics statement:**

Cleveland Family Study was approved by the Institutional Review Board (IRB) of Case Western Reserve University and Mass General Brigham (formerly Partners HealthCare). Written informed consent was obtained from all participants.

**CFS acknowledgements:**

The Cleveland Family Study has been supported in part by National Institutes of Health grants (R01-HL046380, KL2-RR024990, R35-HL135818, and R01-HL113338).

#### CHS

The Cardiovascular Health Study (CHS) is a population-based cohort study initiated by the National Heart, Lung and Blood Institute (NHLBI) in 1987 to determine the risk factors for development and progression of cardiovascular disease (CVD) in older adults, with an emphasis on subclinical measures. The study recruited 5,888 adults aged 65 or older at entry in four U.S. communities and conducted extensive annual clinical exams between 1989-1999 along with semi-annual phone calls, events adjudication, and subsequent data analyses and publications. Additional data are collected by studies ancillary to CHS. In June 1990, four Field Centers (Sacramento, CA; Hagerstown, MD; Winston-Salem, NC; Pittsburgh, PA) completed the recruitment of 5201 participants. Between November 1992 and June 1993, an additional 687 adults of primarily African Americans ethnicity were recruited using similar methods.

Blood samples were drawn from all participants at their baseline examination and during follow-up clinic visits and DNA was subsequently extracted from available samples. CHS analyses were limited to participants with available DNA who consented to genetic studies. The baseline examinations consisted of a home interview and a clinic examination that assessed not only traditional risk factors but also measures of subclinical disease, including carotid ultrasound, echocardiography, electrocardiography, and pulmonary function. Between enrollment and 1998-99, participants were seen in the clinic annually, and contacted by phone at 6-month intervals to collect information about hospitalizations and potential cardiovascular events. Major exam components were repeated during annual follow-up examinations through

1999. Cranial MRI scans, retinal photography, and tests of endothelial function were added as new components. Standard protocols for the identification and adjudication of events were implemented during follow-up. The adjudicated events are CHD, angina, heart failure (HF), stroke, transient ischemic attack (TIA), claudication and mortality. Adjudication of cause of death continues using a streamlined protocol; adjudication of other events ended in June 2015. Deep venous thrombosis and pulmonary embolism events from baseline through 2001 were adjudicated in an ancillary study: the Longitudinal Investigation of Thromboembolism Etiology (LITE). Since 1999, participants have been contacted every 6 months by phone, primarily to ascertain health status and for events follow-up. The study was initially approved by institutional review boards at the Field Centers (Wake Forest, University of California – Davis, Johns Hopkins University, University of Pittsburgh), the Core Laboratory (University of Vermont) and at the Coordinating Center (University of Washington). The University of Washington now handles CHS Data Repository approvals.

**Ethics statement:**

All CHS participants provided informed consent, and the study was approved by the Institutional Review Board [or ethics review committee] of University Washington.

**CHS acknowledgements:**

Cardiovascular Health Study: This research was supported by contracts HHSN268201200036C, HHSN268200800007C, HHSN268201800001C, N01HC55222, N01HC85079, N01HC85080, N01HC85081, N01HC85082, N01HC85083, N01HC85086, 75N92021D00006, and grants U01HL080295, U01HL130114, and HL105756 from the National Heart, Lung, and Blood Institute (NHLBI), with additional contribution from the National Institute of Neurological Disorders and Stroke (NINDS). Additional support was provided by R01AG023629 from the National Institute on Aging (NIA). A full list of principal CHS investigators and institutions can be found at CHS-NHLBI.org. The content is solely the responsibility of the authors and does not necessarily represent the official views of the National Institutes of Health.

#### COPDGene

COPDGene (7) is a cohort study for respiratory disease research, recruiting more than 10,000 subjects between the ages of 45 and 80 who had at least 10 pack-years of smoking during January 2008 - June 2011 at 21 clinical centers. Participants were characterized using spirometry, six-minute walk, inspiratory and expiratory chest CT scans, respiratory symptoms, medical history, medication history and 36-Item short form health survey. In the current analysis, we only used COPDGene control participants (meaning, individuals without COPD).

**Ethics statement:**

All COPDGene participants provided written informed consent, and the study was approved by the Institutional Review Boards of the participating clinical centers.

**COPDGene acknowledgements:**

The COPDGene project described was supported by Award Number U01 HL089897 and Award Number U01 HL089856 from the National Heart, Lung, and Blood Institute. The content is solely the responsibility of the authors and does not necessarily represent the official views of the National Heart, Lung, and Blood Institute or the National Institutes of Health. The COPDGene project is also supported by the COPD Foundation through contributions made to an Industry Advisory Board comprised of AstraZeneca, Boehringer Ingelheim, GlaxoSmithKline, Novartis, Pfizer, Siemens and Sunovion. A full listing of COPDGene investigators can be found at: http://www.copdgene.org/directory

#### FHS

The Framingham Heart Study (dbGaP accession phs000007) began in 1948 with the recruitment of an original cohort of 5,209 men and women (mean age 44 years; 55 percent women). In 1971 a second generation of study participants was enrolled; this cohort (mean age 37 years; 52% women) consisted of 5,124 children and spouses of children of the original cohort. A third-generation cohort of 4,095 children of offspring cohort participants (mean age 40 years; 53 percent women) was enrolled in 2002-2005 and are seen every 4 to 8 years. Details of study designs for the three cohorts are summarized elsewhere (8–10). At each clinic visit, a medical history was obtained, and participants underwent a physical examination. Only study participants consented for genetic and non-genetic data are included. FHS has been approved by the Boston University IRB

**Ethics statement:**

The Framingham Heart Study was approved by the Institutional Review Board of the Boston University Medical Center. All study participants provided written informed consent.

**FHS acknowledgements:**

The Framingham Heart Study (FHS) acknowledges the support of contracts NO1-HC-25195, HHSN268201500001I and 75N92019D00031 from the National Heart, Lung and Blood Institute and grant supplement R01 HL092577-06S1 for this research. We also acknowledge the dedication of the FHS study participants without whom this research would not be possible. Dr. Vasan is supported in part by the Evans Medical Foundation and the Jay and Louis Coffman Endowment from the Department of Medicine, Boston University School of Medicine.

#### GENOA

The Genetic Epidemiology Network of Arteriopathy (GENOA) study (dbGaP accession phs000379), a part of the Family Blood Pressure Program (FBPP Investigators, 2002), consists of hypertensive sibships that were recruited for linkage and association studies in order to identify genes that influence blood pressure and its target organ damage (Daniels, 2004). In the initial phase of the GENOA study (Phase I: 1996-2001), all members of sibships containing ≥ 2 individuals with essential hypertension clinically diagnosed before age 60 were invited to participate, including both hypertensive and normotensive siblings. In the second phase of the GENOA study (Phase II: 2000-2004), 1,239 non-Hispanic white and 1,482 African American participants were successfully re-recruited to measure potential target organ damage due to hypertension.

**Ethics statement:**

Written informed consent was obtained from all subjects and approval was granted by participating institutional review boards (University of Michigan, University of Mississippi Medical Center, and Mayo Clinic).

**GENOA acknowledgements:**

Support for the Genetic Epidemiology Network of Arteriopathy (GENOA) was provided by the National Heart, Lung and Blood Institute (U01 HL054457, U01 HL054464, U01 HL054481, R01 HL119443, and R01 HL087660) of the National Institutes of Health. DNA extraction for "NHLBI TOPMed: Genetic Epidemiology Network of Arteriopathy" (phs001345) was performed at the Mayo Clinic Genotyping Core, and WGS was performed at the DNA Sequencing and Gene Analysis Center at the University of Washington (3R01HL055673-18S1) and the Broad Institute (HHSN268201500014C). We would like to thank the GENOA participants.

#### HCHS/SOL

The Hispanic Community Health Study/Study of Latinos (dbGaP accession phs000810) is a community-based longitudinal cohort study of 16,415 self-identified Hispanic/Latino persons aged 18–74 years and selected from households in predefined census-block groups across four US field centers (in Chicago, Miami, the Bronx, and San Diego). The census-block groups were chosen to provide diversity among cohort participants with regard to socioeconomic status and national origin or background (11,12). The HCHS/SOL cohort includes participants who self-identified as having a Hispanic/Latino background; the largest groups are Central American (n = 1,730), Cuban (n = 2,348), Dominican (n = 1,460), Mexican (n = 6,471), Puerto Rican (n = 2,728), and South American (n = 1,068). The HCHS/SOL baseline clinical examination occurred between 2008 and 2011 and included comprehensive biological, behavioral, and sociodemographic assessments. Visit 2 took place between 2014 and 2017, which re-examined 11,623 participants from the baseline sample. Visit 3 has started in 2020 and will last 4 years, ending on January 2024. In addition to clinic visit, participants are contacted annually to assess clinical outcomes.

The study was approved by the Institutional Review Boards at each participating institution and written informed consent was obtained from all participants.

**Ethics statement:**

This study was approved by the institutional review boards (IRBs) at each field center, where all participants gave written informed consent, and by the Non-Biomedical IRB at the University of North Carolina at Chapel Hill, to the HCHS/SOL Data Coordinating Center. All IRBs approving the study are: Non-Biomedical IRB at the University of North Carolina at Chapel Hill. Chapel Hill, NC; Einstein IRB at the Albert Einstein College of Medicine of Yeshiva University. Bronx, NY; IRB at Office for the Protection of Research Subjects (OPRS), University of Illinois at Chicago. Chicago, IL; Human Subject Research Office, University of Miami. Miami, FL; Institutional Review Board of San Diego State University. San Diego, CA.

**HCHS/SOL acknowledgements:**

The Hispanic Community Health Study/Study of Latinos is a collaborative study supported by contracts from the National Heart, Lung, and Blood Institute (NHLBI) to the University of North Carolina (HHSN268201300001I / N01-HC-65233), University of Miami (HHSN268201300004I / N01-HC- 65234), Albert Einstein College of Medicine (HHSN268201300002I / N01-HC-65235), University of Illinois at Chicago – HHSN268201300003I / N01- HC-65236 Northwestern Univ), and San Diego State University (HHSN268201300005I / N01-HC-65237). The following Institutes/Centers/Offices have contributed to the HCHS/SOL through a transfer of funds to the NHLBI: National Institute on Minority Health and Health Disparities, National Institute on Deafness and Other Communication Disorders, National Institute of Dental and Craniofacial Research, National Institute of Diabetes and Digestive and Kidney Diseases, National Institute of Neurological Disorders and Stroke, NIH Institution-Office of Dietary Supplements.

#### HVH

The Heart and Vascular Health (HVH) VTE Study (dbGaP accession phs000993) is a case-control study of risk factors for cardiovascular outcomes set at Group Health (GH), an integrated health care delivery system in western Washington State. Cases include venous thromboembolism (VTE), myocardial infarction (MI), stroke, and atrial fibrillation (AF), with a shared common control group frequency matched to MI cases on age (within decade) sex, treated hypertension, and calendar year of identification. Study approval was granted by the human subjects committee at GH, and written informed consent was provided by all study participants. Eligibility and risk factor information were collected by trained medical record abstractors from a review of the GH medical record using only data available prior to the event date of cases and a randomly selected date for the controls. All VTE, MI, stroke and AF events were verified by medical record review. For the TOPMed data set, only incident idiopathic cases of VT and early-onset (age <=60 years) cases of AF without underlying heart failure, myocardial infarction, or valvular heart disease were included. Within the HVH study, VT and AF cases were diagnosed in both inpatient and outpatient settings. A venous blood sample was collected from all consenting subjects, and DNA was extracted from white blood cells using standard procedures.

**Ethics statement:**

Study approval was granted by the human subjects committee at Group Health, and written informed consent was provided by all study participants.

**HVH acknowledgements:**

The Heart and Vascular Health Study was supported by grants HL068986, HL085251, HL095080, and HL073410 from the National Heart, Lung, and Blood Institute.

#### JHS

The Jackson Heart Study (dbGaP accession phs000286) is a longitudinal investigation of genetic and environmental risk factors associated with the disproportionate burden of cardiovascular disease in African Americans (13,14). At baseline, the JHS recruited 5306 African American residents of the Jackson, Mississippi Metropolitan Statistical Area, which included approximately 6.6% of all African American adults aged 35-84 residing in the area. Participants were recruited via random sampling (17% of participants), volunteers (30%), prior participants in the Atherosclerosis Risk in Communities (ARIC) study (31%), and secondary family members (22%). Among these participants, approximately 3400 gave consent that allows genetic research. JHS participants received three back-to-back clinical examinations (Exam 1, 2000-2004; Exam 2, 2005-2008; and Exam 3, 2009-2013), and a fourth clinical examination started in 2020. Participants are also contacted annually by telephone to update personal and health information including vital status, interim medical events, hospitalizations, functional status and sociocultural information.

**Ethics statement:**

The Institutional Review Boards at Jackson State University, Tougaloo College, and the University of Mississippi Medical Center approved the study, and all participants provided written informed consent.

**JHS acknowledgements:**

The Jackson Heart Study (JHS) is supported and conducted in collaboration with Jackson State University (HHSN268201800013I), Tougaloo College (HHSN268201800014I), the Mississippi State Department of Health (HHSN268201800015I) and the University of Mississippi Medical Center (HHSN268201800010I, HHSN268201800011I and HHSN268201800012I) contracts from the National Heart, Lung, and Blood Institute (NHLBI) and the National Institute for Minority Health and Health Disparities (NIMHD). The authors also wish to thank the staffs and participants of the JHS.

Genome sequencing (dbGap accession phs000964) was performed at the Northwest Genomics Center (HHSN268201100037C). Core support including centralized genomic read mapping and genotype calling, along with variant quality metrics and filtering were provided by the TOPMed Informatics Research Center (R01HL117626; contract HHSN268201800002I). Core support including phenotype harmonization, data management, sample-identity QC, and general program coordination were provided by the TOPMed Data Coordinating Center (R01HL120393; U01HL120393; contract HHSN268201800001I).

#### Mayo VTE

Study description from dbGaP https://www.ncbi.nlm.nih.gov/projects/gap/cgi-bin/study.cgi?study_id=phs001402.v3.p1:

This study consists of 338 VTE cases from an inception cohort of Olmsted County, MN residents (OC) with a first lifetime objectively-diagnosed idiopathic VTE during the 40-year study period, 1966-2005. All living study subjects were invited to provide a whole blood sample at the Mayo Clinical Research Unit for leukocyte genomic DNA and plasma collection. For living study subjects who did not provide a blood sample, we retrieved any leftover blood ("waste" blood) from samples collected as part of routine clinical diagnostic testing and used this to extract DNA after obtaining patient consent. For deceased cases, with IRB approval, we extracted DNA from any available stored tissue within the Mayo Tissue Archive. This "tissue" DNA has been successfully genotyped in prior studies. Three trained and experienced study nurse abstractors reviewed the complete medical records in the community of all potential cases.

**Ethics statements:**

All Mayo-VTE participants provided informed consent and the study was approved by the Institutional Review Board of Mayo Clinic, Rochester, MN.

**Mayo-VTE acknowledgements:**

Funded, in part, by grants from the National Institutes of Health, National Heart, Lung and Blood Institute (HL66216 and HL83141). the National Human Genome Research Institute (HG04735, HG06379), and research support provided by Mayo Foundation.

#### MESA

The Multi-Ethnic Study of Atherosclerosis (dbGaP accession phs000209) is a study of the characteristics of subclinical cardiovascular disease (disease detected non-invasively before it has produced clinical signs and symptoms) and the risk factors that predict progression to clinically overt cardiovascular disease or progression of the subclinical disease (15). MESA consisted of a diverse, population-based sample of an initial 6,814 asymptomatic men and women aged 45-84. 38 percent of the recruited participants were white, 28 percent African American, 22 percent Hispanic, and 12 percent Asian, predominantly of Chinese descent. Participants were recruited from six field centers across the United States: Wake Forest University, Columbia University, Johns Hopkins University, University of Minnesota, Northwestern University and University of California - Los Angeles. Participants are being followed for identification and characterization of cardiovascular disease events, including acute myocardial infarction and other forms of coronary heart disease (CHD), stroke, and congestive heart failure; for cardiovascular disease interventions; and for mortality. The first examination took place over two years, from July 2000 - July 2002. It was followed by five examination periods that were 17-20 months in length. Participants have been contacted every 9 to 12 months throughout the study to assess clinical morbidity and mortality.

**Ethics statements:**

All MESA participants provided written informed consent, and the study was approved by the Institutional Review Boards at The Lundquist Institute (formerly Los Angeles BioMedical Research Institute) at Harbor-UCLA Medical Center, University of Washington, Wake Forest School of Medicine, Northwestern University, University of Minnesota, Columbia University, and Johns Hopkins University.

**MESA acknowledgements:**

MESA and the MESA SHARe project are conducted and supported by the National Heart, Lung, and Blood Institute (NHLBI) in collaboration with MESA investigators. Support for MESA is provided by contracts HHSN268201500003I, N01-HC-95159, N01-HC-95160, N01-HC-95161, N01-HC-95162, N01-HC-95163, N01-HC-95164, N01-HC-95165, N01-HC-95166, N01-HC-95167,

N01-HC-95168, N01-HC-95169, UL1-TR-000040, UL1-TR-001079, UL1-TR-001420. MESA Family is conducted and supported by the National Heart, Lung, and Blood Institute (NHLBI) in collaboration with MESA investigators. Support is provided by grants and contracts R01HL071051, R01HL071205, R01HL071250, R01HL071251, R01HL071258, R01HL071259, and by the National Center for Research Resources, Grant UL1RR033176. The provision of genotyping data was supported in part by the National Center for Advancing Translational Sciences, CTSI grant UL1TR001881, and the National Institute of Diabetes and Digestive and Kidney Disease Diabetes Research Center (DRC) grant DK063491 to the Southern California Diabetes Endocrinology Research Center.

#### WHI

The Women's Health Initiative (WHI) cohort. The WHI is a prospective national health study focused on identifying optimal strategies for preventing chronic diseases that are the major causes of death and disability in postmenopausal women. The WHI initially recruited 161,808 women between 1993 and 1997 with the goal of including a socio-demographically diverse population with diversity background groups proportionate to the total minority population of US women aged 50-79 years. The WHI consists of two major parts: a set of randomized Clinical Trials and an Observational Study. The WHI Clinical Trials (CT; N=68,132) includes three overlapping components, each a randomized controlled comparison: the Hormone Therapy Trials (HT), Dietary Modification Trial, and Calcium and Vitamin D Trial. A parallel prospective observational study (OS; N = 93,676) examined biomarkers and risk factors associated with various chronic diseases. While the HT trials ended in the mid-2000s, active follow-up of the WHI-CT and WHI-OS cohorts has continued for over 25 years, with the accumulation of large numbers of diverse clinical outcomes, risk factor measurements, medication use, and many other types of data.

**Ethics statement:**

All WHI participants provided informed consent and the study was approved by the Institutional Review Board (IRB) of the Fred Hutchinson Cancer Research Center.

**WHI acknowledgements:**

The WHI program is funded by the National Heart, Lung, and Blood Institute, National Institutes of Health, U.S. Department of Health and Human Services through contracts 75N92021D00001, 75N92021D00002, 75N92021D00003, 75N92021D00004, 75N92021D00005.

#### Supplementary Note 7: TOPMed and CCDG acknowledgements

Molecular data for the Trans-Omics in Precision Medicine (TOPMed) program was supported by the National Heart, Lung and Blood Institute (NHLBI). Genome sequencing for "NHLBI TOPMed: Genetics of Cardiometabolic Health in the Old Order Amish Study" (phs000956) were performed at the Broad Institute of MIT and Harvard (HHSN268201500014C). Genome sequencing for "NHLBI TOPMed: Whole Genome Sequencing and Related Phenotypes in the Framingham Heart Study" (phs000974.v4.p3) was performed at the Broad Institute Genomics Platform (3R01HL092577-06S1, 3U54HG003067-12S2). Genome sequencing for the "NHLBI TOPMed: Genetic Epidemiology Network of Arteriopathy (GENOA)" (phs001345.v2.p1) was performed at the Broad Institute Genomics Platform (HHSN268201500014C) and the Northwest Genomics Center (3R01HL055673-18S1). Genome sequencing for "NHLBI TOPMed: The Jackson Heart Study" (phs000964.v1.p1) was performed at the Northwest Genomics Center (HHSN268201100037C). Genome sequencing for the "NHLBI TOPMed: The Atherosclerosis Risk in Communities Study" (phs001211.v3.p2) was performed at the Baylor College of Medicine Human Genome Sequencing Center (HHSN268201500015C and 3U54HG003273-12S2) and the Broad Institute for MIT and Harvard (3R01HL092577- 06S1). Genome sequencing for "NHLBI TOPMed: Coronary Artery Risk Development in Young Adults Study" (phs001612.v1.p1) was performed at the Baylor College of Medicine Human Genome Sequencing Center (HHSN268201600033I). Genome sequencing for “NHLBI TOPMed: Cleveland Family Study” (phs000954.v3.p2) was performed at the Northwest Genomics Center (3R01HL098433-05S1, HHSN268201600032I). Genome sequencing for “NHLBI TOPMed: Genetic Epidemiology of COPD (COPDGene) in the TOPMed Program” (phs000951) was performed at the University of Washington Northwest Genomics Center (3R01 HL089856-08S1) and the Broad Institute of MIT and Harvard (HHSN268201500014C). Genomics sequencing for “NHLBI TOPMed: Cardiovascular Health Study” (phs001368.v2.p1) was performed at the Baylor College of Medicine Human Genome Sequencing Center (3U54HG003273-12S2, HHSN268201500015C, HHSN268201600033I). Genome sequencing for “NHLBI TOPMed: Hispanic Community Health Study/Study of Latinos” (phs001395.v1.p1) was performed at the Baylor College of Medicine Human Genome Sequencing Center (HHSN268201600033I). Genome sequencing for “NHLBI TOPMed: Heart and Vascular Health Study (HVH)” (phs000993.v5.p2) was performed at the Baylor College of Medicine Human Genome Sequencing Center (3U54HG003273- 12S2, HHSN26820150001 5C). Genome sequencing for “NHLBI TOPMed: Women’s Health Initiative (WHI)” (phs001237.v2.p1) was performed at the Broad Institute of MIT and Harvard (HHSN268201500014C). Genome sequencing for “NHLBI TOPMed: Multi-Ethnic Study of Atherosclerosis” (phs001416.v2.p1) was performed at Broad Institute Genomics Platform (HHSN268201500014C, 3U54HG003067-13S1). Genome sequencing for “NHLBI TOPMed: Mayo Clinic Venous Thromboembolism Study (Mayo_VTE)” (phs001402.v3.p1) was performed at the Baylor College of Medicine Human Genome Sequencing Center (3U54HG003273-12S2, HHSN268201500015C). Core support including centralized genomic read mapping and genotype calling, along with variant quality metrics and filtering were provided by the TOPMed Informatics Research Center (3R01HL-117626-02S1; contract HHSN268201800002I). Core support including phenotype harmonization, data management, sample-identity QC, and general program coordination were provided by the TOPMed Data Coordinating Center (R01HL-120393; U01HL-120393; contract HHSN268201800001I). We gratefully acknowledge the studies and participants who provided biological samples and data for TOPMed. The Genome Sequencing Program (GSP) was funded by the National Human Genome Research Institute (NHGRI), the National Heart, Lung, and Blood Institute (NHLBI), and the National Eye Institute (NEI). The GSP Coordinating Center (U24 HG008956) contributed to cross-program scientific initiatives and provided logistical and general study coordination. The Centers for Common Disease Genomics (CCDG) program was supported by NHGRI and NHLBI, and whole genome sequencing was performed at the Baylor College of Medicine Human Genome Sequencing Center (UM1 HG008898).

#### Supplementary Note 8: TOPMed consortium investigators

Namiko Abe[1], Gonçalo Abecasis[2], Francois Aguet[3], Christine Albert[4], Laura Almasy[5], Alvaro Alonso[6], Seth Ament[7], Peter Anderson[8], Pramod Anugu[9], Deborah Applebaum-Bowden[10], Kristin Ardlie[3], Dan Arking[11], Donna K Arnett[12], Allison Ashley-Koch[13], Stella Aslibekyan[14], Tim Assimes[15], Paul Auer[16], Dimitrios Avramopoulos[11], Najib Ayas[17], Adithya Balasubramanian[18], John Barnard[19], Kathleen Barnes[20], R. Graham Barr[21], Emily Barron-Casella[11], Lucas Barwick[22], Terri Beaty[11], Gerald Beck[23], Diane Becker[24], Lewis Becker[11], Rebecca Beer[25], Amber Beitelshees[7], Emelia Benjamin[26], Takis Benos[27], Marcos Bezerra[28], Larry Bielak[2], Joshua Bis[29],

Thomas Blackwell[2], John Blangero[30], Eric Boerwinkle[31], Donald W. Bowden[32], Russell Bowler[33], Jennifer Brody[8], Ulrich Broeckel[34], Jai Broome[8], Deborah Brown[35], Karen Bunting[1], Esteban Burchard[36], Carlos Bustamante[37], Erin Buth[38], Brian Cade[39], Jonathan Cardwell[40], Vincent Carey[41], Julie Carrier[42], April Carson[43], Cara Carty[44], Richard Casaburi[45], Juan P Casas Romero[46], James Casella[11], Peter Castaldi[47], Mark Chaffin[3], Christy Chang[7], Yi-Cheng Chang[48], Daniel Chasman[49], Sameer Chavan[40], Bo-Juen Chen[1], Wei-Min Chen[50], Yii-Der Ida Chen[51], Michael Cho[41], Seung Hoan Choi[3], Lee-Ming Chuang[52], Mina Chung[53], Ren-Hua Chung[54], Clary Clish[55], Suzy Comhair[56], Matthew Conomos[38], Elaine Cornell[57], Adolfo Correa[58], Carolyn Crandall[45], James Crapo[59], L. Adrienne Cupples[60], Joanne Curran[61], Jeffrey Curtis[62], Brian Custer[63], Coleen Damcott[7], Dawood Darbar[64], Sean David[65], Colleen Davis[8], Michelle Daya[40], Mariza de Andrade[66], Lisa de las Fuentes[67], Paul de Vries[68], Michael DeBaun[69], Ranjan Deka[70], Dawn DeMeo[41], Scott Devine[7], Huyen Dinh[18], Harsha Doddapaneni[18], Qing Duan[71], Shannon Dugan-Perez[18], Ravi Duggirala[72], Jon Peter Durda[57], Susan K. Dutcher[73], Charles Eaton[74], Lynette Ekunwe[9], Adel El Boueiz[75], Patrick Ellinor[76], Leslie Emery[8], Serpil Erzurum[19], Charles Farber[50], Jesse Farek[18], Tasha Fingerlin[77], Matthew Flickinger[2], Myriam Fornage[31], Nora Franceschini[78], Chris Frazar[8], Mao Fu[7], Stephanie M. Fullerton[8], Lucinda Fulton[79], Stacey Gabriel[3], Weiniu Gan[25], Shanshan Gao[40], Yan Gao[9], Margery Gass[80], Heather Geiger[81], Bruce Gelb[82], Mark Geraci[27], Soren Germer[1], Robert Gerszten[83], Auyon Ghosh[41], Richard Gibbs[18], Chris Gignoux[15], Mark Gladwin[27], David Glahn[84], Stephanie Gogarten[8], Da-Wei Gong[7], Harald Goring[85], Sharon Graw[86], Kathryn J. Gray[87], Daniel Grine[40], Colin Gross[2], C. Charles Gu[79], Yue Guan[7], Xiuqing Guo[51], Namrata Gupta[3], David M. Haas[88], Jeff Haessler[80], Michael Hall[89], Yi Han[18], Patrick Hanly[90], Daniel Harris[91], Nicola L. Hawley[92], Jiang He[93], Ben Heavner[38], Susan Heckbert[94], Ryan Hernandez[36], David Herrington[95], Craig Hersh[96], Bertha Hidalgo[14], James Hixson[31], Brian Hobbs[41], John Hokanson[40], Elliott Hong[7], Karin Hoth[97], Chao (Agnes) Hsiung[98], Jianhong Hu[18], Yi-Jen Hung[99], Haley Huston[100], Chii Min Hwu[101], Marguerite Ryan Irvin[14], Rebecca Jackson[102], Deepti Jain[8], Cashell Jaquish[103], Jill Johnsen[104], Andrew Johnson[25], Craig Johnson[8], Rich Johnston[6], Kimberly Jones[11], Hyun Min Kang[105], Robert Kaplan[106], Sharon Kardia[2], Shannon Kelly[36], Eimear Kenny[82], Michael Kessler[7], Alyna Khan[8], Ziad Khan[18], Wonji Kim[107], John Kimoff[108], Greg Kinney[109], Barbara Konkle[110], Charles Kooperberg[80], Holly Kramer[111], Christoph

Lange[112], Ethan Lange[40], Leslie Lange[113], Cathy Laurie[8], Cecelia Laurie[8], Meryl LeBoff[41], Jiwon Lee[41], Sandra Lee[18], Wen-Jane Lee[101], Jonathon LeFaive[2], David Levine[8], Dan Levy[25], Joshua Lewis[7], Xiaohui Li[51], Yun Li[71], Henry Lin[51], Honghuang Lin[114], Xihong Lin[115], Simin Liu[116], Yongmei Liu[117], Yu Liu[118], Ruth J.F. Loos[119], Steven Lubitz[76], Kathryn Lunetta[114], James Luo[25], Ulysses Magalang[120], Michael Mahaney[61], Barry Make[11], Ani Manichaikul[50], Alisa Manning[121], JoAnn Manson[41], Lisa Martin[122], Melissa Marton[81], Susan Mathai[40], Rasika Mathias[11], Susanne May[38], Patrick McArdle[7], Merry-Lynn McDonald[123], Sean McFarland[107], Stephen McGarvey[124], Daniel McGoldrick[125], Caitlin McHugh[38], Becky McNeil[126], Hao Mei[9], James Meigs[127], Vipin Menon[18], Luisa Mestroni[86], Ginger Metcalf[18], Deborah A Meyers[128], Emmanuel Mignot[129], Julie Mikulla[25], Nancy Min[9], Mollie Minear[130], Ryan L Minster[27], Braxton D. Mitchell[7], Matt Moll[47], Zeineen Momin[18], May E. Montasser[7], Courtney Montgomery[131], Donna Muzny[18], Josyf C Mychaleckyj[50], Girish Nadkarni[82], Rakhi Naik[11], Take Naseri[132], Pradeep Natarajan[3], Sergei Nekhai[133], Sarah C. Nelson[38], Bonnie Neltner[40], Caitlin Nessner[18], Deborah Nickerson[134], Osuji Nkechinyere[18], Kari North[71], Jeff O'Connell[135], Tim O'Connor[7], Heather Ochs-Balcom[136], Geoffrey Okwuonu[18], Allan Pack[137], David T. Paik[138], Nicholette Palmer[139], James Pankow[140], George Papanicolaou[25], Cora Parker[141], Gina Peloso[142], Juan Manuel Peralta[72], Marco Perez[15], James Perry[7], Ulrike Peters[143], Patricia Peyser[2], Lawrence S Phillips[6], Jacob Pleiness[2], Toni Pollin[7], Wendy Post[144], Julia Powers Becker[145], Meher Preethi Boorgula[40], Michael Preuss[82], Bruce Psaty[8], Pankaj Qasba[25], Dandi Qiao[41], Zhaohui Qin[6], Nicholas Rafaels[146], Laura Raffield[147], Mahitha Rajendran[18], Vasan S. Ramachandran[114], D.C. Rao[79], Laura Rasmussen-Torvik[148], Aakrosh Ratan[50], Susan Redline[47], Robert Reed[7], Catherine Reeves[149], Elizabeth Regan[59], Alex Reiner[150], Muagututi'a Sefuiva Reupena[151], Ken Rice[8], Stephen Rich[50], Rebecca Robillard[152], Nicolas Robine[81], Dan Roden[153], Carolina Roselli[3], Jerome Rotter[154], Ingo Ruczinski[11], Alexi Runnels[81], Pamela Russell[40], Sarah Ruuska[100], Kathleen Ryan[7], Ester Cerdeira Sabino[155], Danish Saleheen[21], Shabnam Salimi[156], Sejal Salvi[18], Steven Salzberg[11], Kevin Sandow[157], Vijay G. Sankaran[158], Jireh Santibanez[18], Karen Schwander[79], David Schwartz[40], Frank Sciurba[27], Christine Seidman[159], Jonathan Seidman[160], Frédéric Sériès[161], Vivien Sheehan[162], Stephanie L. Sherman[163], Amol Shetty[7], Aniket Shetty[40], Wayne Hui- Heng Sheu[101], M. Benjamin Shoemaker[164], Brian Silver[165], Edwin Silverman[41], Robert Skomro[166], Albert Vernon Smith[167], Jennifer Smith[2], Josh Smith[8], Nicholas Smith[94], Tanja

Smith[1], Sylvia Smoller[106], Beverly Snively[168], Michael Snyder[15], Tamar Sofer[41], Nona Sotoodehnia[8], Adrienne M. Stilp[8], Garrett Storm[169], Elizabeth Streeten[7], Jessica Lasky Su[170], Yun Ju Sung[79], Jody Sylvia[41], Adam Szpiro[8], Daniel Taliun[2], Hua Tang[171], Margaret Taub[11], Kent D. Taylor[172], Matthew Taylor[86], Simeon Taylor[7], Marilyn Telen[13], Timothy A. Thornton[8], Machiko Threlkeld[173], Lesley Tinker[174], David Tirschwell[8], Sarah Tishkoff[175], Hemant Tiwari[176], Catherine Tong[177], Russell Tracy[178], Michael Tsai[140], Dhananjay Vaidya[11], David Van Den Berg[179], Peter VandeHaar[2], Scott Vrieze[140], Tarik Walker[40], Robert Wallace[97], Avram Walts[40], Fei Fei Wang[8], Heming Wang[180], Jiongming Wang[167], Karol Watson[45], Jennifer Watt[18], Daniel E. Weeks[27], Joshua Weinstock[105], Bruce Weir[8], Scott T Weiss[181], Lu-Chen Weng[76], Jennifer Wessel[182], Cristen Willer[62], Kayleen Williams[38], L. Keoki Williams[183], Carla Wilson[41], James Wilson[184], Lara Winterkorn[81], Quenna Wong[8], Joseph Wu[138], Huichun Xu[7], Lisa Yanek[11], Ivana Yang[40], Ketian Yu[2], Seyedeh Maryam Zekavat[3], Yingze Zhang[185], Snow Xueyan Zhao[59], Wei Zhao[186], Xiaofeng Zhu[187], Michael Zody[1], Sebastian Zoellner[2]

1 - New York Genome Center, New York, New York; 2 - University of Michigan, Ann Arbor, Michigan; 3 - Broad Institute, Cambridge, Massachusetts; 4 - Cedars Sinai, Boston, Massachusetts; 5 - Children's Hospital of Philadelphia, University of Pennsylvania, Philadelphia, Pennsylvania; 6 - Emory University, Atlanta, Georgia; 7 - University of Maryland, Baltimore, Maryland; 8 - University of Washington, Seattle, Washington; 9 - University of Mississippi, Jackson, Mississippi; 10 - National Institutes of Health, Bethesda, Maryland; 11 - Johns Hopkins University, Baltimore, Maryland; 12 - University of Kentucky, Lexington, Kentucky; 13 - Duke University, Durham, North Carolina; 14 - University of Alabama, Birmingham, Alabama; 15 - Stanford University, Stanford, California; 16 - Medical College of Wisconsin, Milwaukee, Wisconsin; 17 - Medicine, Providence Health Care, Vancouver; 18 - Baylor College of Medicine Human Genome Sequencing Center, Houston, Texas; 19 - Cleveland Clinic, Cleveland, Ohio; 20 - Tempus, University of Colorado Anschutz Medical Campus, Aurora, Colorado; 21 - Columbia University, New York, New York; 22 - LTRC, The Emmes Corporation, Rockville, Maryland; 23 - Quantitative Health Sciences, Cleveland Clinic, Cleveland, Ohio; 24 - Medicine, Johns Hopkins University, Baltimore, Maryland; 25 - National Heart, Lung, and Blood Institute, National

Institutes of Health, Bethesda, Maryland; 26 - Boston University School of Medicine, Boston University, Massachusetts General Hospital, Boston, Massachusetts; 27 - University of Pittsburgh, Pittsburgh, Pennsylvania; 28 - Fundação de Hematologia e Hemoterapia de Pernambuco - Hemope, Recife; 29 - Cardiovascular Health Research Unit, Department of Medicine, University of Washington, Seattle, Washington; 30 - Human Genetics, University of Texas Rio Grande Valley School of Medicine, Brownsville, Texas; 31 - University of Texas Health at Houston, Houston, Texas; 32 - Department of Biochemistry, Wake Forest Baptist Health, Winston-Salem, North Carolina; 33 - National Jewish Health, National Jewish Health, Denver, Colorado; 34 - Pediatrics, Medical College of Wisconsin, Milwaukee, Wisconsin; 35 - Pediatrics, University of Texas Health at Houston, Houston, Texas; 36 - University of California, San Francisco, San Francisco, California; 37 – Biomedical Data Science, Stanford University, Stanford, California; 38 - Biostatistics, University of Washington, Seattle, Washington; 39 - Brigham and Women's Hospital, Brigham & Women's Hospital, Boston, Massachusetts; 40 - University of Colorado at Denver, Denver, Colorado; 41 - Brigham & Women's Hospital, Boston, Massachusetts; 42 - University of Montreal; 43 - Medicine, University of Mississippi, Jackson, Mississippi; 44 - Washington State University, Pullman, Washington; 45 - University of California, Los Angeles, Los Angeles, California; 46 - Brigham & Women's Hospital; 47 - Medicine, Brigham & Women's Hospital, Boston, Massachusetts; 48 - National Taiwan University, Taipei; 49 - Division of Preventive Medicine, Brigham & Women's Hospital, Boston, Massachusetts; 50 - University of Virginia, Charlottesville, Virginia; 51 - Lundquist Institute, Torrance, California; 52 - National Taiwan University Hospital, National Taiwan University, Taipei; 53 - Cleveland Clinic, Cleveland Clinic, Cleveland, Ohio; 54 - National Health Research Institute Taiwan, Miaoli County; 55 - Metabolomics Platform, Broad Institute, Cambridge, Massachusetts; 56 - Immunity and Immunology, Cleveland Clinic, Cleveland, Ohio; 57 - University of Vermont, Burlington, Vermont; 58 - Population Health Science, University of Mississippi, Jackson, Mississippi; 59 - National Jewish Health, Denver, Colorado; 60 - Biostatistics, Boston University, Boston, Massachusetts; 61 - University of Texas Rio Grande Valley School of Medicine, Brownsville, Texas; 62 - Internal Medicine, University of Michigan, Ann Arbor, Michigan; 63 - Vitalant Research Institute, San Francisco, California; 64 - University of Illinois at Chicago, Chicago, Illinois; 65 - University of Chicago, Chicago, Illinois; 66 - Health Quantitative Sciences Research, Mayo Clinic, Rochester, Minnesota; 67 - Department of Medicine, Cardiovascular Division, Washington University in St Louis, St. Louis, Missouri; 68 - Human Genetics Center, Department of Epidemiology, Human Genetics, and Environmental Sciences, University of Texas Health at Houston, Houston, Texas; 69 - Vanderbilt University, Nashville, Tennessee; 70 - University of Cincinnati, Cincinnati, Ohio; 71 - University of North Carolina, Chapel Hill, North Carolina; 72 - University of Texas Rio Grande Valley School of Medicine, Edinburg, Texas; 73 - Genetics, Washington University in St Louis, St Louis, Missouri; 74 - Brown University, Providence, Rhode Island; 75 - Channing Division of Network Medicine, Harvard University, Cambridge, Massachusetts; 76 - Massachusetts General Hospital, Boston, Massachusetts; 77 - Center for Genes, Environment and Health, National Jewish Health, Denver, Colorado; 78 - Epidemiology, University of North Carolina, Chapel Hill, North Carolina; 79 - Washington University in St Louis, St Louis, Missouri; 80 - Fred Hutchinson Cancer Research Center, Seattle, Washington; 81 - New York Genome Center, New York City, New York; 82 - Icahn School of Medicine at Mount Sinai, New York, New York; 83 - Beth Israel Deaconess Medical Center, Boston, Massachusetts; 84 - Department of Psychiatry, Boston Children's Hospital, Harvard Medical School, Boston, Massachusetts; 85 - University of Texas Rio Grande Valley School of Medicine, San Antonio, Texas; 86 - University of Colorado Anschutz Medical Campus, Aurora, Colorado; 87 - Obstetrics and Gynecology, Mass General Brigham, Boston, Massachusetts; 88 - OB/GYN, Indiana University, Indianapolis, Indiana; 89 - Cardiology, University of Mississippi, Jackson, Mississippi; 90 - Medicine, University of Calgary, Calgary; 91 - Genetics, University of Maryland, Philadelphia, Pennsylvania; 92 - Department of Chronic Disease Epidemiology, Yale University, New Haven, Connecticut; 93 - Tulane University, New Orleans, Louisiana; 94 - Epidemiology, University of Washington, Seattle, Washington; 95 - Wake Forest Baptist Health, Winston-Salem, North Carolina; 96 - Channing Division of Network Medicine, Brigham & Women's Hospital, Boston, Massachusetts; 97 - University of Iowa, Iowa City, Iowa; 98 - Institute of Population Health Sciences, NHRI, National Health Research Institute Taiwan, Miaoli County; 99 - Tri-Service General Hospital National Defense Medical Center; 100 - Blood Works Northwest, Seattle, Washington; 101 - Taichung Veterans General Hospital

Taiwan, Taichung City; 102 - Internal Medicine, Division of Endocrinology, Diabetes and Metabolism, Oklahoma State University Medical Center, Columbus, Ohio; 103 - NHLBI, National Heart, Lung, and Blood Institute, National Institutes of Health, Bethesda, Maryland; 104 - Research Institute, Blood Works Northwest, Seattle, Washington; 105 - Biostatistics, University of Michigan, Ann Arbor, Michigan; 106 - Albert Einstein College of Medicine, New York, New York; 107 - Harvard University, Cambridge, Massachusetts; 108 - McGill University, Montreal; 109 - Epidemiology, University of Colorado at Denver, Aurora, Colorado; 110 - Medicine, Blood Works Northwest, Seattle, Washington; 111 - Public Health Sciences, Loyola University, Maywood, Illinois; 112 - Biostats, Harvard School of Public Health, Boston, Massachusetts; 113 - Medicine, University of Colorado at Denver, Aurora, Colorado; 114 - Boston University, Boston, Massachusetts; 115 - Harvard School of Public Health, Boston, Massachusetts; 116 - Epidemiology and Medicine, Brown University, Providence, Rhode Island; 117 - Cardiology, Duke University, Durham, North Carolina; 118 - Cardiovascular Institute, Stanford University, Stanford, California; 119 - The Charles Bronfman Institute for Personalized Medicine, Icahn School of Medicine at Mount Sinai, New York, New York; 120 - Division of Pulmonary, Critical Care and Sleep Medicine, Ohio State University, Columbus, Ohio; 121 - Broad Institute, Harvard University, Massachusetts General Hospital; 122 - cardiology, George Washington University, Washington, District of Columbia; 123 - University of Alabama at Birmingham, University of Alabama, Birmingham, Alabama; 124 - Epidemiology, Brown University, Providence, Rhode Island; 125 - Genome Sciences, University of Washington, Seattle, Washington; 126 - RTI International; 127 - Medicine, Massachusetts General Hospital, Boston, Massachusetts; 128 - University of Arizona, Tucson, Arizona; 129 - Center For Sleep Sciences and Medicine, Stanford University, Palo Alto, California; 130 - National Institute of Child Health and Human Development, National Institutes of Health, Bethesda, Maryland; 131 - Genes and Human Disease, Oklahoma Medical Research Foundation, Oklahoma City, Oklahoma; 132 - Ministry of Health, Government of Samoa, Apia; 133 - Howard University, Washington, District of Columbia; 134 - Department of Genome Sciences, University of Washington, Seattle, Washington; 135 - University of Maryland, Baltimore, Maryland; 136 - University at Buffalo, Buffalo, New York; 137 - Division of Sleep Medicine/Department of Medicine, University of

Pennsylvania, Philadelphia, Pennsylvania; 138 - Stanford Cardiovascular Institute, Stanford University, Stanford, California; 139 - Biochemistry, Wake Forest Baptist Health, Winston-Salem, North Carolina; 140 - University of Minnesota, Minneapolis, Minnesota; 141 - Biostatistics and Epidemiology Division, RTI International, Research Triangle Park, North Carolina; 142 - Department of Biostatistics, Boston University, Boston, Massachusetts; 143 - Fred Hutch and UW, Fred Hutchinson Cancer Research Center, Seattle, Washington; 144 - Cardiology/Medicine, Johns Hopkins University, Baltimore, Maryland; 145 - Medicine, University of Colorado at Denver, Denver, Colorado; 146 - CCPM, University of Colorado at Denver, Denver, Colorado; 147 - Genetics, University of North Carolina, Chapel Hill, North Carolina; 148 - Northwestern University, Chicago, Illinois; 149 - New York Genome Center, New York Genome Center, New York City, New York; 150 - Fred Hutchinson Cancer Research Center, University of Washington, Seattle, Washington; 151 - Lutia I Puava Ae Mapu I Fagalele, Apia; 152 - Sleep Research Unit, University of Ottawa Institute for Mental Health Research, University of Ottawa, Ottawa; 153 - Medicine, Pharmacology, Biomedical Informatics, Vanderbilt University, Nashville, Tennessee; 154 - Pediatrics, Lundquist Institute, Torrance, California; 155 - Faculdade de Medicina, Universidade de Sao Paulo, Sao Paulo; 156 - Pathology, University of Maryland, Seattle, Washington; 157 - TGPS, Lundquist Institute, Torrance, California; 158 - Division of Hematology/Oncology, Harvard University, Boston, Massachusetts; 159 - Genetics, Harvard Medical School, Boston, Massachusetts; 160 - Harvard Medical School, Boston, Massachusetts; 161 - Université Laval, Quebec City; 162 - Pediatrics, Emory University, Atlanta, Georgia; 163 - Human Genetics, Emory University, Atlanta, Georgia; 164 - Medicine/Cardiology, Vanderbilt University, Nashville, Tennessee; 165 - UMass Memorial Medical Center, Worcester, Massachusetts; 166 - University of Saskatchewan, Saskatoon; 167 - University of Michigan; 168 - Biostatistical Sciences, Wake Forest Baptist Health, Winston-Salem, North Carolina; 169 - Genomic Cardiology, University of Colorado at Denver, Aurora, Colorado; 170 - Channing Department of Medicine, Brigham & Women's Hospital, Boston, Massachusetts; 171 - Genetics, Stanford University, Stanford, California; 172 - Institute for Translational Genomics and Populations Sciences, Lundquist Institute, Torrance, California; 173 - University of Washington, Department of Genome Sciences, University of Washington, Seattle, Washington; 174 - Cancer

Prevention Division of Public Health Sciences, Fred Hutchinson Cancer Research Center, Seattle, Washington; 175 - Genetics, University of Pennsylvania, Philadelphia, Pennsylvania; 176 - Biostatistics, University of Alabama, Birmingham, Alabama; 177 - Department of Biostatistics, University of Washington, Seattle, Washington; 178 - Pathology & Laboratory Medicine, University of Vermont, Burlington, Vermont; 179 - USC Methylation Characterization Center, University of Southern California, University of Southern California, California; 180 - Brigham & Women's Hospital, Mass General Brigham, Boston, Massachusetts; 181 - Channing Division of Network Medicine, Department of Medicine, Brigham & Women's Hospital, Boston, Massachusetts; 182 - Epidemiology, Indiana University, Indianapolis, Indiana; 183 - Henry Ford Health System, Detroit, Michigan; 184 - Cardiology, Beth Israel Deaconess Medical Center, Cambridge, Massachusetts; 185 - Medicine, University of Pittsburgh, Pittsburgh, Pennsylvania; 186 - Department of Epidemiology, University of Michigan, Ann Arbor, Michigan; 187 - Department of Population and Quantitative Health Sciences, Case Western Reserve University, Cleveland, Ohio
